# A Phenotypic Paradigm for Cerebral Palsy Genetics

**DOI:** 10.64898/2026.01.13.25341946

**Authors:** Adam S Arterbery, Michael A Gargano, Anita Bagley, Jagadish Chandrabose Sundaramurthi, Lauren Rekerle, Thania Ordaz-Robles, Daniel Danis, Adam SL Graefe, Ana L Arenas-Díaz, Jeremy P Bauer, Hannah Blau, Leigh Carmody, Kristen l Carroll, Janice Davis, Philip F Giampietro, Anxhela Gjyshi Gustafson, Monserat Hernandez, Julius OB Jacobsen, Paige Lemhouse, David Millet, Shubhra Mukherjee, Patrick Nairne, Emily Nice, Talia Plotkin, Kenneth Powell, Ellen M Raney, Mallory Shingle, Damian Smedley, Peter A Smith, Demiana A. Soliman, David E Westberry, Jon R Davids, Peter N Robinson

## Abstract

Disease-causing genetic variants can be found in a subset of individuals with cerebral palsy (CP), with variants deemed causal of CP having been published for at least 515 genes. We develop a statistical approach that treats CP as a phenotypic feature for which some genetic disorders confer an increased risk. Based on comprehensive literature curation we show that the null hypothesis of no CP association can be rejected for only 89 of the 515 genes. We applied these findings to the analysis of a cohort of 460 children diagnosed with CP in the Shriner Children’s network. We identified pathogenic or likely pathogenic (P/LP) variants in 60 genes in 15.8% of the children. Only 16 of the 60 genes had significant evidence for CP association in our literature analysis. Our results suggest that a stratified approach to attributing causality to genetic variants in CP could support precision genomic medicine for affected individuals.

## Introduction

Cerebral palsy (CP) represents a clinically and etiologically heterogeneous group of permanent but not unchanging disorders of movement, posture, and motor function resulting from non-progressive disturbances of the developing fetal or infant brain^1^. It remains the most common cause of childhood physical disability, with a prevalence of approximately 1.6 to 3.6 per 1000 live births (0.16%-0.36%), a rate largely unchanged over the past half-century^2–8^. CP first manifests in infancy or early childhood, and the motor impairment is usually accompanied by delayed achievement of motor milestones. Diagnosis of CP is made based on a combination of findings from medical history, standardized neurological and motor assessment tools, and neuroimaging. Although motor defects are the cardinal feature, more than half of individuals with CP have associated comorbidities, including intellectual disability, autism spectrum disorder, epilepsy, language and communication impairment, and sensory deficits^9^.

Historically, CP was generally attributed to perinatal factors such as prematurity, infection, hypoxia–ischemia, and perinatal stroke, but population-based studies show these factors account for less than 10% of cases^10,11^. Recent studies have documented the role of Mendelian disease as a cause or risk factor for CP, with pathogenic single nucleotide and copy number variants being identified in a substantial subset of CP cases^12–18^. Some have referred to Mendelian diseases that present with a CP-like phenotype of early-onset, non-progressive motor impairment as “CP mimics”^19–21^. Others have argued that CP is a neurodevelopmental disorder diagnosed on clinical signs, not etiology, and that the diagnosis of CP should not be changed if a causative genetic variant is found^22^, and yet others have suggested that a CP diagnosis and an etiological genetic diagnosis should coexist^23^. The lack of consensus on the definition of CP and the role of genetics may be one of the reasons for the substantial variability in whether a child with a nonprogressive motor disability due to a genetic etiology will be diagnosed with CP^24^.

These observations prompted us to consider a model whereby CP is treated as a phenotypic feature of numerous other diseases. This model, if correct, would imply that the genetic architecture of CP is similar to that of phenotypes such as cleft palate that span both complex and Mendelian etiologies^25–27^. There is evidence for a complex genetic etiology of clefting in some individuals with numerous documented genetic and environmental risk factors,^28^ and on the other hand, Mendelian diseases with a strongly increased risk of orofacial clefting have been identified. For instance, 8 of 31 individuals with causal *HNRNPK* variants diagnosed with Au-Kline syndrome had cleft palate^29^; in all, 491 mainly Mendelian diseases are associated with *cleft palate* (HP:0000175) in the Human Phenotype Ontology^30^ annotation resource. Thus, similar to cleft palate, CP can occur in isolation or as a feature of Mendelian disease. A unifying conceptual framework for both CP and clefting is that environmental factors, high-effect variants seen in Mendelian disease, and common, low-effect variants with a modest association to the phenotype can all modify risk.

Conceptualizing the contribution of genetic variants to CP in terms of increased risk in this way implies that the association of a Mendelian disease with CP can be assessed using simple enrichment-based statistical tests. We hypothesized that not all the genes reported to harbor causal variants in individuals with CP in the medical literature are truly associated with CP. Further, we reasoned that an improved understanding of which genes are associated with CP could inform the interpretation of genetic findings in individuals with CP. In this work, we present a statistical analysis of a comprehensive selection of publications about diagnostic cohorts of individuals with CP and associated genes and Mendelian diseases. According to our model, the null hypothesis of no association can be rejected for 89 genes out of the 515 genes reported in individuals with CP in 21 published genomic diagnostic cohorts. Finally, we present results of genomic analysis of a cohort of 453 families with 460 children diagnosed with CP at Shriners Children’s Facilities. We sought pathogenic/likely pathogenic (P/LP) variants in the 515 genes and a set of other genes associated with Mendelian diseases that display some CP-like manifestations. Fewer than one-third of the genes in which P/LP variants were detected in our cohort show statistically significant evidence of association with CP. Our findings suggest that some previously proposed gene–CP relationships may not reflect a true etiologic link. Our approach has important implications for the interpretation of clinical sequencing results in children with CP and the development of precision genomic medicine approaches to leverage genetic findings to inform clinical care of affected individuals.

## Results

In this work, we investigated a cohort of 460 children (from 453 families) diagnosed with CP at Shriners Children’s Facilities. Shriners Children’s is a non-profit network of pediatric hospitals and medical facilities providing specialty care for orthopedic conditions, burns, spinal cord injuries, and craniofacial conditions. Treatment is offered to all children, regardless of a family’s ability to pay. The network delivers comprehensive services including surgery, rehabilitation, motion analysis, low-dose imaging, and orthotic/prosthetic care, supported by physical and occupational therapy.

### WGS analysis of 460 children diagnosed with CP

A total of 453 families with 460 children diagnosed with CP were recruited from eight care locations within the Shriners Children’s network (**Supplemental Table S1**). Trio WGS sequencing was performed where possible, but COVID regulations in force during much of the study stipulated that no more than one parent was allowed to accompany children to clinic visits (**Supplemental Table S2**). Probands and parents underwent WGS using the Illumina NovaSeq 6000 sequencing platform. Alignment and variant calling were performed using the DRAGEN Germline 3.9.5 pipeline (Illumina) aiming for a mean read depth of over 30-fold.

Human Phenotype Ontology (HPO) terms describing the clinical findings of each child were extracted using text mining from the clinical notes in the electronic healthcare records, and formatted as Global Alliance for Genomics and Health (GA4GH) phenopackets (Methods). We used these terms for Exomiser analysis of variant data from each family, focusing on results for genes previously associated with CP or with flagged P/LP variants.^31^ This led to a diagnosis in 70 (15.5%) of the 453 families with 460 affected children, corresponding to 72 affected children (15.8%; **Supplemental Figures S1-S70; Supplemental Tables S3-S4**).

The clinical characteristics of the study participants are shown in **Table 1**. There were higher rates of Seizure and different distributions of Gross Motor Function Classification System (GMFCS) and Communication Function Classification System (CFCS) scores among study participants for whom a P/LP variant was identified (“solved” group); proportionally more individuals in the solved group were in the more severe score bins IV and V.

**Table 1.** Clinical and demographic characteristics of study participants. Data are shown for participants in whom a variant was identified in a CP candidate gene (Solved) or not (Unsolved). Abbreviations: GMFCS-Gross Motor Function Classification System; CFCS-Communication Function Classification System; EDACS-Eating and Drinking Ability Classification System; MWU-Mann-Whitney U test. FET-Fisher exact test. The GMFCS score (I-V, coded 1-5) was compared between groups using a two-sided MWU (U = 15973.5, p = 0.0068; Median (Interquartile range [IQR]) — solved: 2.5 (2.0-4.0); unsolved: 2.0 (2.0-3.0). The CFCS score was compared analogously: U = 15138.0, p = 0.00073; Median (IQR) — solved: 2.0 (1.0-4.0); unsolved: 1.0 (1.0-2.0). The EDACS score was compared analogously: U = 13657.0, p = 0.046; Median (IQR) — solved 1: 1.0 (1.0-2.0); solved: 1.0 (1.0-2.0).

|  | Solved (n: 72) | Unsolved (n: 388) |
| --- | --- | --- |
| **Sex** |  |  |
| Male | 46 (64%) | 224 (58%) |
| Female | 26 (36%) | 164 (42%) |
| **Age at consent** |  |  |
| ≤ 4 years | 2 (3%) | 9 (2%) |
| 5 - 9 years | 24 (33%) | 133 (34%) |
| 10 - 14 years | 29 (40%) | 159 (41%) |
| > 15 years | 17 (24%) | 86 (22%) |
| **Gestational age at birth** (MWU, $p = 0.033$) | | |
| ≤ 27 weeks | 6 (12%) | 36 (16%) |
| 28 - 31 weeks | 10 (19%) | 58 (26%) |
| 32 - 36 weeks | 10 (19%) | 49 (22%) |
| > 37 weeks | 26 (50%) | 76 (35%) |
| **Seizures** (FET $p=0.0007$) | | |
| No | 7 (26%) | 32 (68%) |
| Yes | 20 (74%) | 15 (32%) |
| **GMFCS** (MWU: $p = 0.0068$) | | |
| I | 8 (11%) | 89 (23%) |
| II | 27 (39%) | 155 (41%) |
| III | 12 (17%) | 52 (14%) |
| IV | 13 (19%) | 56 (15%) |
| V | 10 (14%) | 30 (8%) |
| **CFCS** (MWU; $p = 0.00073$) | | |
| I | 31 (47%) | 243 (65%) |
| II | 5 (8%) | 49 (13%) |
| III | 8 (12%) | 26 (7%) |
| IV | 14 (21%) | 34 (9%) |
| V | 8 (12%) | 22 (6%) |
| **EDACS** (MWU; $p = 0.046$) | | |
| I | 40 (61%) | 264 (72%) |
| II | 12 (18%) | 50 (14%) |
| III | 2 (3%) | 24 (7%) |
| IV | 3 (5%) | 11 (3%) |
| V | 9 (14%) | 19 (5%) |

### What does it mean to be a CP-associated gene?

We then asked what the evidence is for association of variants in Mendelian-disease genes and how that would influence our interpretation of the genetic findings of our study. We reasoned that we could analyze published case and cohort reports under a null hypothesis of no association to assess the relevance of specific genes for CP. Available literature can be divided into two main categories: CP NGS cohorts and gene-specific cohorts.

We identified 21 published cohort studies on the genetics of CP in which groups of individuals diagnosed with CP were tested by next-generation sequencing (mainly exome and genome sequencing). Altogether, the cohorts included 5440 individuals with CP; of these, 1402 were solved by the identification of causal variants in a total of 515 genes. For our analysis, we merged the counts of solved and unsolved cases from these 21 cohorts; for brevity, we will refer to the combined cohort as the “CP NGS cohort”. Most genes (340 of the total of 515) were identified in only one cohort, but 106 were identified in 2 cohorts, 34 in 3 cohorts, and so on, with 1 gene identified in 14 cohorts. We tested for enrichment (higher than expected counts) based on a null hypothesis that genes reported as causal in the 1402 individuals were equally like to be any of a set of 1488 genes associated with early-onset manifestations of CP or neurodevelopmental delay in the HPO resource (**Methods**). Under this model, 10 genes were significantly enriched (**Figure 2A**; **Table 2**; **Supplemental Table S5**).

**Figure 1.**
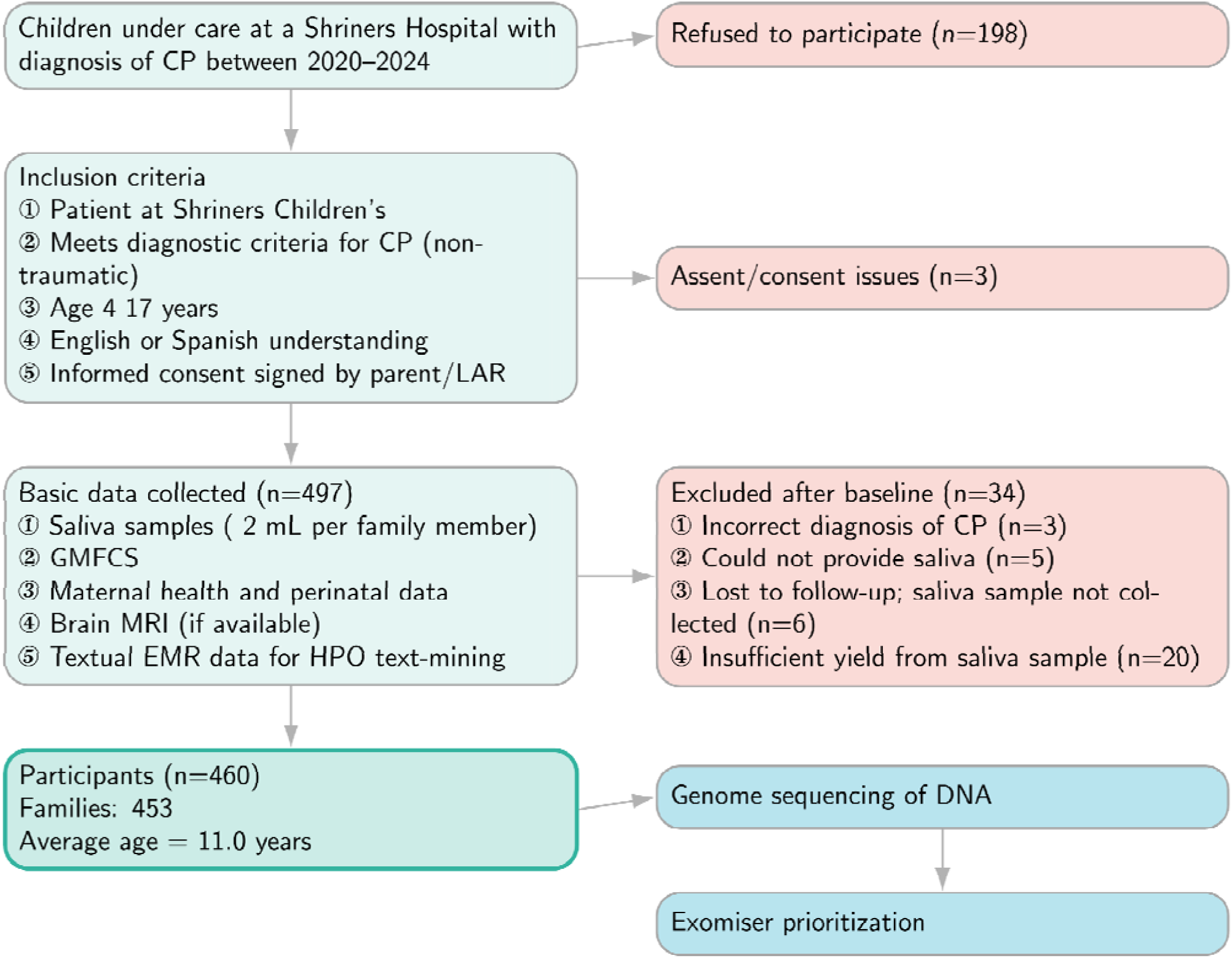
Overview of the study design. A total of 460 CP cases were recruited (453 families), and basic information collected. GMFSC: Gross Motor Function Classification System; LAR: Legally Appointed Representative; MRI: Magnetic Resonance Imaging; EMR: Electronic Medical Record.

**Figure 2.**
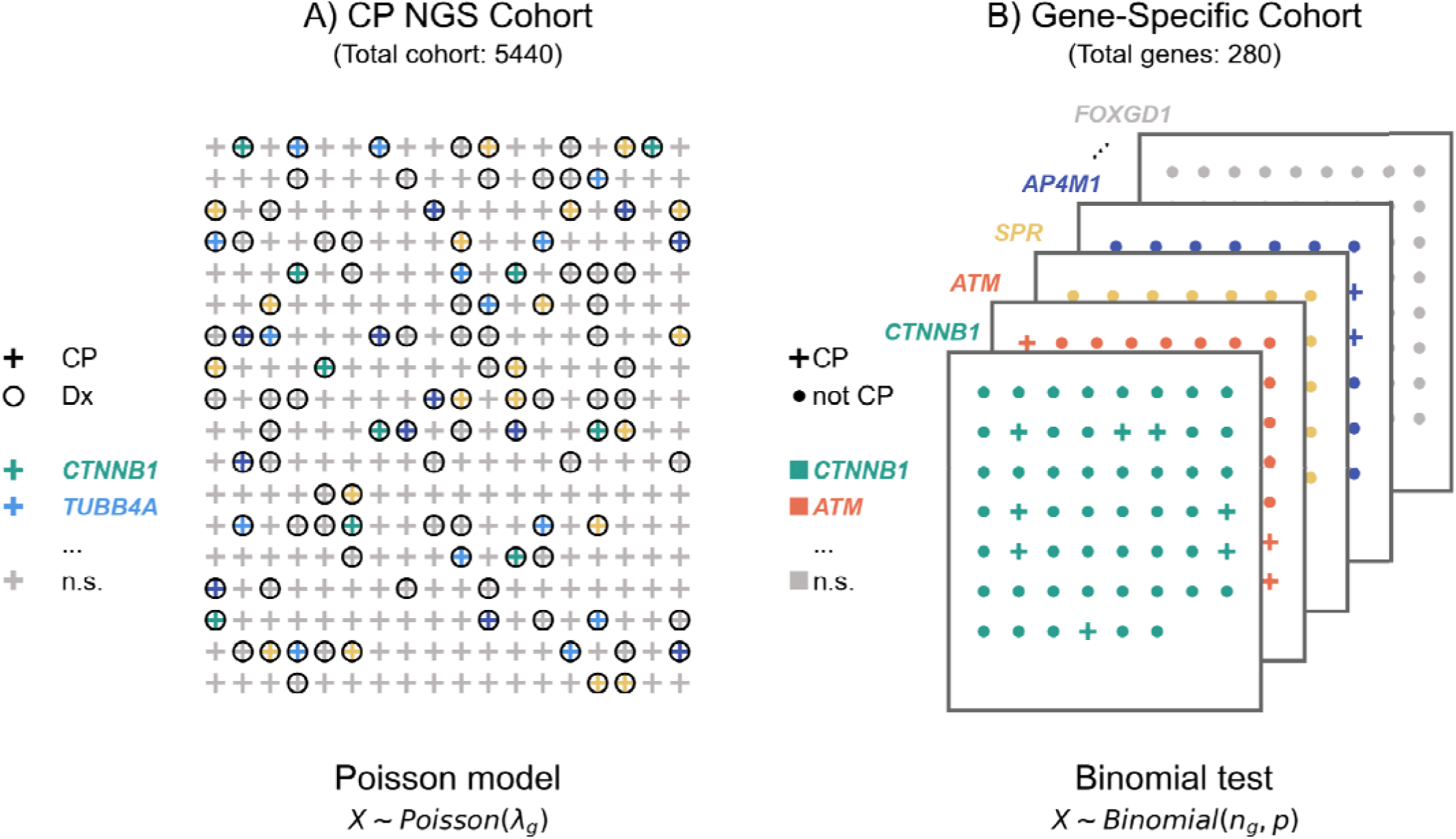
Statistical analysis for association with CP. Two approaches were applied to assess the association of pathogenic variation in genes with CP. **(A) Poisson test of CP NGS Cohort Data.** We curated 21 previously published cohorts of individuals with CP who underwent diagnostic next-generation sequencing and calculated the total number of times each gene was deemed to be causally related to CP by the original authors (a total of 515 genes were observed at least once). Under a null hypothesis that if these 515 genes were unrelated to CP, then their relative frequencies (λ*_g_*) among the 1402 individuals given a diagnosis should reflect the chance of being chosen from a set of genes associated with early-onset manifestations of CP. All individuals in this cohort have CP and are shown as “+”; in some cases, the individuals were diagnosed with a genetic disease (“Dx”) (shown as a circle around the “+”). Some of these genes showed significant enrichment in the cohort and are shown in color. If no genetic diagnosis was made, no circle is shown. **(B) Binomial test of 280 gene-specific cohorts.** We comprehensively curated published literature on CP candidate genes, and recorded how many individuals were described as having CP (colored circles) in the entire cohort (). Under a null hypothesis of no association one can model the CP count as a binomial distribution with a parameter () of 0.3% based on the population prevalence of CP. One such test was performed for each of 280 curated gene specific cohorts. In each of these cohorts, all individuals are reported to have pathogenic variation in the gene of interest (e.g., *CTTNB1*). Some of the individuals were diagnosed with CP (shown as “+”), others were not (shown as dots). Genes showing significant enrichment are shown in color, and the remaining genes are shown in grey.

**Table 2.**
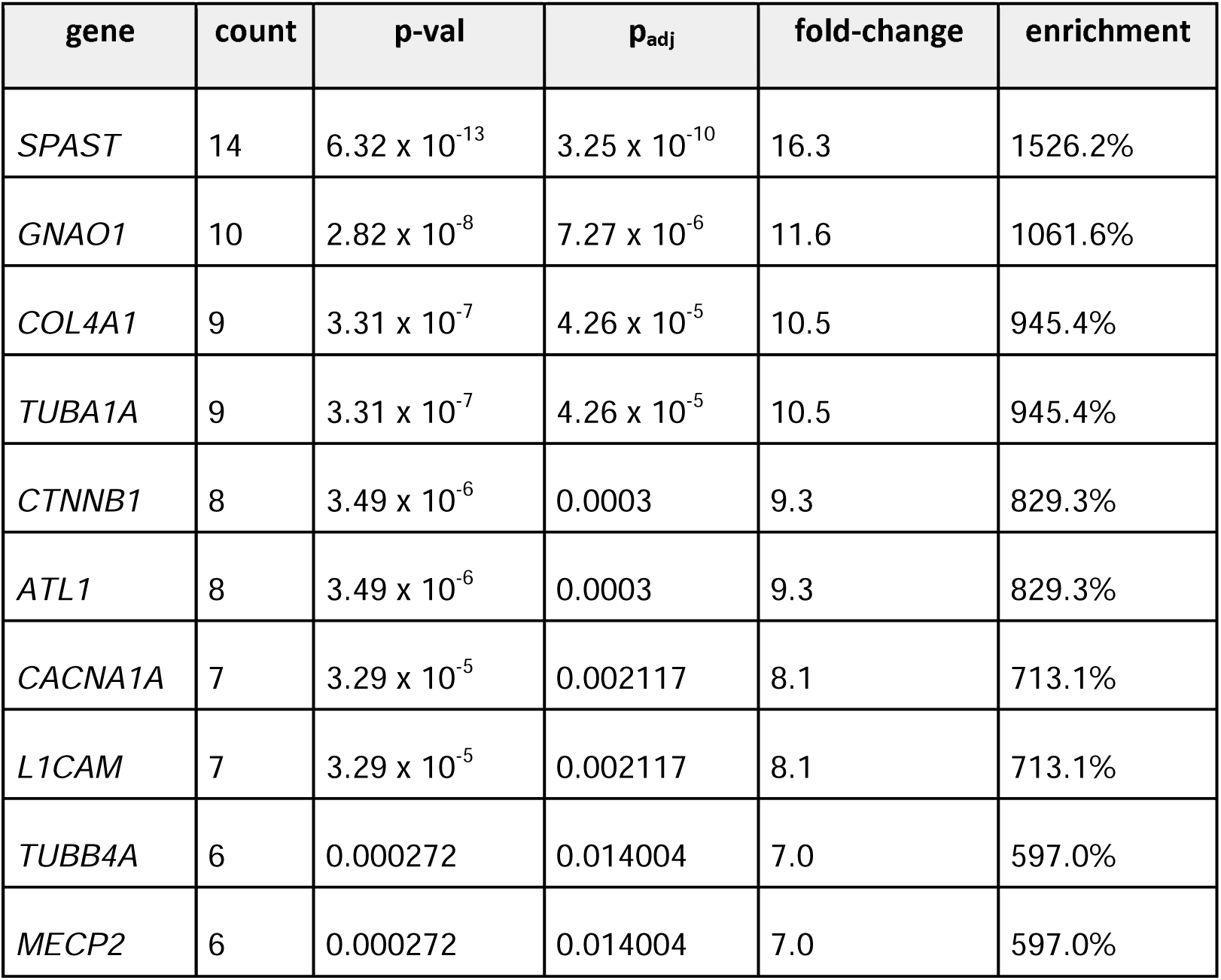
Enriched genes in the 21 CP NGS cohorts. A Poisson test for enrichment was conducted for each of the 515 genes identified as causal in one or more of the 21 cohorts. The null hypothesis was that any of 1488 candidate CP genes (**Methods**) was equally likely to be identified in the 1402 solved cases, corresponding to a lambda of 0.942. “Count” shows the total number of individuals reported to have causal variants in the given genes in the 21 cohort articles. The fold-change column shows the relative enrichment that was observed compared to this expectation, and the enrichment column shows the corresponding percent enrichment. p-val: Upper-tail p-values were calculated from a Poisson model comparing the observed counts to the expected rate. p_adj_: P values were adjusted for multiple testing using the Benjamin Hochberg method (total of 515 tests).

Data from the 21 CP NGS cohorts is obviously biased because only individuals diagnosed with CP were included in the cohorts. We hypothesize that some of these associations may have arisen by pure chance. For instance, only a single individual with pathogenic *LIPH* variation was reported in in one of the 21 CP NGS cohorts.^12^ *LIPH* variants in this gene are associated with the disease hypotrichosis 7 (MIM:604379), which has no neurological or other manifestations clearly associated with CP. On the other hand, of 171 individuals with *ITPR1* variants, six were diagnosed with cerebral palsy^32^. We reasoned that the probability of observing k cases of CP among n individuals reported with a variant in a given gene and with the null hypothesis that CP occurs at its background population frequency (here taken as 0.3%; see Methods for data sources and rationale) can be modeled according to a binomial distribution. Applying this framework, *ITPR1* shows a statistically significant enrichment of CP among variant carriers, whereas *LIPH* does not.

We therefore applied a binomial test to 280 genes chosen from the 515 genes reported in the CP NGS cohorts including all 175 genes that were reported in two or more of the CP NGS cohorts. Data across 280 genes was derived from 5508 publications describing 43,897 individuals, 692 of whom were reported to have CP (**Supplemental Table S6)**. Of note, the 21 publications describing the CP NGS cohorts were omitted from this analysis. We then assessed whether there was sufficient evidence to reject the null hypothesis of no association based on a binomial test corrected for multiple testing by the Bonferroni method (Methods). 86 of 280 tested genes showed sufficient evidence to reject the null hypothesis of no association (binomial test with Bonferroni correction for multiple testing; **Table 3**; **Supplemental Table S7; Figure 2**).

**Table 3.** Top 20 enriched genes from the 277 gene-specific cohorts. Publications other than the 21 CP NGS cohort articles were curated for diseases associated with the genes indicated in the first column. The number of individuals reported to have CP is shown in the second column, and the number of separate publications reporting at least one individual with a causal variant in the indicated gene who was reported to have CP is shown in the third column. The fourth column shows the corresponding p-value (Binomial test). Fold-change and enrichment are shown as in Table 2. In all, 86 of 277 tested genes were statistically significant. A complete list is available in **Supplemental Table S7**.

| gene | total | pubs | pval | fold-change | enrichment |
| --- | --- | --- | --- | --- | --- |
| *CTNNB1* | 55/405 (13.6%) | 8/54 | $2.69 \times 10^{-71}$ | 45.3 | 4426.7% |
| *ATM* | 33/198 (16.7%) | 4/6 | $1.45 \times 10^{-46}$ | 55.6 | 5455.6% |
| *SPR* | 20/55 (36.4%) | 2/7 | $1.59 \times 10^{-36}$ | 121.2 | 12021.2% |
| *AP4M1* | 15/27 (55.6%) | 4/12 | $2.41 \times 10^{-31}$ | 185.2 | 18418.5% |
| *PLP1* | 24/280 (8.6%) | 11/44 | $4.26 \times 10^{-27}$ | 28.6 | 2757.1% |
| *KANK1* | 14/42 (33.3%) | 5/7 | $2.34 \times 10^{-25}$ | 111.1 | 11011.1% |
| *PDHX* | 14/52 (26.9%) | 1/14 | $7.61 \times 10^{-24}$ | 89.7 | 8874.4% |
| *STXBP1* | 15/95 (15.8%) | 2/4 | $1.26 \times 10^{-21}$ | 52.6 | 5163.2% |
| *COL4A1* | 23/508 (4.5%) | 13/108 | $9.35 \times 10^{-20}$ | 15.1 | 1409.2% |
| *SLC6A3* | 10/28 (35.7%) | 3/8 | $7.38 \times 10^{-19}$ | 119.0 | 11804.8% |
| *SLC16A2* | 14/112 (12.5%) | 7/26 | $8.73 \times 10^{-19}$ | 41.7 | 4066.7% |
| *GNAO1* | 18/279 (6.5%) | 13/64 | $1.73 \times 10^{-18}$ | 21.5 | 2050.5% |
| *DYRK1A* | 14/220 (6.4%) | 3/35 | $1.26 \times 10^{-14}$ | 21.2 | 2021.2% |
| *AUTS2* | 12/142 (8.5%) | 4/27 | $3.23 \times 10^{-14}$ | 28.2 | 2716.9% |
| *NT5C2* | 7/31 (22.6%) | 3/8 | $5.4 \times 10^{-12}$ | 75.3 | 7426.9% |
| *RAB3GAP1* | 9/91 (9.9%) | 2/23 | $1.24 \times 10^{-11}$ | 33.0 | 3196.7% |
| *KAT6A* | 10/167 (6.0%) | 1/29 | $1.36 \times 10^{-10}$ | 20.0 | 1896.0% |
| *CACNA1A* | 19/1064 (1.8%) | 6/191 | $1.35 \times 10^{-9}$ | 6.0 | 495.2% |
| *ATP1A3* | 10/220 (4.5%) | 2/11 | $1.98 \times 10^{-9}$ | 15.2 | 1415.2% |
| *GCDH* | 11/290 (3.8%) | 3/27 | $2.08 \times 10^{-9}$ | 12.6 | 1164.4% |

### Significantly enriched genes identified in the Shriners Children’s Cohort

As mentioned above, P/LP variants were identified in 70 families in the Shiners cohort, affecting 60 distinct genes (**Supplemental Table S3**). We first asked if these genes showed more overlap than would be expected by chance when comparing the enriched genes identified by the above two analysis approaches. We use a Fisher exact test to assess the degree of overlap between the union of genes identified as enriched in the CP NGS cohort and the gene-specific cohort analyses (the union of the 10 genes from the CP NGS cohort and the 86 genes from the gene-specific cohorts was 89 genes) with the 60 genes identified in our analysis, using all Mendelian disease-associated genes (5441 genes derived from HPO project annotations). 16 of the 60 genes overlapped, corresponding to a 16.3 fold enrichment over the overlap expected by chance (Fisher Exact p=5.46 x 10^-16^). Of the 16 overlapping genes, six genes were significant in both the CP NGS cohort and the gene-specific cohort analysis (**Figure 3**).

**Figure 3.**
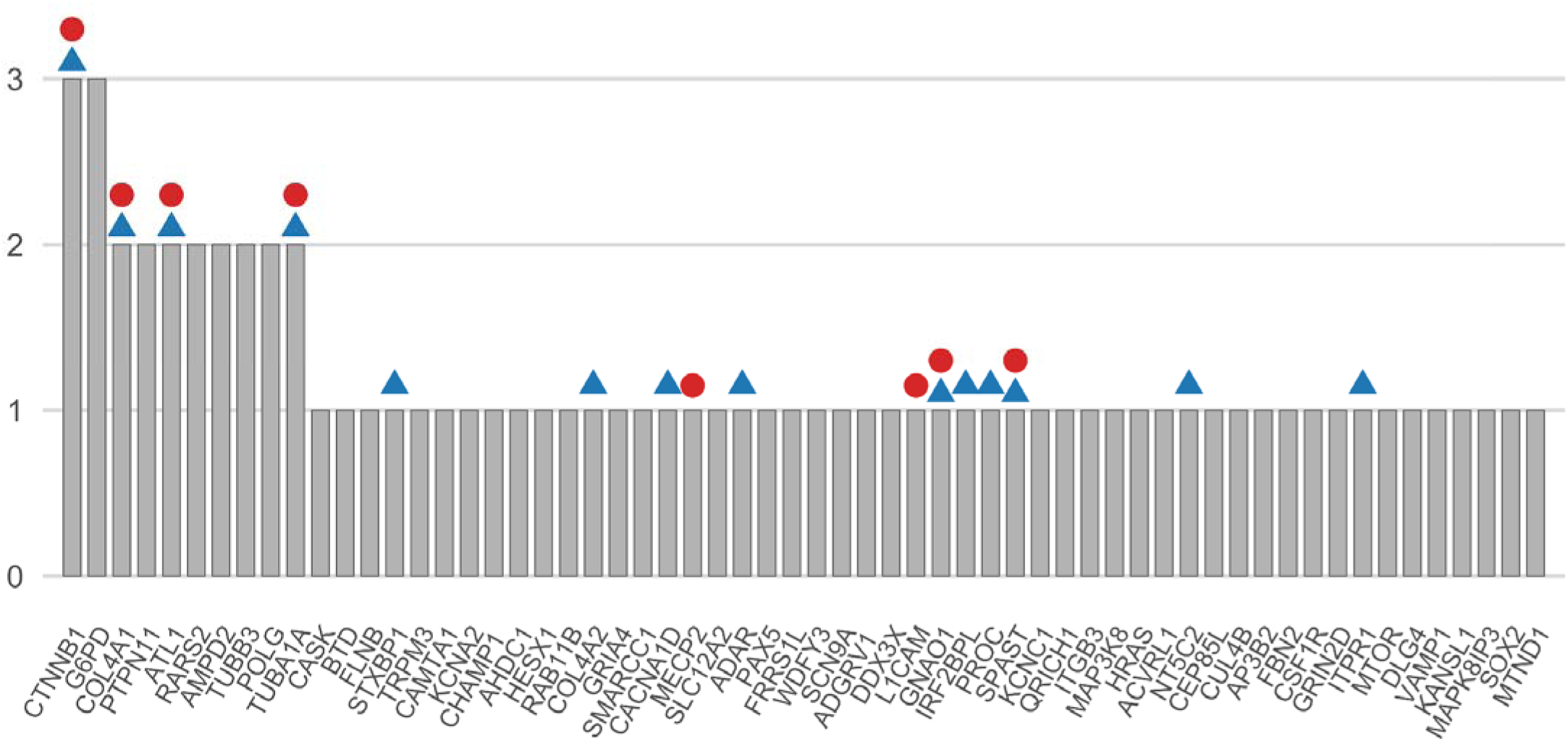
The 60 genes with P/LP-variants in CP candidate genes. P/LP variants were identified in a total of 60 unique genes in 73 children from 70 families. Count refers to the number of families identified with a variant in each gene. A circle above the gene signifies that the gene was enriched in our cohort analysis (Poisson test), and a triangle signifies that the gene was enriched in our gene-specific analysis (Binomial test).

## Discussion

In this study, we aimed to investigate how much evidence is available for the association of certain Mendelian disease-associated genes and CP. We evaluated 21 published CP NGS cohorts in which a total of 515 genes were interpreted to harbor causal variants. We analyzed 5508 additional published articles about individuals or cohorts with diseases associated with these genes and identified evidence sufficient to reject the null hypothesis of no CP association for only 89 of the 515 genes.

Genomic diagnostic investigations are now commonly employed in the workup of children with a phenotype of CP. In our study of 453 families with one or more children diagnosed with CP, we identified P/LP variants in 70 (15.5%) of the 453 families, comprising 60 unique genes. Based on our statistical analysis, evidence for a causal role exists for only 16 of the 60 genes.

To promote precision genomic medicine for CP, it is important to understand the mechanisms by which genetic diseases can be associated with CP. For instance, Glucose-6-phosphate dehydrogenase (G6PD) deficiency can be associated with prolonged neonatal jaundice, which if untreated can result in kernicterus and spastic CP^33^. In resource-poor clinical settings, lack of access to common treatments for hyperbilirubinemia implies a continued risk of CP for individuals with G6PD deficiency^34^, but the three boys in our study with P/LP *G6PD* variants had no history of kernicterus, and we interpret the *G6PD* findings as incidental rather than causal. The population prevalence of rare genetic diseases may be as high as about 5%^35,36^. Therefore, it is possible that many genetic diseases identified in individuals with CP are not causally related to the CP. For instance, one individual with pathogenic variation in the *LIPH* gene was reported in a cohort of 1345 individuals with CP who underwent diagnostic exome sequencing^12^. Biallelic P/LP variants in this gene are associated with the disease hypotrichosis 7, or woolly hair with or without hypotrichosis (MIM:604379), which have no neurological or other manifestations clearly associated with CP. On the other hand, a range of statistical evidence exists to associate certain genetic diseases with CP; in our study, for instance, we identified P/LP *CTNNB1* variants in three children with CP; causal *CTNNB1* variants were also identified in 8 of the 21 CP NGS cohorts we analyzed (p=2.41 x 10^-11^) and in 55/405 individuals reported in other publications about *CTNNB1* from the medical genetics literature (p=7.46 x 10^-69^) (**Supplemental Tables S5 and S7**). Therefore, it appears much more plausible that *CTNNB1* variants increase risk for CP than for variants in *LIPH*.

There are several ways in which a better understanding of the genetic etiology of CP could improve clinical management. Genomic newborn screening has the potential to detect the majority of genetic diseases in a single assay, and several clinical studies are exploring its utility and related ethical concerns.^37,38^ Additionally, whole exome or genome sequencing may be performed diagnostically at different ages for a number of reasons. If a P/LP variant is found in a gene known to be associated with a significantly increased risk for CP, targeted interventions and preventive measures can be initiated at an earlier stage.^39^

Early intervention for children younger than two years with or at risk of CP is critical. Current clinical practice recommends screening infants at high risk of CP (for example, following preterm birth or in infants with perinatal intraventricular hemorrhage) and instituting appropriate measures if the diagnosis of CP is made.^40,41^ Therefore, clinicians may consider whether children with P/LP variants in CP-associated genes should be assessed for the degree of CP risk and be referred to CP specialist centers if an above average risk is determined. Future clinical studies will be required to determine the potential clinical benefit of this strategy.

General treatment options for children with CP include physical therapy, botulinum toxin, orthotics, and orthopedic surgery.^42^ Options for disease-specific treatment for CP include some inborn errors of metabolism that can present with CP,^20,43–45^ hereditary bleeding disorders such as hemophilia and factor XIII deficiency,^46,47^, hereditary disorders of movement associated with dystonia,^48,49^ pyridoxine-dependent epilepsy,^50^ Coenzyme Q10 deficiency,^51^ mitochondrial disorders^20^ and others. Recent estimates state that actionable findings are revealed by genomic analysis in approximately 8.5% to 24% of cases with a genetic diagnosis.^18,20,39,52^

Additionally, a major motivation for parents of children with CP to have genomic diagnostics is the potential to end the diagnostic odyssey, to optimize patient management and reduce unnecessary physician consultation and diagnostic testing. Potentially, improved understanding of the etiology of CP will prove helpful for understanding natural history and response to treatments. In some cases, a diagnosis provides families with access to health care programs and services or specialized support groups.^53^ It is therefore essential to evaluate whether the finding of a P/LP variant in a child with CP truly justifies the inference of causality for the CP phenotype. According to the clinical situation, it may be appropriate to continue the diagnostic workup if a P/LP variant is identified in a gene that is not significantly associated with an increased risk of CP. Depending on the clinical context, it may be appropriate to continue the diagnostic workup when a P/LP variant is identified in a gene with only limited or uncertain association with CP, under the assumption that another etiology may be present and that identifying it could further guide clinical management. However, it should be acknowledged that our current state of knowledge on this issue is incomplete, and patient-specific, individualized decisions should be made according to clinical considerations.

Limitations of our approach towards assessing the association of Mendelian disease-associated genes with CP include variability in how the clinical diagnosis of CP is made,^54^ as well as forms of publication bias that may lead to a tendency for more severe cases to be published.^55^ Additionally, although we have endeavored to comprehensively curate publications with case reports in the genes of interest, some publications could not be accessed due to access restrictions, and some publications may not have presented all findings for a given case. We identified evidence of CP association for only 89 of the 515 genes analyzed, but are not able to rule out that more genes have a true association. An ideal dataset for examining CP association of a gene associated with one or more Mendelian diseases would consist in a sufficiently large cohort of individuals who had been examined by geneticists and CP specialists and for whom detailed phenotypic information had been recorded using standards such as HPO and GA4GH phenopackets. Despite the above limitations, our literature analysis does suggest that Mendelian disease-associated genes may be associated with a spectrum of increased risk for CP (as suggested by the spectrum of percentage enrichment scores in **Tables 2** and **3** and **Supplemental Tables S5** and **S7**).

Further retrospective investigations and ideally prospective clinical trials will be required to determine the clinical utility and cost-effectiveness of integrating genomic diagnostics into routine CP care and to identify which individual subgroups benefit most from early genetic testing. To understand the natural history of rare diseases that are significantly associated with CP and to define the role of precision medicine approaches to clinical care of affected children, it will be necessary to acquire deep phenotypic data and structured information on relevant clinical outcomes. Major roadblocks include concerns about EHR data quality and completeness, only partial integration of genomics data in EHR, and the lack of widespread data sharing to power statistically significant connections between phenotypes and genotypes. The latter could be addressed by widespread adoption of standards from the Global Alliance for Genomics and Health, which enable standardized and interoperable data formats for federated, secure discovery and sharing of EHR-derived data using the Fast Healthcare Interoperability Resources (FHIR).^56–59^ Widespread adoption of the GA4GH Phenopacket Schema for reporting clinical findings in publications would substantially increase the value of published literature for our understanding of the natural history of rare disease and cerebral palsy. Improved data interoperability would enable routine clinical and registry data to be shared and integrated across many other disease groups as well.

## Conclusions

We have proposed a statistical approach to assess whether a gene is associated with an increased risk of CP. We have shown that insufficient evidence exists to reject the null hypothesis of no CP association for a majority of genes flagged as causal in previous CP NGS cohort studies, as well as for most of the genes found to harbor P/LP variants in our study. Our approach is based on a paradigm that considers CP to be a disease component (phenotypic feature) rather than a precise disease diagnosis, a viewpoint that is in line with previous opinions about how to define CP^22^. The results of our study suggest that comprehensive collections of detailed clinical data could be used to support translational research, genomic diagnostics, and potentially precision clinical management of individuals with CP.

## Methods

### Cohort

Inclusion criteria for the study stipulated that study participants were individuals between 4 and 17 years of age, patients at a Shriners Children’s facility, and had a current diagnosis of CP, not caused by a traumatic brain injury. The participant or the legally authorized representative (LAR) must be able to read and/or understand English or Spanish. Exclusion criteria comprised physical or mental limitation(s) that would prevent the collection of the saliva specimen (e.g. xerostomia, facial abnormalities) and history of a diagnosis of any neurodevelopmental disorders other than CP.

The study aimed to enroll 500 patients and both parents (biological trios), for a total sample size of 1500 whole genome sequencing (WGS). Owing to COVID regulations that stipulated that only one parent was allowed to accompany a child to clinic visit, trios were not available for all patients. The study began in 2021 with five Shriners Children’s (SC) sites: Chicago, Greenville, Northern California, Portland, and Shreveport. In 2022, SC Philadelphia and SC Salt Lake City were added, and in 2023 SC Mexico City was added (**Supplemental Table S1**). A total of 453 families with 463 children diagnosed with CP were successfully recruited into the study. Two of the individuals reported here were previously described in case reports.^32,60^

### Ethical considerations

The study was approved by the Institutional Review Board of Shriners Children’s (WCG IRB Protocol #20212489). Additionally, ethical approval for this study was granted by the Ethics Committee at Shriners Children’s Mexico City. Approval No: CEI-2023-01. Written consent was generally obtained from both parents of the child whose biosamples were used in this study, but due to COVID restrictions that entailed that only one parent could be present, in some cases only one parent signed.

### Encoding clinical data using Human Phenotype Ontology and Global Alliance for Genomics and Health Phenopackets

The Human Phenotype Ontology (HPO) provides a standardized vocabulary of 19,414 terms (version 2025-11-24) that describe the phenotypic abnormalities of human disease. Additionally, the HPO provides a comprehensive corpus of phenotype annotations (HPOA) that form computational models of 8534 rare diseases. HPO applications include genomic interpretation for diagnostics, gene-disease discovery, machine learning (ML) and electronic health record (EHR) cohort analytics. HPO analysis enables fuzzy, specificity weighted phenotype profile matching that assists in the analysis of human diseases and phenotypes by offering a computational bridge between genome biology and clinical medicine^61–63^.

The Global Alliance for Genomics and Health (GA4GH) Phenopacket Schema provides a standard schema for sharing clinical and genomic information about an individual, including phenotypic descriptions, numerical measurements, genetic information, diagnoses, and treatments^64^, and is designed to work with HPO terms. For the current project, electronic health records (EHR) for the affected children were deidentified, extracted, and exported into comma-separated file format. Text mining was performed using fenominal, an HPO text-mining application^65^. HPO terms and other metadata were extracted from the deidentified EHR records using a number of heuristics to reduce false-positive results (e.g., texts related to family history or to warnings about medication effects are skipped; for HPO terms that were called as both observed and excluded at different encounters, the term was recorded as excluded if more than half of the individual occurrences were recorded as excluded). Finally, clinical information was formatted according to the Global Alliance for Genomics and Health (GA4GH) Phenopacket Schema^64^. The phenopackets were used as input files for further analysis as described below.

### Whole-Genome Sequencing

Probands and, if available, their parents underwent WGS using the Illumina NovaSeq 6000 sequencing platform (Illumina). Libraries were constructed using the Illumina DNA PCR-Free Prep Tagmentation kit (Illumina) for paired-end parallel sequencing (2 × 150 bases) aiming for an average coverage of 30 fold. DRAGEN Germline Whole Genome pipeline was used with default settings for alignment and variant calling.

### Variant Prioritization

We analyzed the WGS data using Exomiser (version 14.1.0 and data version 2502), a bioinformatics software comprising a suite of algorithms for prioritizing disease-gene variants using random-walk analysis of protein interaction networks, clinical phenotype comparison with known patients based on HPO terms, and cross-species phenotype comparisons, as well as a wide range of other computational filters for variant frequency, predicted pathogenicity, and pedigree analysis^66^. For variants contributing to the final combined score, Exomiser applies a set of rules to automatically assign ACMG categories based on the variant frequency (PM2, BA1), pathogenicity scores (PP3, BP4), other computational and predictive data (PVS1, PS1, PM4, PM5), functional data (PM1), segregation (BS4) and de novo data (PS2), allelic data (PM3, BP2), prior clinical reports in ClinVar (PP5, BP6), and finally phenotypic data (PP4). Exomiser will assign PP4 up to PP4_Moderate in cases where there is a strong phenotype match score (>= 0.6). The assigned ACMG categories are combined using the updated Bayesian scoring framework^67^ to calculate the final classification of Pathogenic / Likely Pathogenic / VUS / Likely Benign / Benign.

### Curation of CP NGS Cohorts

We focused our analysis on genes proposed to be candidate cerebral palsy-related genes in 21 previously published studies that involved diagnostic genomic analysis of cohorts of individuals diagnosed with CP^3,12,17,18,49,68–83^, which for conciseness we will refer to as “CP NGS cohorts”. We counted how many times variants in these genes were deemed causal or associated with a patient’s diagnosis of CP by the authors of the studies (**Supplemental Table S8**).

### Curation of Gene-Specific Cohorts

The CP NGS cohorts have an obvious bias, because an inclusion criterion of all of the cohorts was that each study participant be diagnosed with CP. We reasoned that some of the 515 genes identified in the CP NGS cohorts might not have a true association with CP given the population prevalence of CP. Therefore, referring to the 515 genes as “CP candidate genes”, we then compiled a comprehensive selection of published case and cohort reports about CP candidate genes as follows. Publications mentioned in the Molecular genetics section of the OMIM entries for the diseases associated with the gene were included. We additionally identified publications using the PubMed query “<GENE symbol>[title] AND (variant or mutation)”. Additional publications were identified by inspection. For each published case or cohort we counted each individual explicitly diagnosed with “cerebral palsy”. Additionally, individuals were counted as being diagnosed with CP if the publication described that they were being cared for in a CP clinic, e.g., “established care in a cerebral palsy clinic,” or if the initial clinical suspicion was reported as being CP.

We curated all 172 CP candidate genes that were reported in at least two of the 21 NGS cohorts (omitting only *TENM1*, *KDM7A*, *NAA35*, which were identified in two cohorts each but are not listed as disease associated genes in OMIM). We curated an additional 105 candidate CP genes that had been reported in only one NGS cohort. 5482 publications related to these 277 genes were curated, comprising a total of 43,779 individuals (692 with CP). For each gene, the number of individuals reported to have CP as well as the total number of individuals with reportedly causal variants in the genes were recorded (**Supplemental Table S6)**. For conciseness, we refer to each of the 277 cohorts as “gene-specific cohorts”.

### Statistical analysis of enrichment of CP-associated genes

Our hypothesis is that genes that are repeatedly observed to harbor pathogenic variants in individuals with CP are more likely to have a causal relation to CP than genes that are rarely found to harbor such variants. We acknowledge that curated datasets described above are not fully adequate for rigorously testing this hypothesis. Publications about cohorts of individuals with CP have an obvious ascertainment bias, and therefore, the mere observation of a case with a pathogenic variant in a certain gene does not necessarily mean there is a causal relationship between the variant and the diagnosis of CP. Furthermore, the general medical genetics literature about individuals with variants in specific genes is subject to issues with data quality and comprehensiveness, a bias to publish severely affected individuals, and variation in practices for diagnosing CP. Nonetheless, we argue that this is the best currently available data for our purpose. The following sections describe the statistical methodology we used.

### Statistical analysis of CP NGS cohorts

Together, the 21 CP NGS cohorts comprised 5440 individuals with CP; 1281 were solved, corresponding to an average diagnostic yield of 23.5% (**Supplemental Figure S71**; **Supplemental Table S3**). The diagnosed cases involved a total of 515 genes. We reasoned that if a gene was reported in many individuals, this can be taken as evidence of a true CP association. For instance, 340 of the 515 genes were reported in only a single individual. In contrast, the most commonly reported gene was *SPAST*, variants in which were reported in 29 individuals from 14 of the 21 cohorts.

Our null hypothesis is that the P/LP variants observed in these studies have nothing to do with the CP diagnosis, but rather are related to other randomly selected Mendelian diseases. We defined a null model based on a set of potentially CP-associated genes, which we defined as all genes associated with diseases with infantile or childhood onset that are annotated to the HPO terms *Cerebral palsy* (HP:0100021; n=32), *Spasticity* (HP:0001257; n=441), *Hypertonia* (HP:0001348; n=52), *Dystonia* (HP:0001332; 266), *Ataxia* (HP:0001251; n=435), or Neurodevelopmental delay (HP:0012758; n=1258). In each case, diseases annotated to either one of these terms or to a more specific descendent of the terms were included. Together with the 515 candidate genes derived from the CP NGS cohorts, this corresponded to a total of 1488 genes (the total is less than the sum of the genes associated with each HPO term because of overlaps). The null hypothesis posits that each of the 1281 individuals in the 21 cohorts with solved cases was equally likely to harbor a causal genetic variant in one of the 1488 candidate genes. This leads to a lambda of 1281/1488 = 0.861. With this, we calculated a Poisson upper-tail p-value as

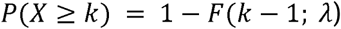

where F(k; λ) denotes the cumulative distribution function of a Poisson random variable with rate λ. One test was performed for each of the 515 genes identified in the CP NGS cohorts; Benjamini-Hochberg correction for multiple testing was applied.^84^ The fold-change (fc) was calculated as the observed count divided by λ, and the percent enrichment was calculated as (fc - 1) * 100.

### Statistical analysis of gene-specific cohorts

For each gene-specific cohort we performed a binomial test for enrichment. The null hypothesis was that the number of individuals diagnosed with CP in the cohort is not more than would be expected given the population prevalence of CP and the size of the cohort. A range of prevalence estimates for CP has been published in the literature, running from 1.6, 2, or 2 to 3.6 per 1000 (0.16%-0.36%).^2–8^ For the analysis presented here, we used p = 0.003 (i.e., an overall frequency of CP in the general population of 0.3%). The choice of a relatively high estimate is conservative. Then, for each gene cohort, we take the cohort size to be the number of curated individuals, and the number of events as the number of those individuals diagnosed with CP (x). In R, this is dbinom(x=x, size=cohort_size, p=p). No correction for multiple testing was applied since the data for each gene-specific cohort were independent of each other. Fold-change and percent enrichment were calculated as above.

## Data Availability

De-identified individual participant data are available in the text, tables and figures of the article. We do not have consent to share the original EHR and genomic sequencing data. Requests for further information should be directed by email to the corresponding author and will be responded to promptly.

## Weblinks

Exomiser: https://github.com/Exomiser

Human Phenotype Ontology: https://hpo.jax.org/

Global Alliance for Genomics and Health Phenopacket Schema: https://github.com/ga4gh/phenopacket-schema

