## Supplemental figures for "A Phenotypic Paradigm for Cerebral Palsy Genetics"

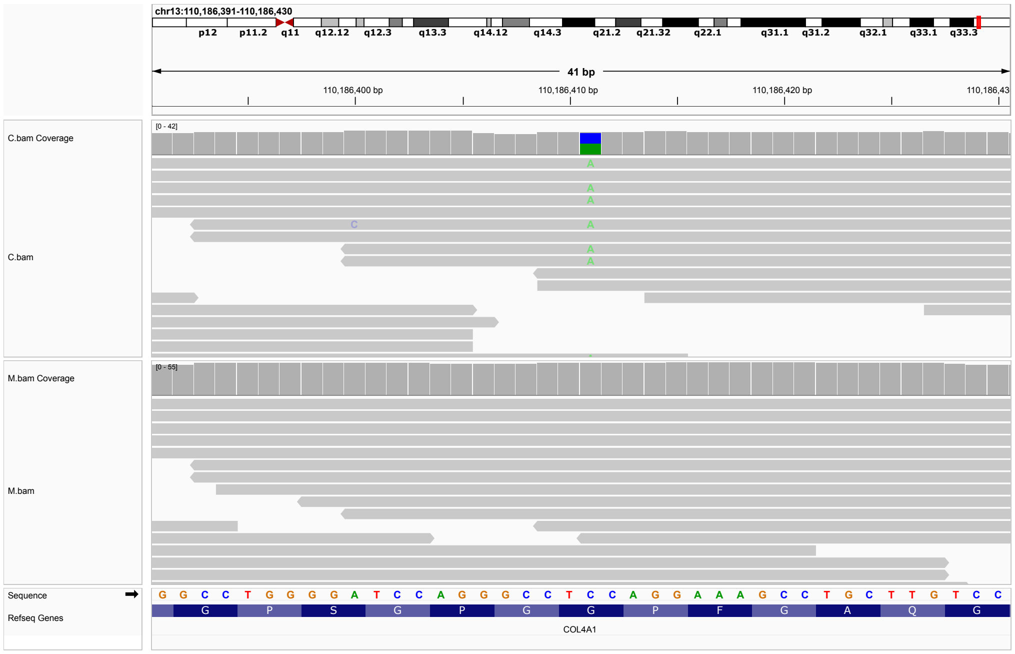


**Figure S1: P1.** Missense variant in *COL4A1*, 13-110186411-C-A, ENST00000375820.10:c.1871G>T:p.(Gly624Val). C: child. M: mother. Paternal sample not available. ACMG evaluation: LP: PM1,PM2,PP2,PP3. PM2 and PP3 were applied because most of the pathogenic variants reported in COL4A1-related disorders affect highly conserved glycine residues within the collagenous domain of the protein [1].

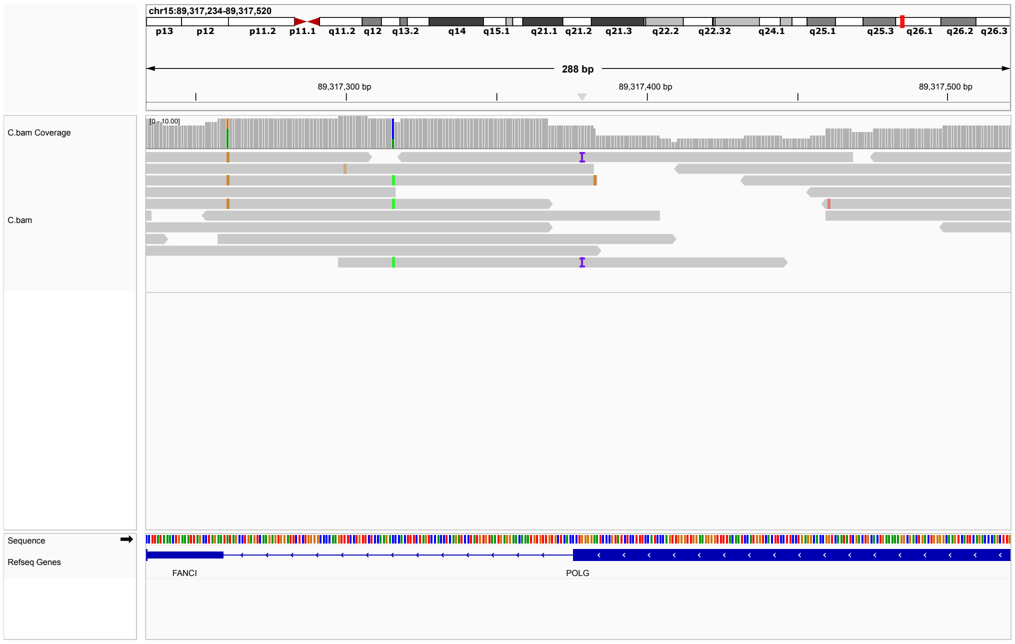


**Figure S2: P2.** Single-nucleotide insertion in *POLG*, 15-89317378-T-TG, ENST00000268124.11:c.3640dup, p.(Gln1214Profs*3). ACMG classification: LP: PVS1, PM2, PP4. Numerous premature truncation variants in this region of the gene have been reported as LP/P in ClinVar, e.g., p.Gln1214Ter (Variation ID: 488798), p.Glu1225fs (Variation ID: 619449), p.Gly1211fs (Variation ID: 21315), p.Tyr1210Ter (Variation ID: 2677952), p.Tyr1210Ter (Variation ID: 1357431). Low coverage but no indication of PCR duplication. Parental samples not available.

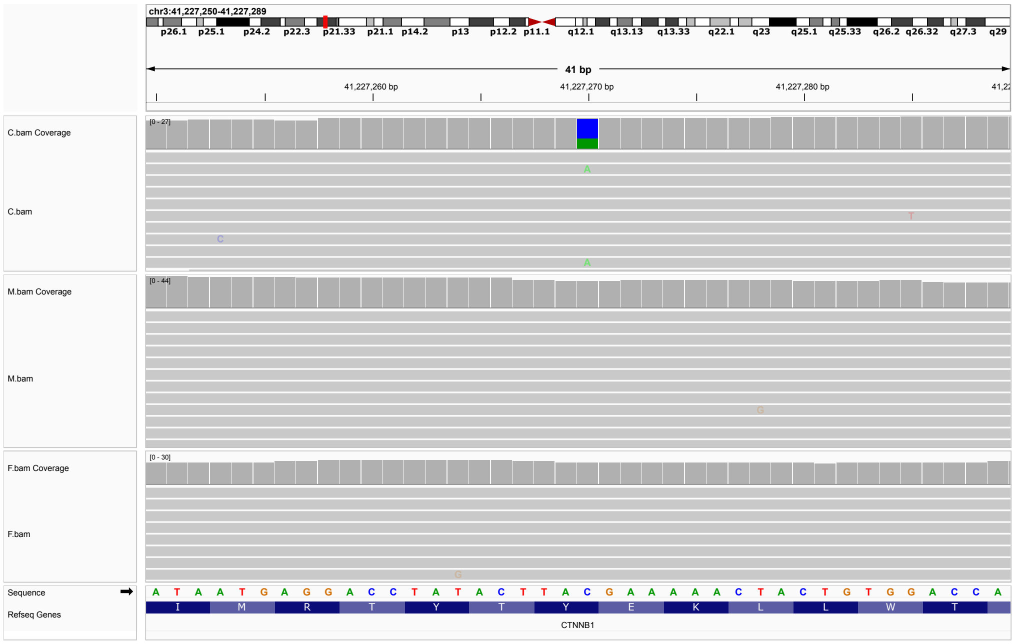


**Figure S3: P3.** Stop-gained variant in *CNNTB1*, 3-41227270-C-A,ENST00000349496.11, c.999C>A, p.(Tyr333*). ACMG classification: P: PVS1, PS2, PM2_Supporting, PP4, PP5_Strong. The variant has been reported as pathogenic four times in ClinVar (Variation ID: 450550). The variant was found as a de novo event.

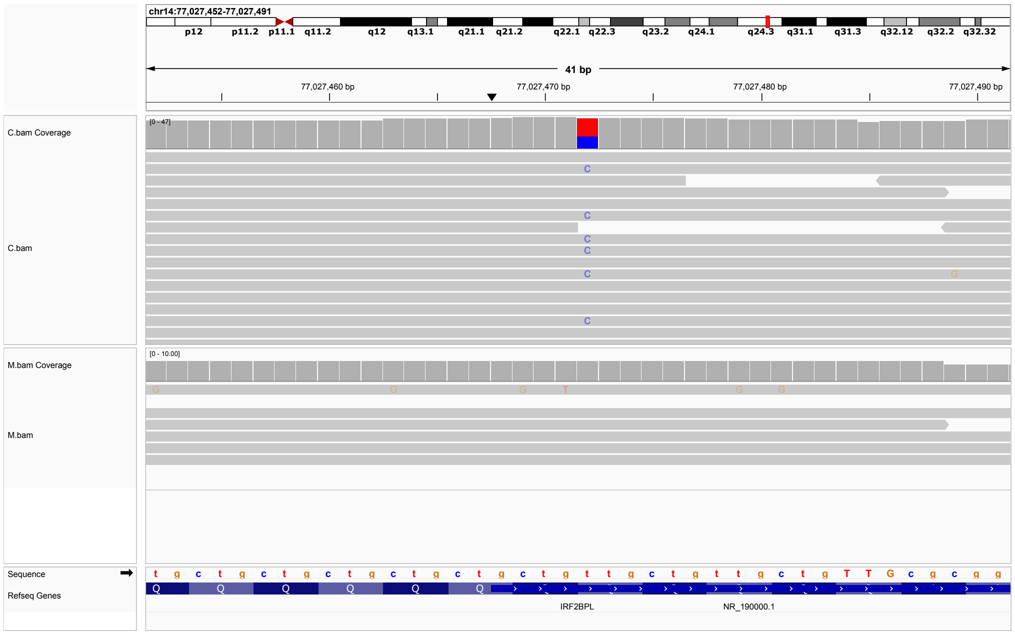


**Figure S4: P4.** Two-nucleotide insertion variant in *IRF2BPL*, 14-77027472-T-TGC, ENST00000238647.5, c.320_321insGC, p.(Gln108Hisfs*45). ACMG classification: LP: PVS1_Strong, PM2_Supporting, PP5. One submission in clinvar classifies the variant as LP (Variation ID: 3251965). Paternal sample not available. An additional heterozygous variant was identified in MATR3 (5-139322030-G-A; ENST00000394805.8,c.1734+1G>A:p.?; rs1581255571; ACMG classification: P: PVS1, PM2_Supporting, PP4. The *MATR3* variant was assessed to be unlikely to be related to the diagnosis of CP because *MATR3* is associated with Amyotrophic lateral sclerosis 21 (OMIM:606070), an adult-onset disease.

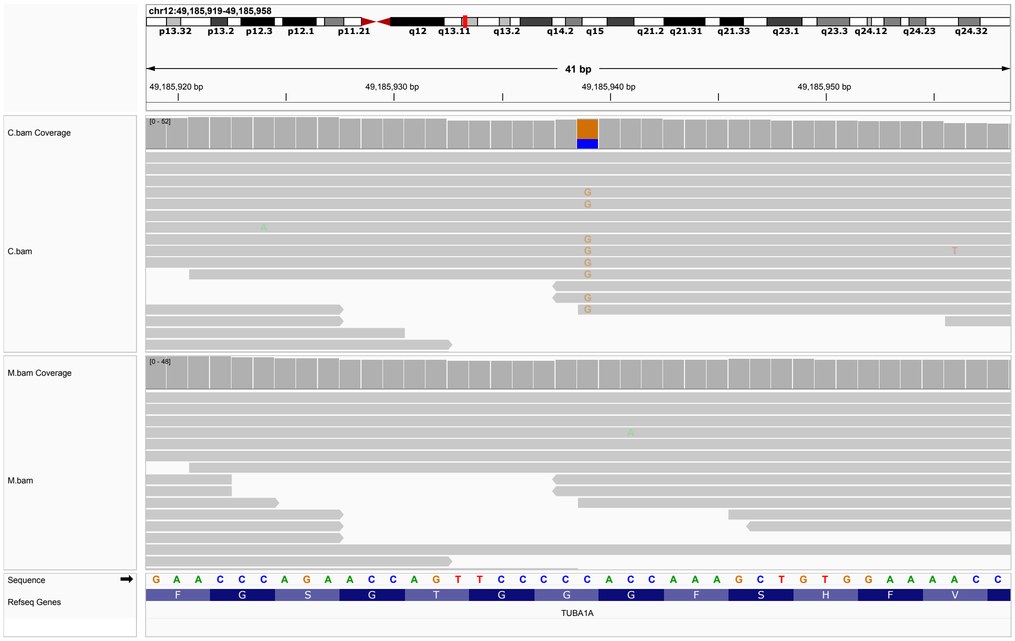


**Figure S5: P5.** Missense variant in *TUBA1A*, 12-49185939-C-G, ENST00000301071.12, c.427G¿C, p.(Gly143Arg). ACMG classification: LP: PM2_Supporting, PP2, PP3_Strong. A single submitter in ClinVar classified the variant as VUS (Variation ID: 1719091). Paternal sample not available.

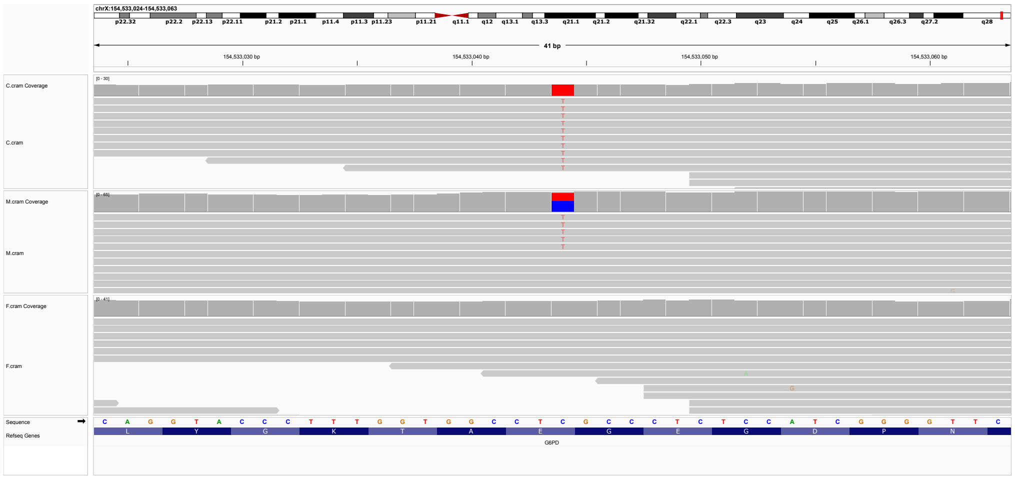


**Figure S6: P6.** Hemizygous missense variant in *G6PD*, X-154533044-C-T, ENST00000393562.10, c.949G¿A, p.(Glu317Lys) ACMG classification: Exomiser VUS (PM1, PP2, PP3, PP5_Strong, BS2), but ClinVar: P/LP (Variation ID: 10401, 19 submissions).

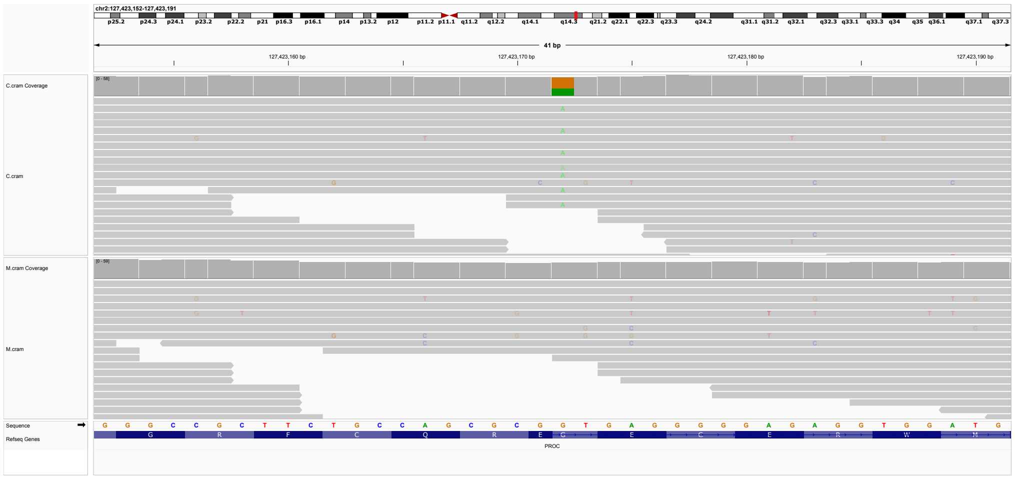

**Figure S7: P7.** Hemizygous missense variant in *PROC*, 2-127423172-G-A,ENST00000234071.8:c.400+1G>A:p.?, ACMG classification: P: PVS1, PS1_Supporting, PM2_Supporting, ClinVar pathogenic (Variation ID: 661205).

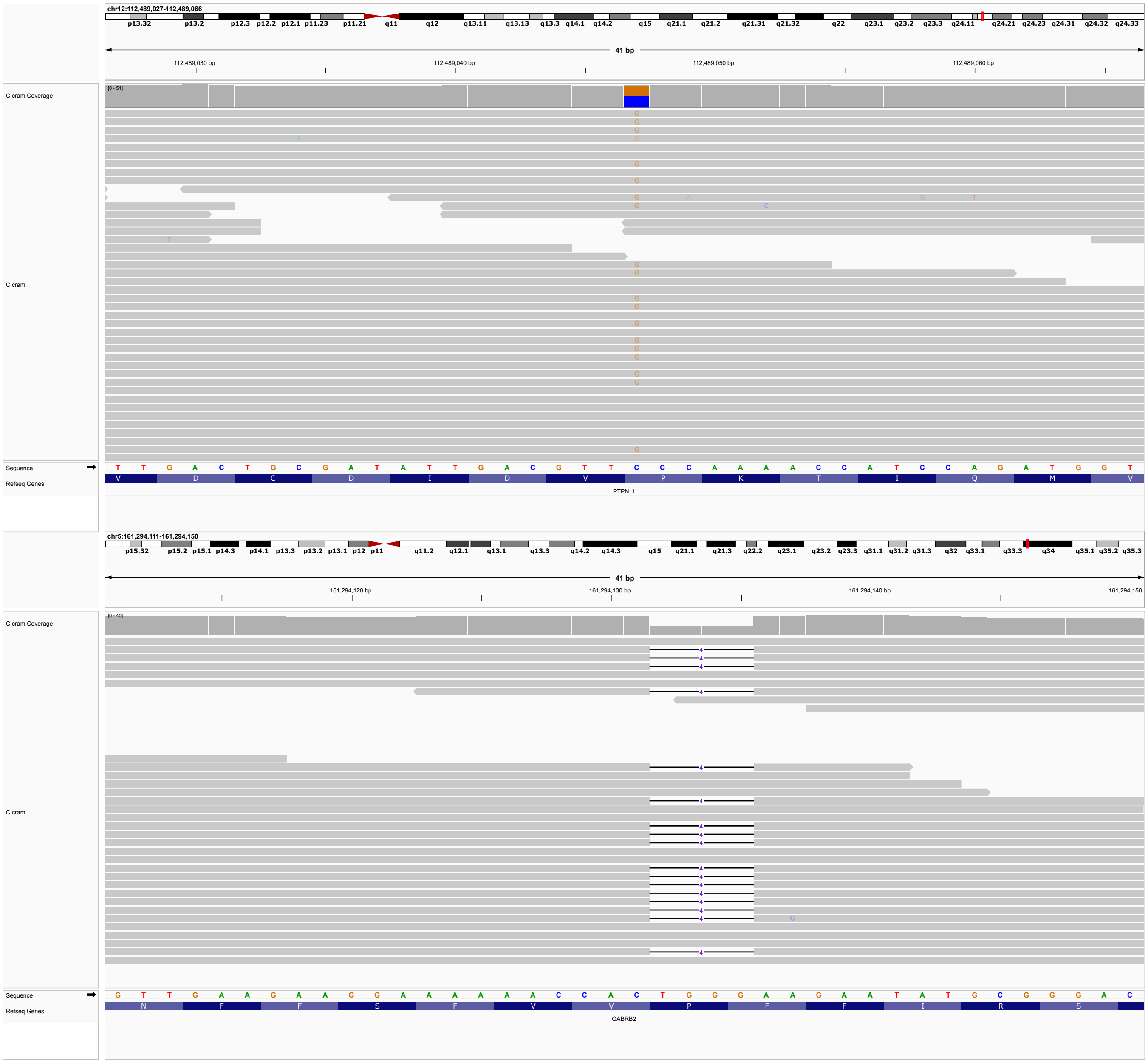


**Figure S8: P8.** This individual was fgound to have two potentially relevant heterozygous P/LP variants. In PTPN11, 12-112489047-C-G, ENST00000351677.7, c.1471C>G, p.(Pro491Ala), ACMG classification: LP, PM1_Supporting, PM5, PP2, PP5_Strong, CLinVar 6 P/LP assessments (Variation ID: 181503); and *GABRB2*, 5-161294131-CTGGG-C, ENST00000393959.6:c.1485_1488del:p.(Phe495Leufs*16), ACMG classification LP: PVS1_Strong, PM2_Supporting, PP4, no ClinVar entry. Parental samples were not available.

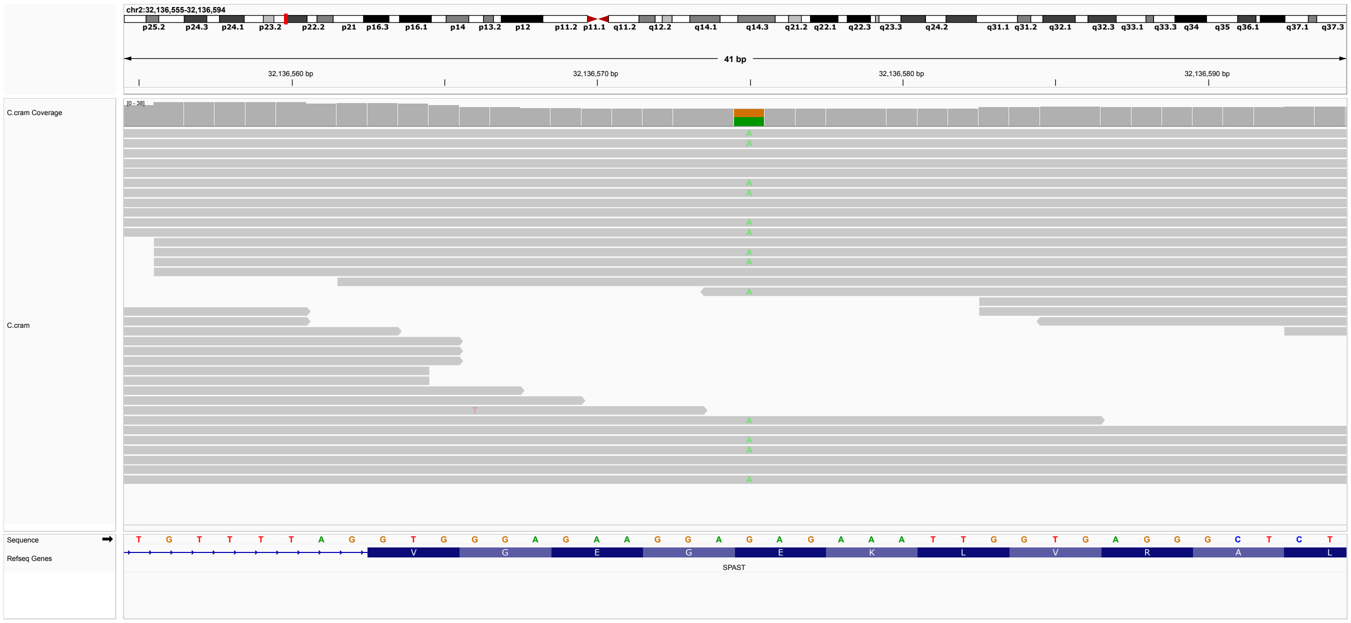

**Figure S9: P9.** Heterozygous missense variant in SPAST, 2-32136575-G-A, ENST00000315285.9, c.1258G¿A, p.(Glu420Lys). ACMG classification: P: PM1,PM2,PM5,PP3,PS1. Parental samples not available. Three other variants at the same position are listed in ClinVar: p.Glu420Asp (LP, Variation ID: 1348218), p.Glu420Gly (P, Variation ID: 1686225), p.Glu420Gln (P, Variation ID:576595).

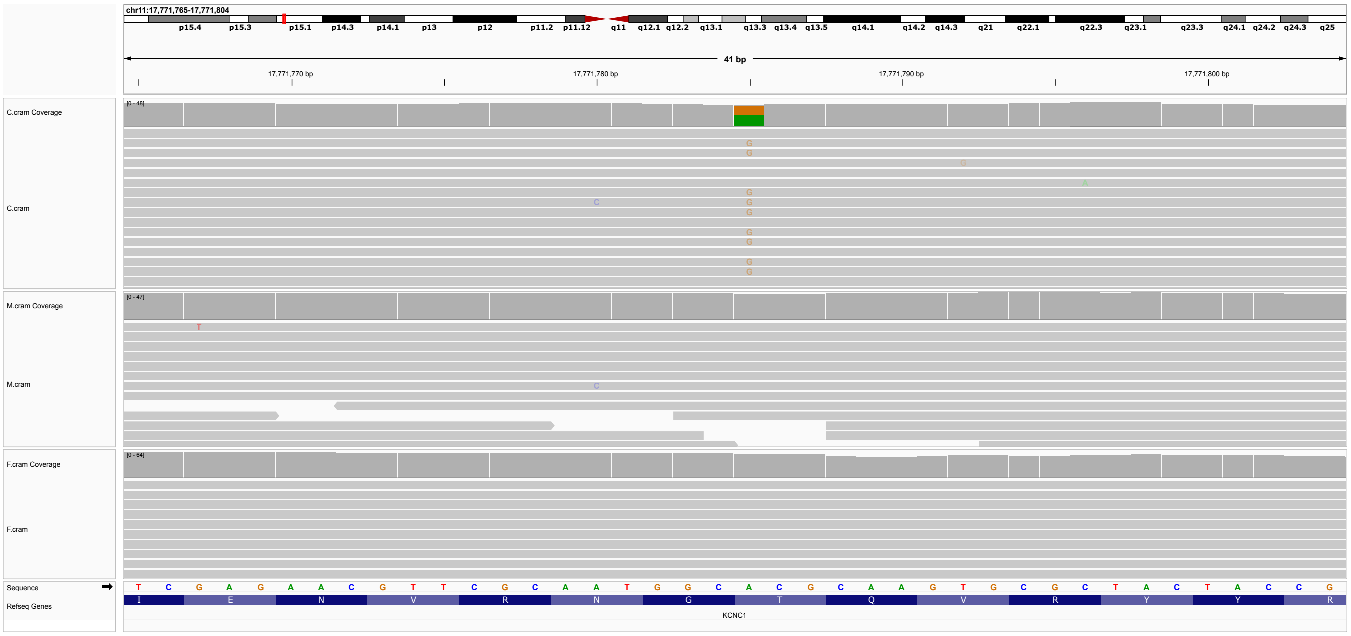


**Figure S10: P10.** De novo heerozygous missense variant in KCNC1, 11-17771785-A-G, ENST00000265969.8, c.691A¿G, p.(Thr231Ala). ACMG classification LP: PS2, PM2_Supporting, PP2, PP5. ClinVar LP (Variation ID: 568146). KCNC1 variants are associated with Epilepsy, progressive myoclonic 7 (OMIM:616187).

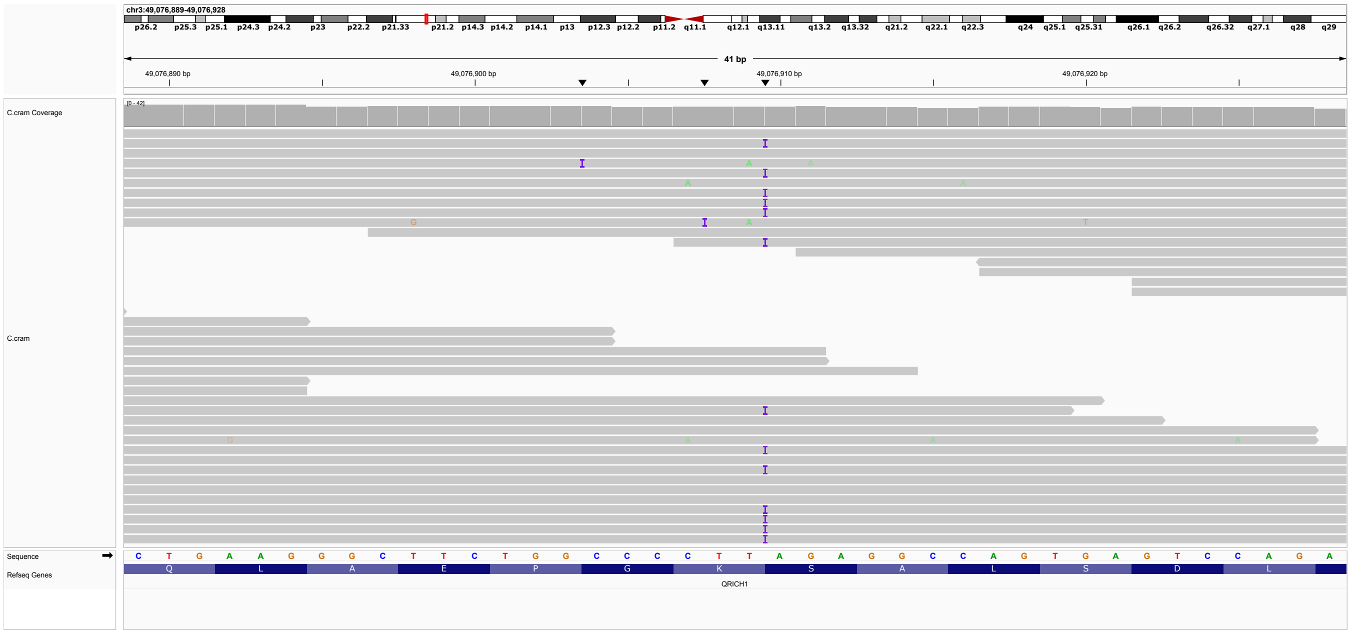


**Figure S11: P11.** One-nucleotide duplication in QRICH1, 3-49076909-T-TA, ENST00000395443.7,c.108dup,p.(Lys37*). ACMG classification LP: PVS1, PM2_Supporting.

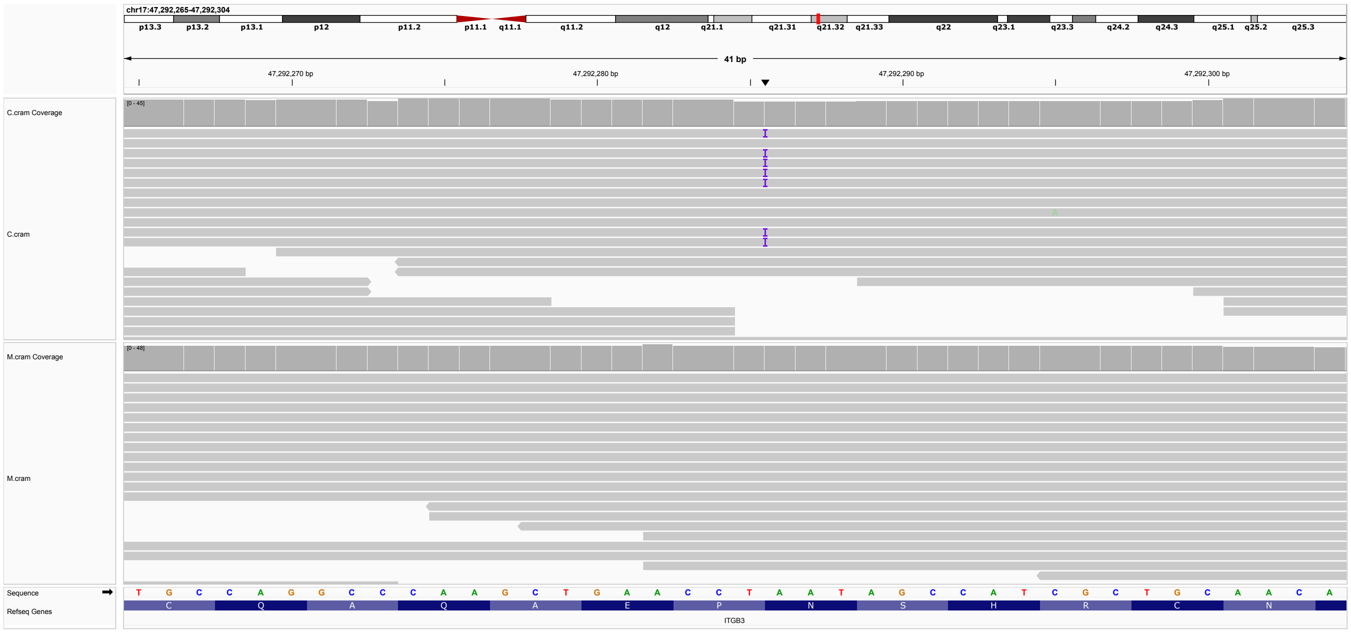


**Figure S12: P12.** One-nucleotide duplication in ITGB3, 17-47292285-T-TA, ENST00000559488.7, c.1409dup, p.(Asn470Lysfs*2). ACMG classification P: PVS1, PP5_VeryStrong.

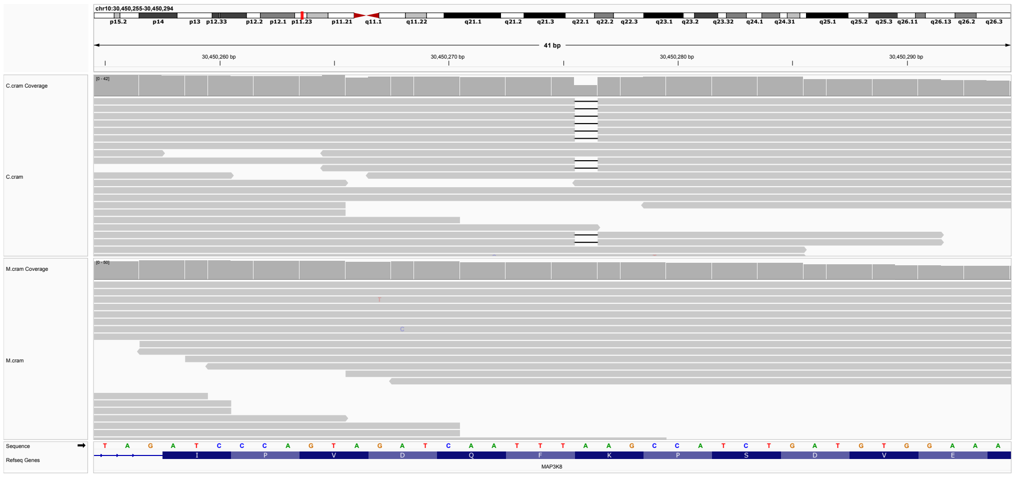


**Figure S13: P13.** Heterozygous single-nucleotide deletion in *MAP3K8*, 10-30450275-TA-T, ENST00000263056.6, c.524del, p.(Lys175Serfs*51). ACMG classification LP: PVS1, PM2. Somatic variants in MAP3K8 can be associated with lung cancer (OMIM:211980) [2]. No disease associated with germline-variants in this gene has been reported, but alterations in transcriptional dysregulation of trophic pathways including mitogen-activated protein kinase (MAPK) signalling pathway have been demonstrated in patient-derived cell lines [3]. Paternal sample not available.

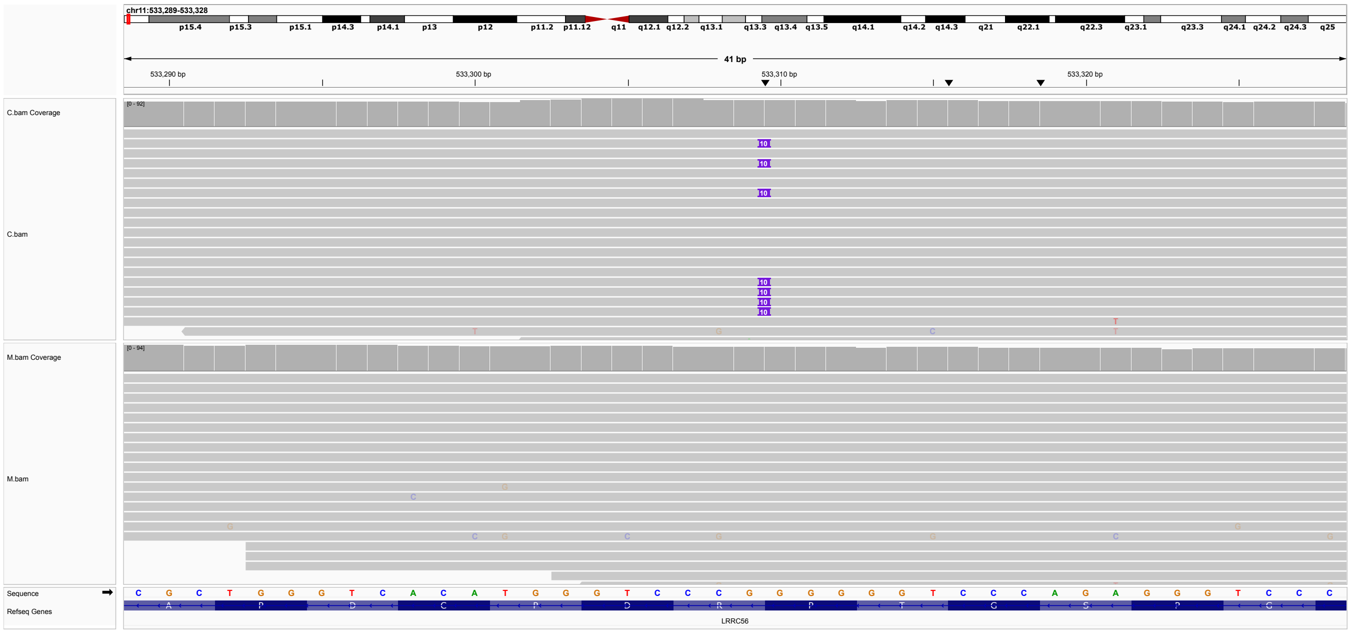

**Figure S14: P14.** Heterozygous insertion in HRAS, 11-533309-G-GGGGGGTCCCA, HRAS:ENST00000417302.7:c.490_499dup:p.(Pro167Leufs*36). ACMG classification LP: PVS1, PM2. Most published pathogenic variants in HRAS are missense. An additional nearby frameshoft variant is registered in ClinVar, NM_176795.5(HRAS):c.488_497del (p.Leu163fs) (Variation ID: 1321186), associated with Costello syndrome (Accession: SCV002014563.2 and SCV003835072.1) and without specified diagnosis (Accession: SCV005690321.1). ENST00000417302.7 corresponds to NM_176795.5 (MANE Plus Clinical). Paternal sample not available.

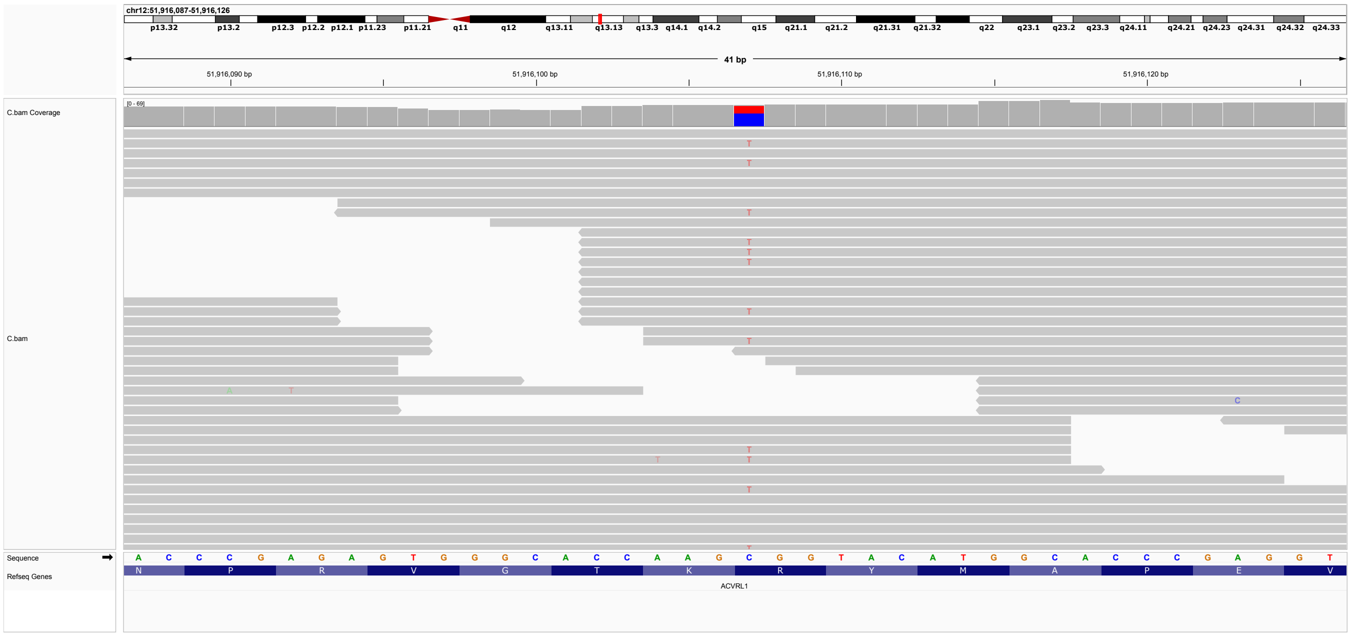


**Figure S15: P15.** Heterozygous missense variant in *ACVRL1*, 12-51916107-C-T, ENST00000388922.9, c.1120C¿T, p.(Arg374Trp). ACMG classification LP: PM1, PM5, PP3, PP5. ClinVar P/LP (Variation ID:8249, 11 submissions). Parental samples not available.

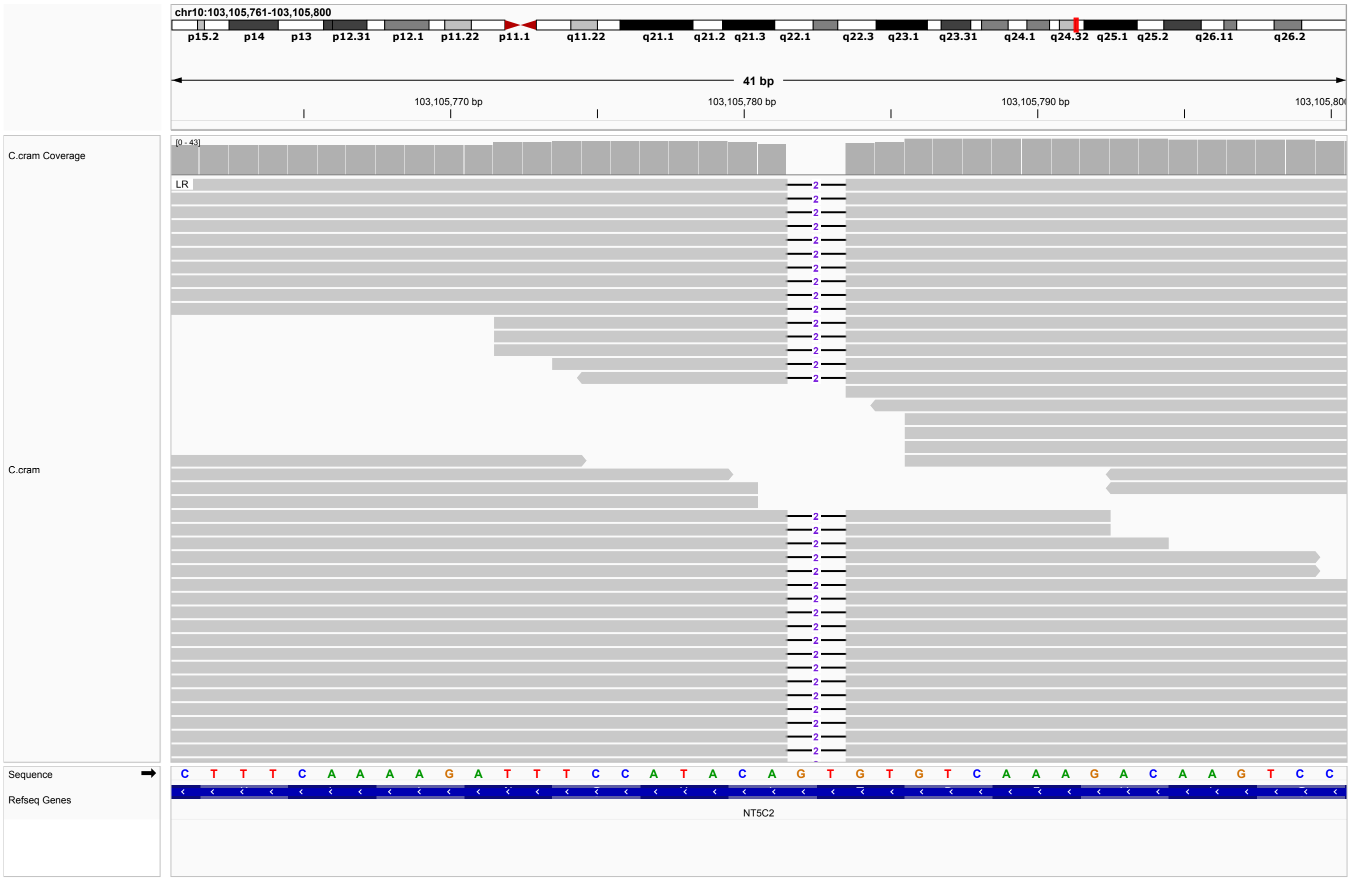


**Figure S16: P16.** Homozygous two-nucleotide deletion in NT5C2, ENST00000404739.8, c.312_313del, p.(Leu105Valfs*21). ACMG classification P: PVS1,PM2,PP3, ClinVar P/LP (Variation ID: 541762, three submissions). Variants in NT5C2 are associated with Spastic paraplegia 45, autosomal recessive (OMIM:613162). Other premature truncation codon variants have been identified in this disorder. Parental samples not available.

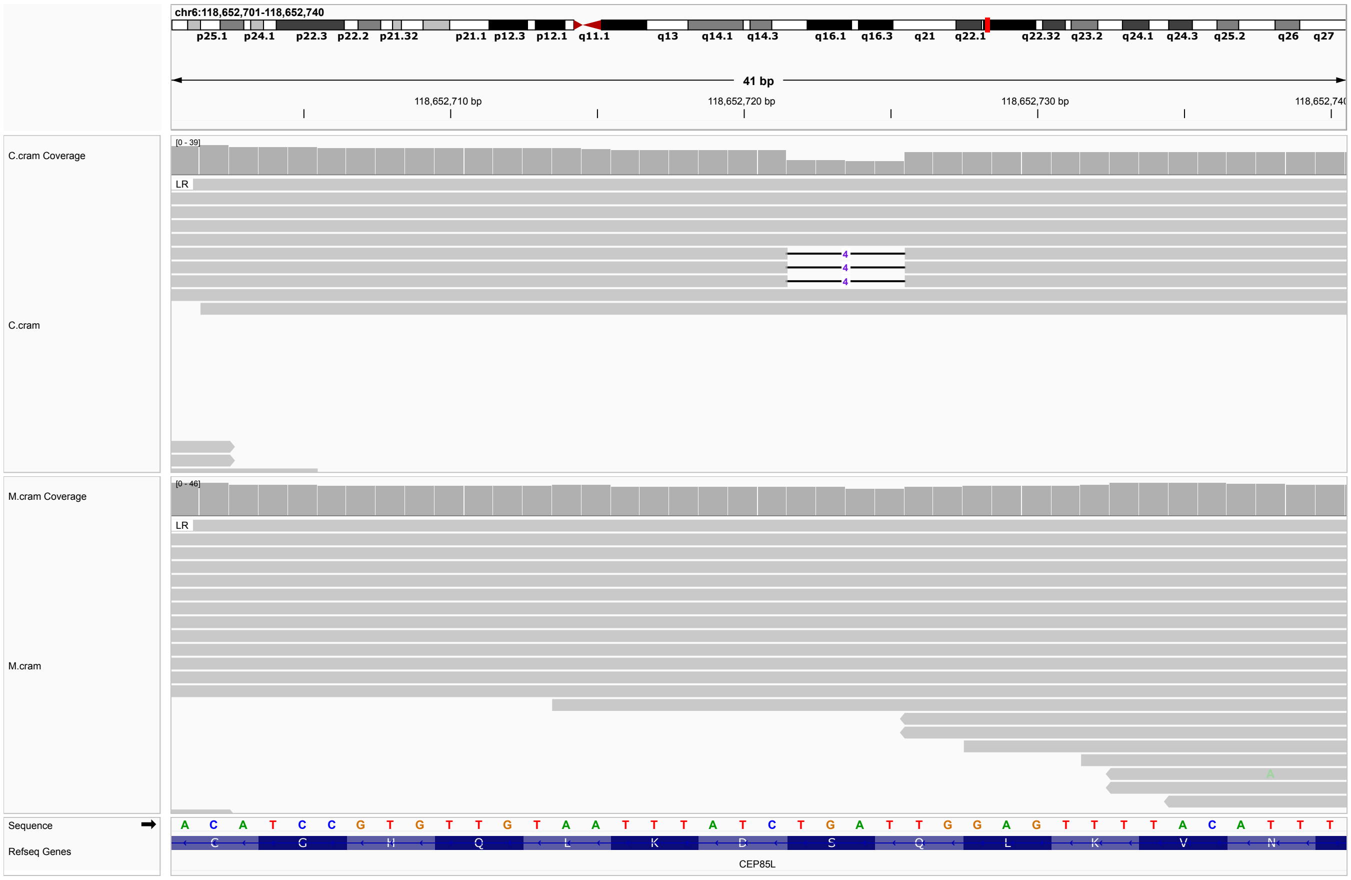


**Figure S17: P17.** 6-118652721-CTGAT-C, NM_001178035.2(CEP85L):c.57_60del (p.Ser20fs). ACMG classification P: PVS1,PP5,PP3. ClinVar LP (Variation ID: 3027447, one submission) Heterozygous CEP85L variants can cause lissencephaly-10 (OMIM:618873).

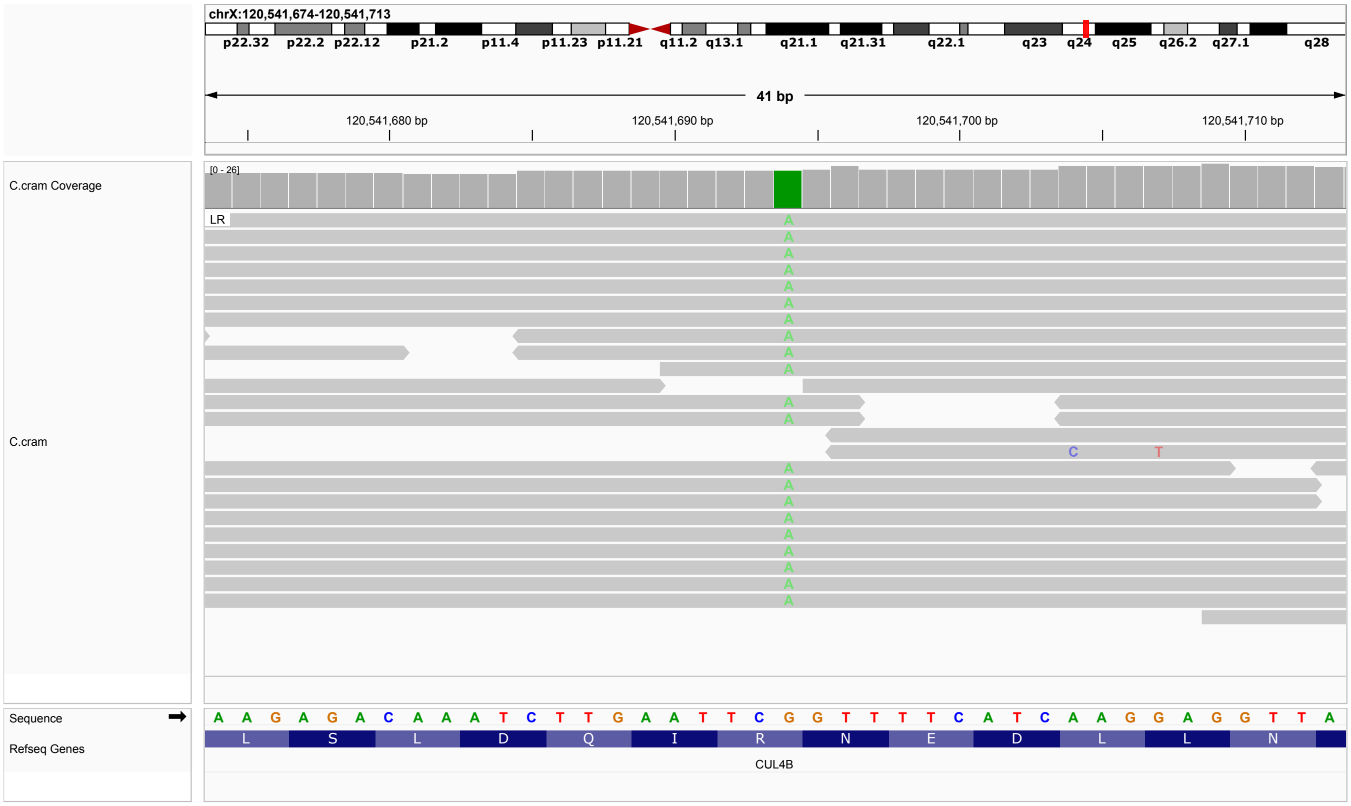


**Figure S18: P18.** Hemizygous top-gained variant in *CUL4B*, X-120541694-G-A, ENST00000371322.11, c.1351C¿T, p.(Arg451*). ACMG classification P: PM2,PP5,PVS1. One submission in ClinVar interprets the variant as P (Variation ID: 620257).

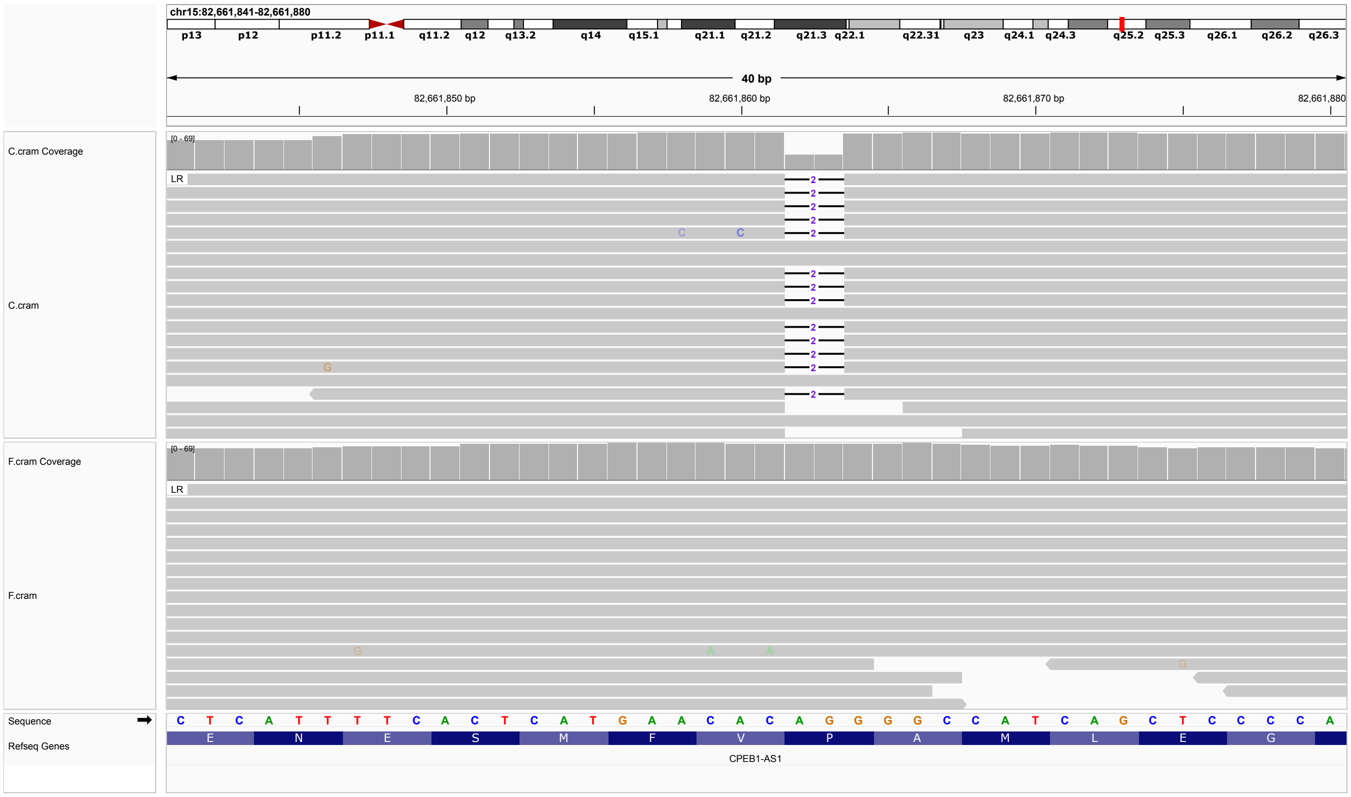


**Figure S19: P19.** Heterozygous deletion in *AP3B2*, 15-82661861-CAG-C, ENST00000535359.6, c.2978_2979del, p.(Pro993Argfs*5). ACMG classification P: PVS1, PP4, PP5. ClinVar: P/LP (Variation ID: 987398, 3 submissions).

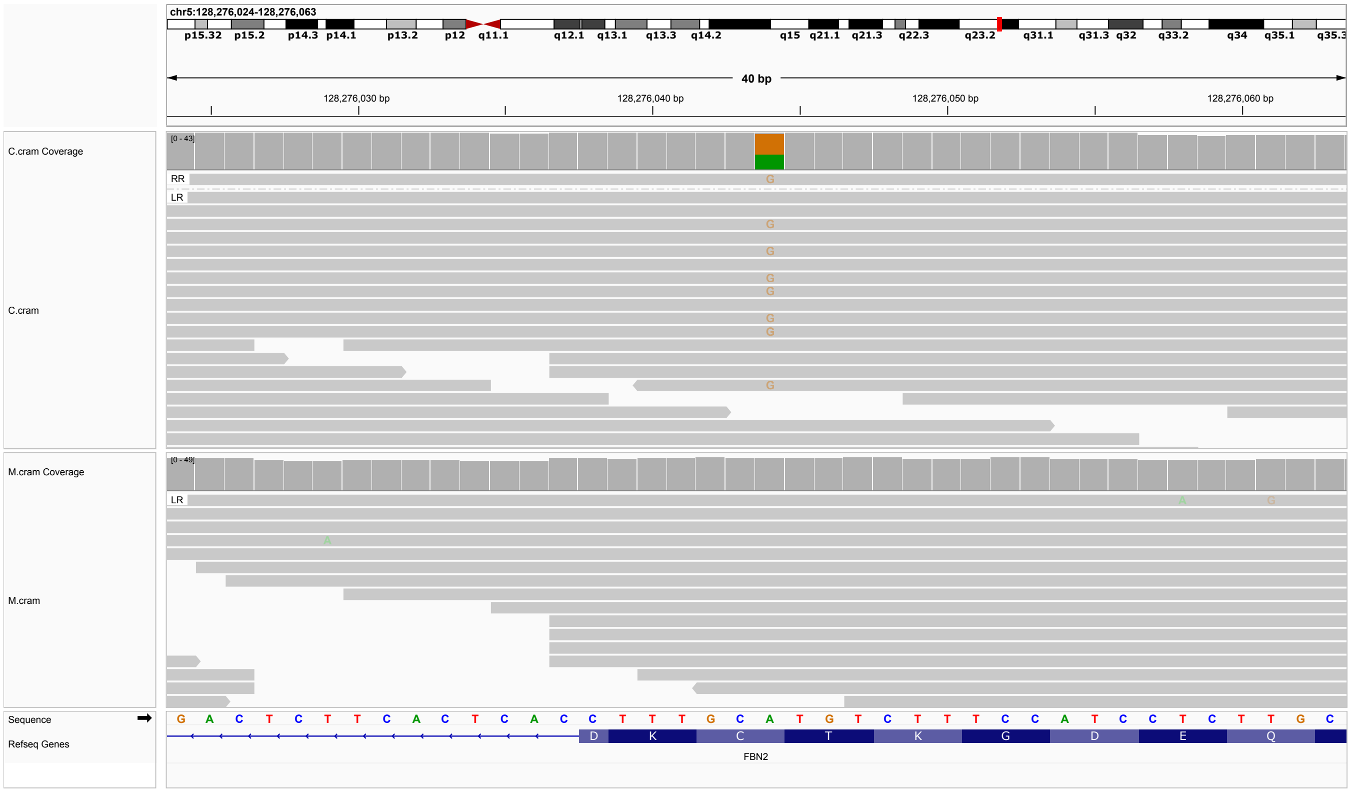


**Figure S20: P20.** Heterozygous missense variant in *FBN2*, 5-128276044-A-G, ENST00000262464.9, c.7588T¿C,p.(Cys2530Arg). ACMG classification LP: PM2,PP3,PP4,PM1. PM1 was assigned because variants affecting one of the six cysteine residues of the cbEGF modules of fibrillin-2 are a common cause of congenital contratural arachnodactyly [4]. The one available ClinVar entry interprets the variant as VUS (Variation ID: 2898900).

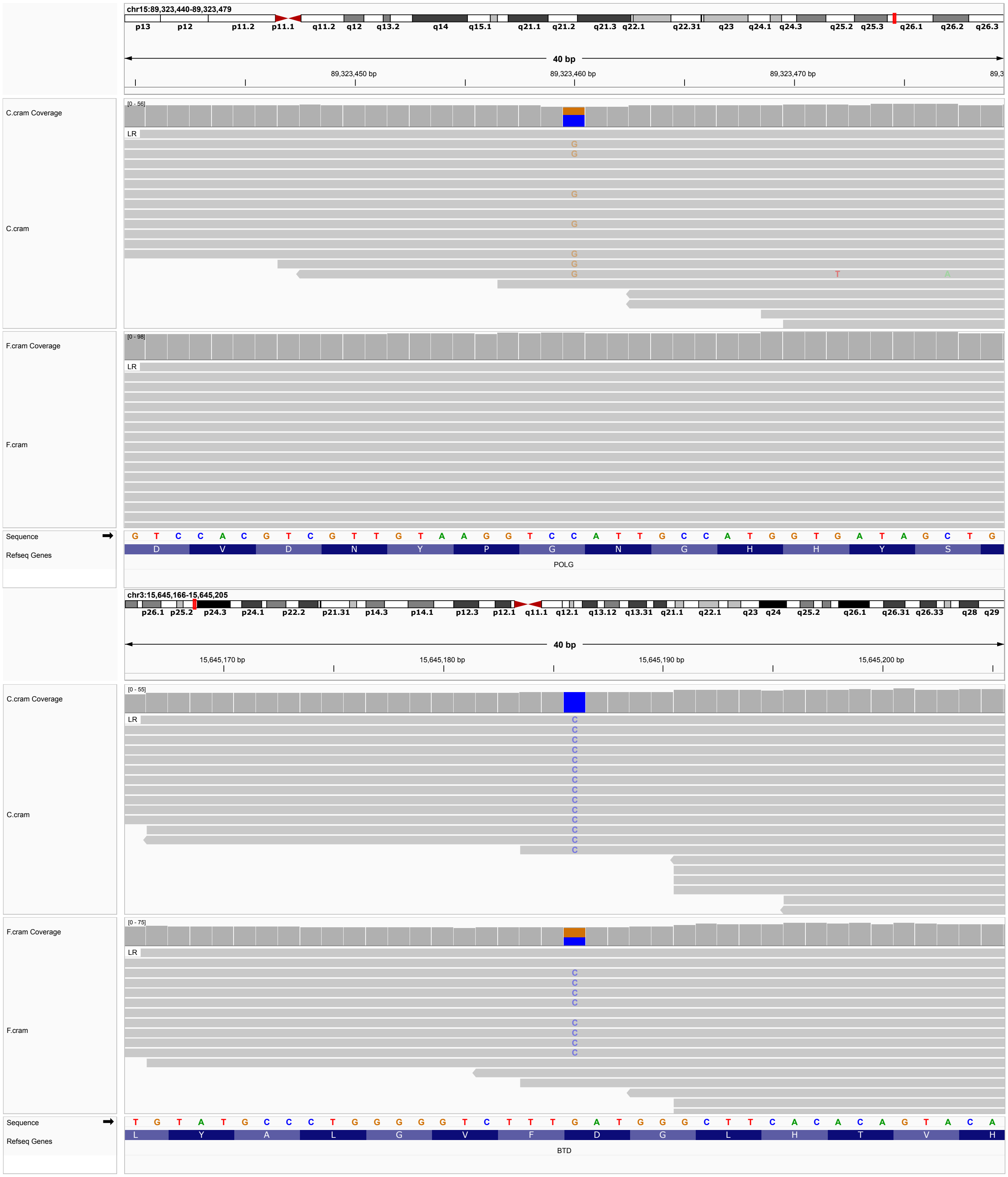


**Figure S21: P21.** Heterozygous missense variant in *POLG*, 15-89323460-C-G, ENST00000268124.11, c.2209G>C, p.(Gly737Arg). ACMG classification LP: PP3, PP4, PP5. ClinVar: P/LP (Variation ID: 13513, 29 submissions). Additionally, a homozygous missense variant in *BTD*, 3-15645186-G-C, ENST00000643237.3, c.1270G¿C, p.(Asp424His) was identified. ACMG classification: LP: PM1,PM5,PP3,PP5. ClinVar: Path (36 submission), LP (3 submissions), VUS (1 submission), Variation ID: 1900. Maternal sample not available.

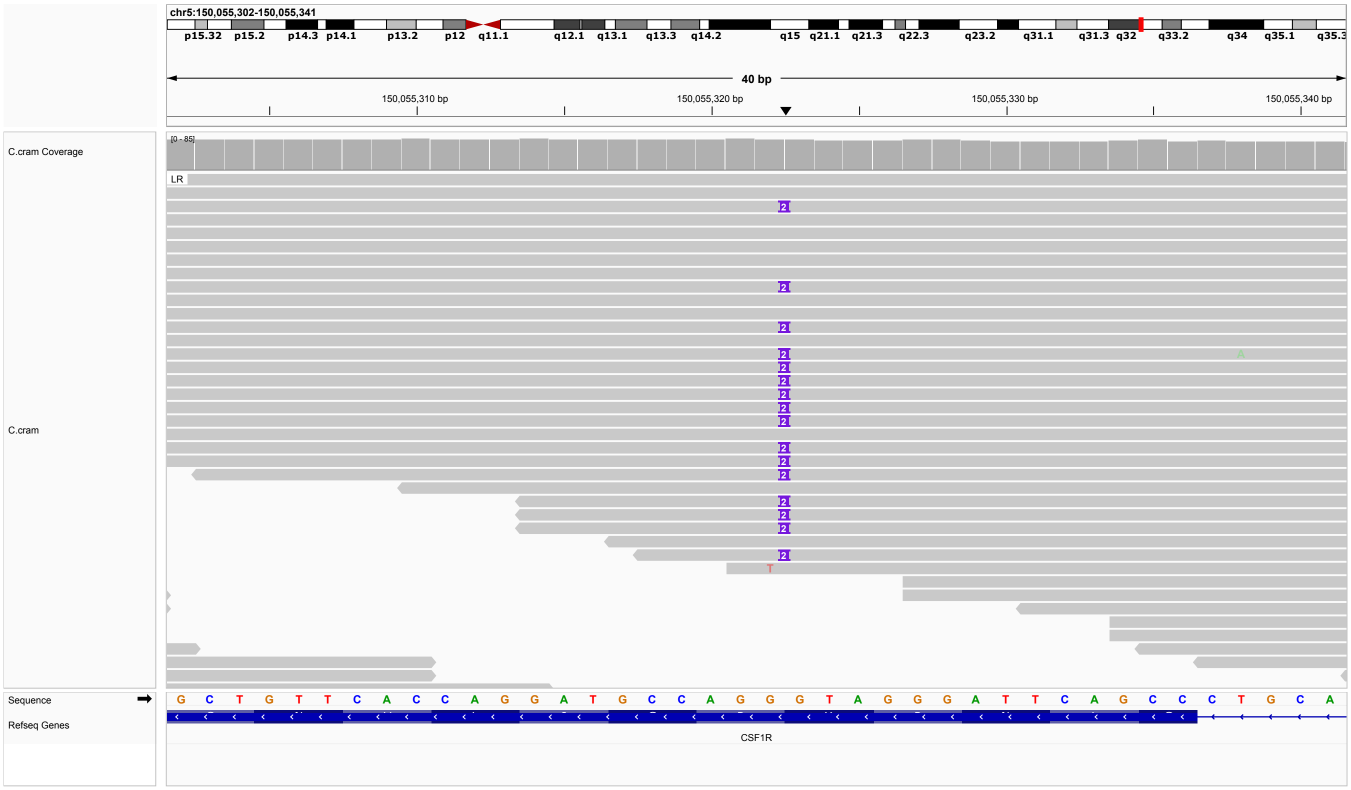


**Figure S22: P22.** Heterozygous duplication in CSF1R, 5-150055322-G-GGT, ENST00000675795.1, c.2567_2568dup, p.(Pro857Thrfs*6). ACMG classification P: PVS1, PM2, PP4. Heterozygous pathogenic CSF1R variants are associated with Leukoencephalopathy, diffuse hereditary, with spheroids 1 (OMIM:221820). Paternal sample not available.

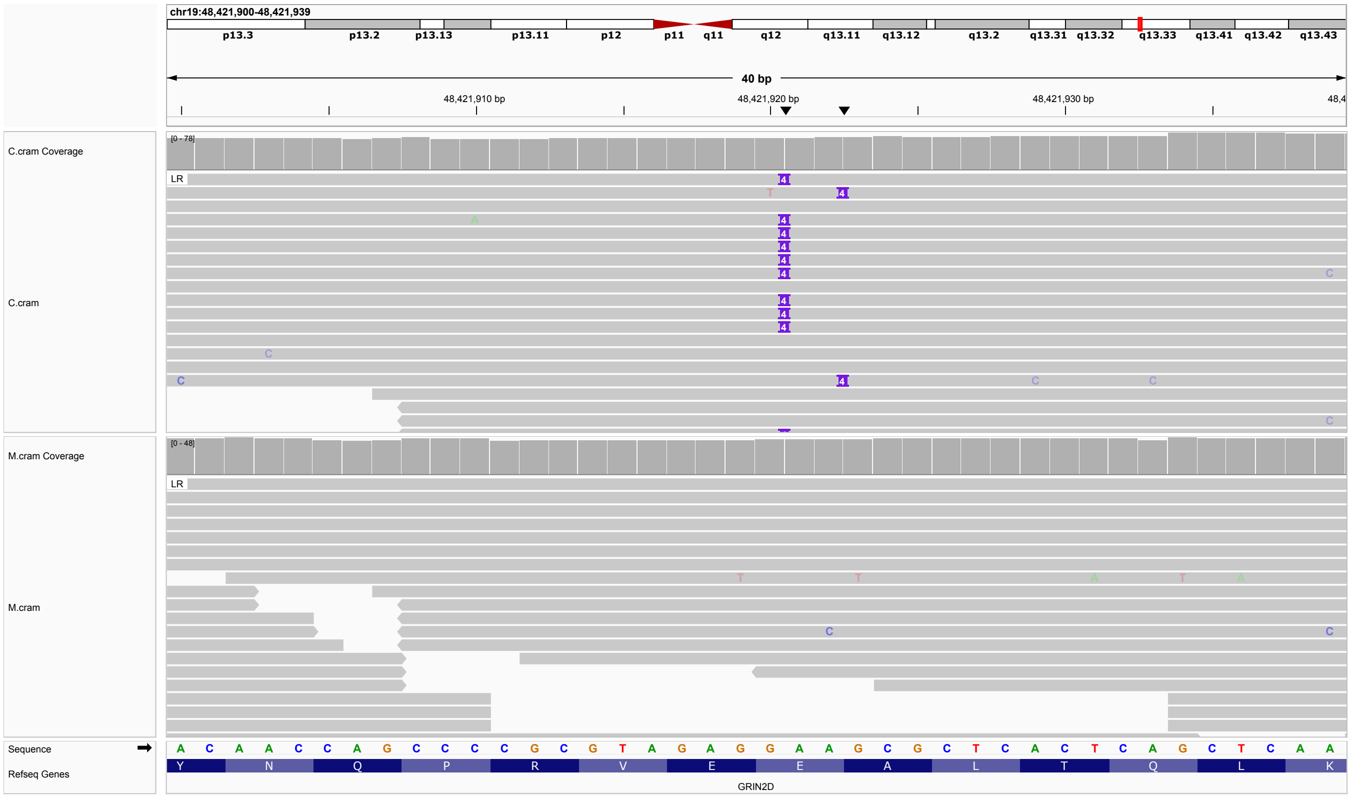


**Figure S23: P23.** Heterozygous 4-nucleotide duplication in *GRIN2D*, 19-48421920-G-GAAGC, ENST00000263269.4, c.2228_2231dup, p.(Leu745Serfs*71). ACMG classification P: PVS1, PM2, PP4. Developmental and epileptic encephalopathy-46 (DEE46; OMIM:617162) is caused by heterozygous mutation in the GRIN2D gene.

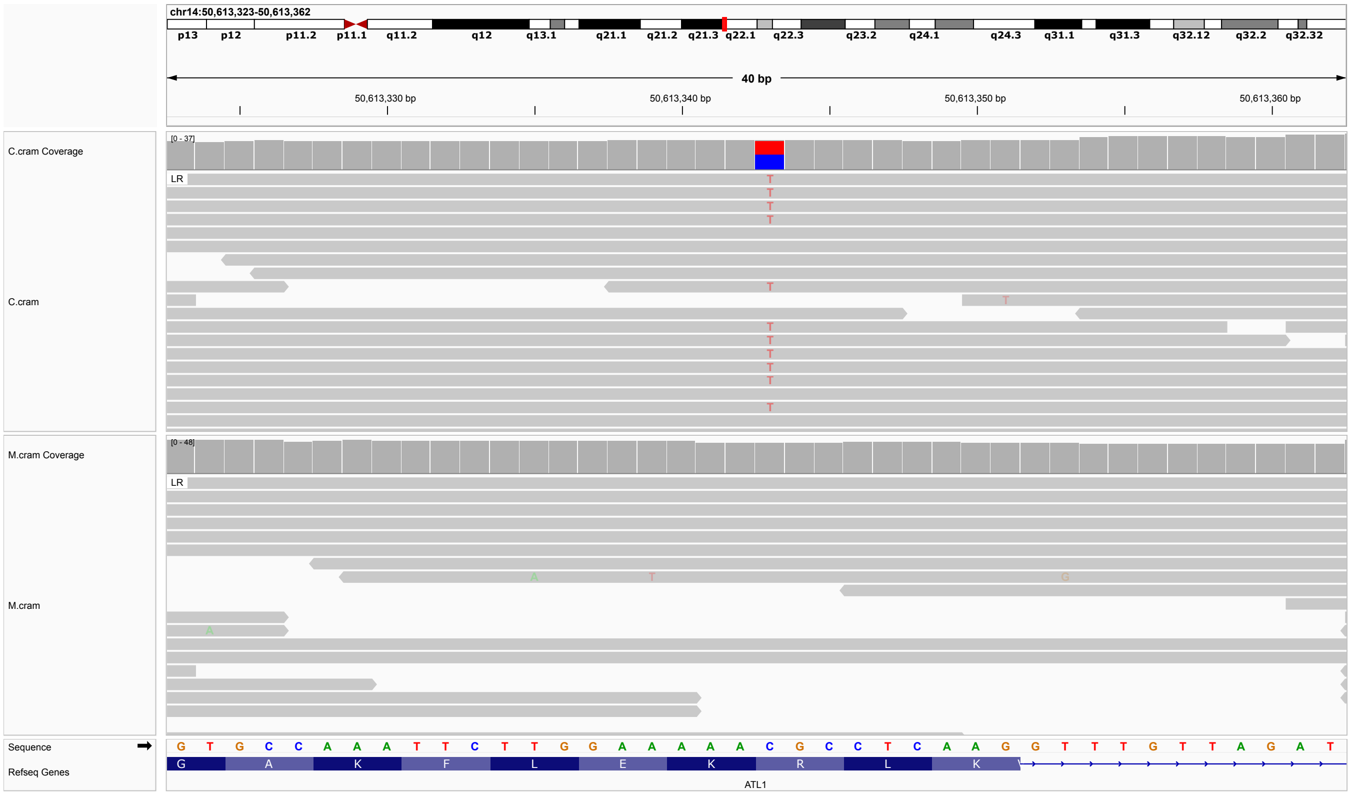


**Figure S24: P24.** Heterozygous missense variant in *ATL1*, 14-50613343-C-T, ENST00000358385.12, c.715C¿T, p.(Arg239Cys). ACMG classification LP: PM5, PP2, PP4, PP5. Listed in ClinVar as P/LP (17 submissions, Variation ID: 4346).

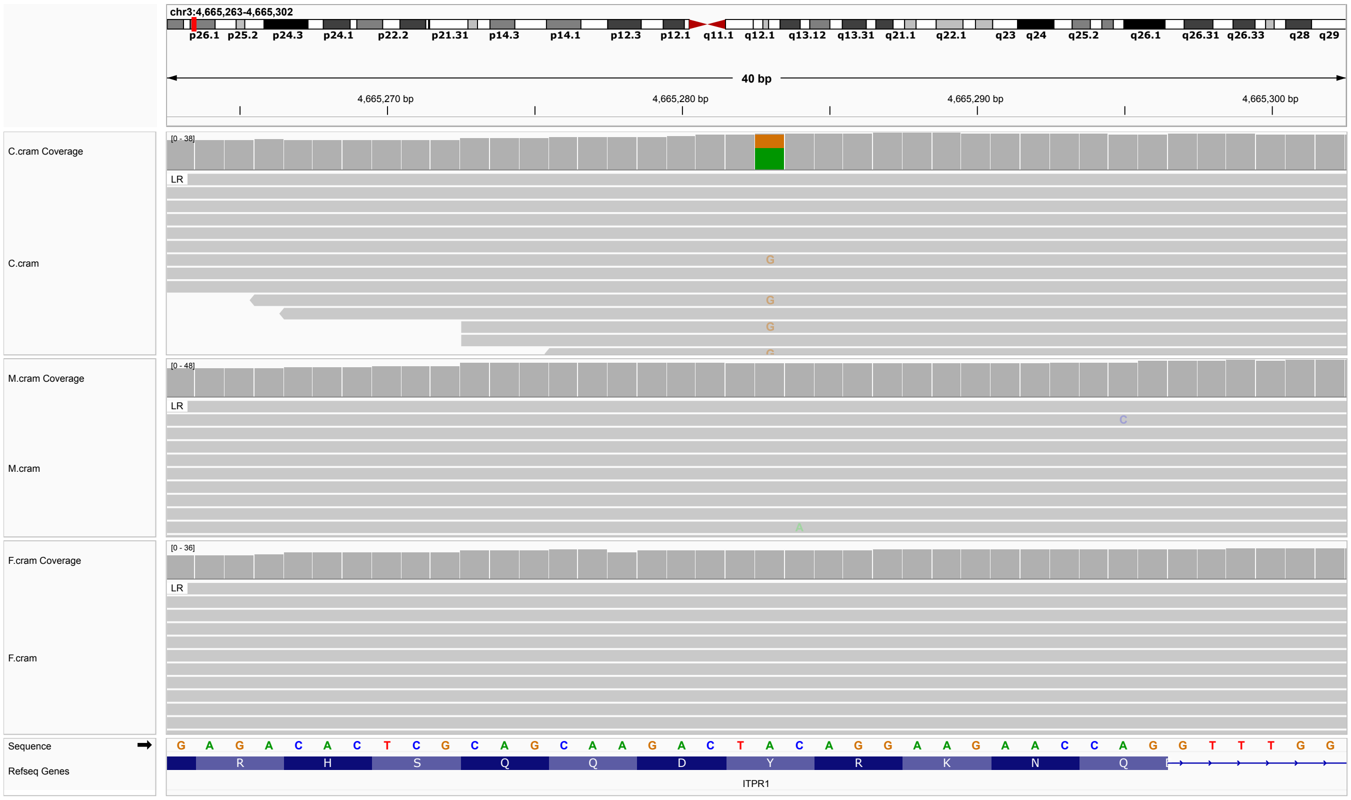


**Figure S25: P25.** De novo heterozygous missense variant in *ITPR1*, 3-4665283-A-G, ENST00000649015.2, c.1700A¿G, p.(Tyr567Cys). ACMG classification P: PS2, PM1, PM2, PM5, PP3, PP4, PP5. The variant is listed in ClinVar as pathogenic (Variation ID: 2579948, two submissions).

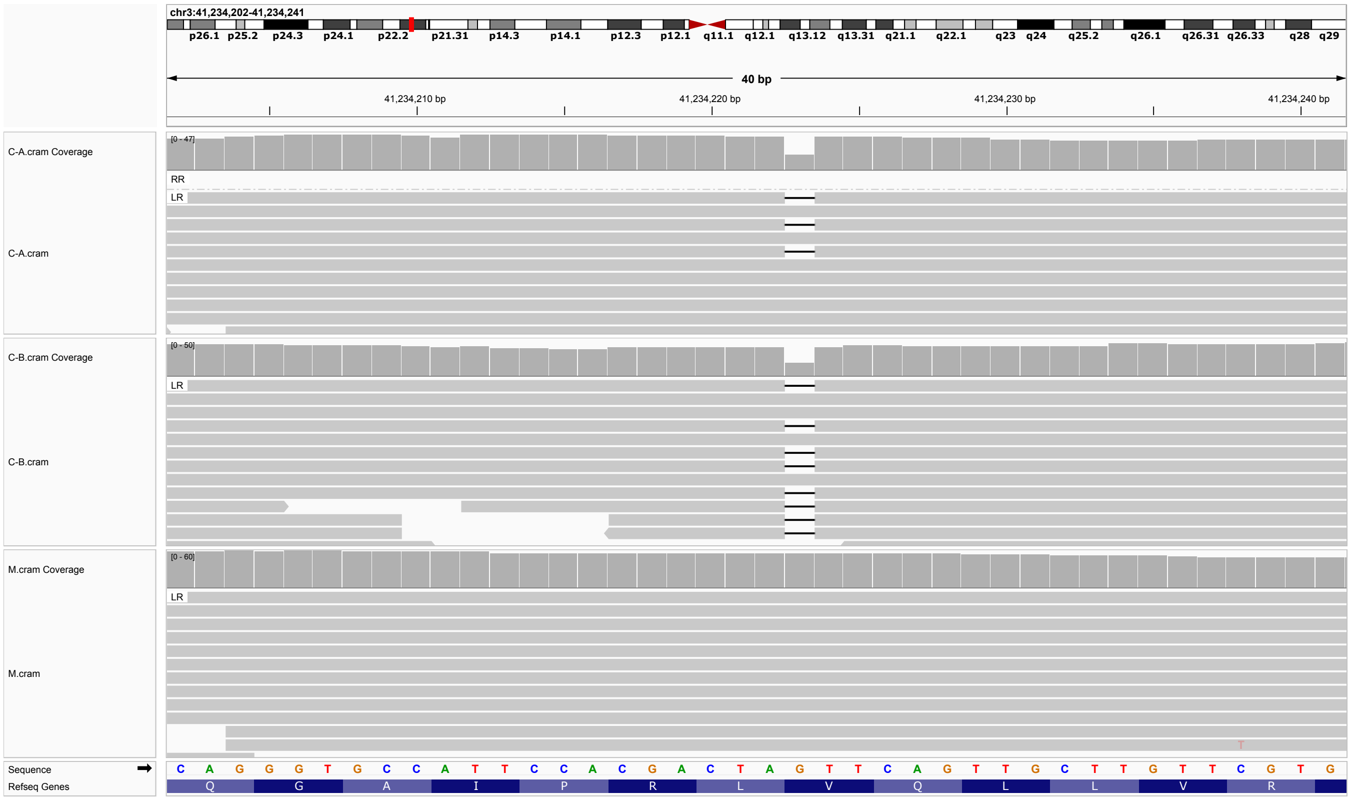


**Figure S26: P26.** Heterozygous one-nucleotide deletion in *CTNNB1*, 3-41234222-AG-A, ENST00000349496.11, c.1609del, p.(Val537Phefs*33) ACMG classification P: PVS1, PM2, PP4. Two affected children (C-A and C-B). Paternal sample not available. Both parents are clinically unaffected, and thus a germline mosaic appears possible.

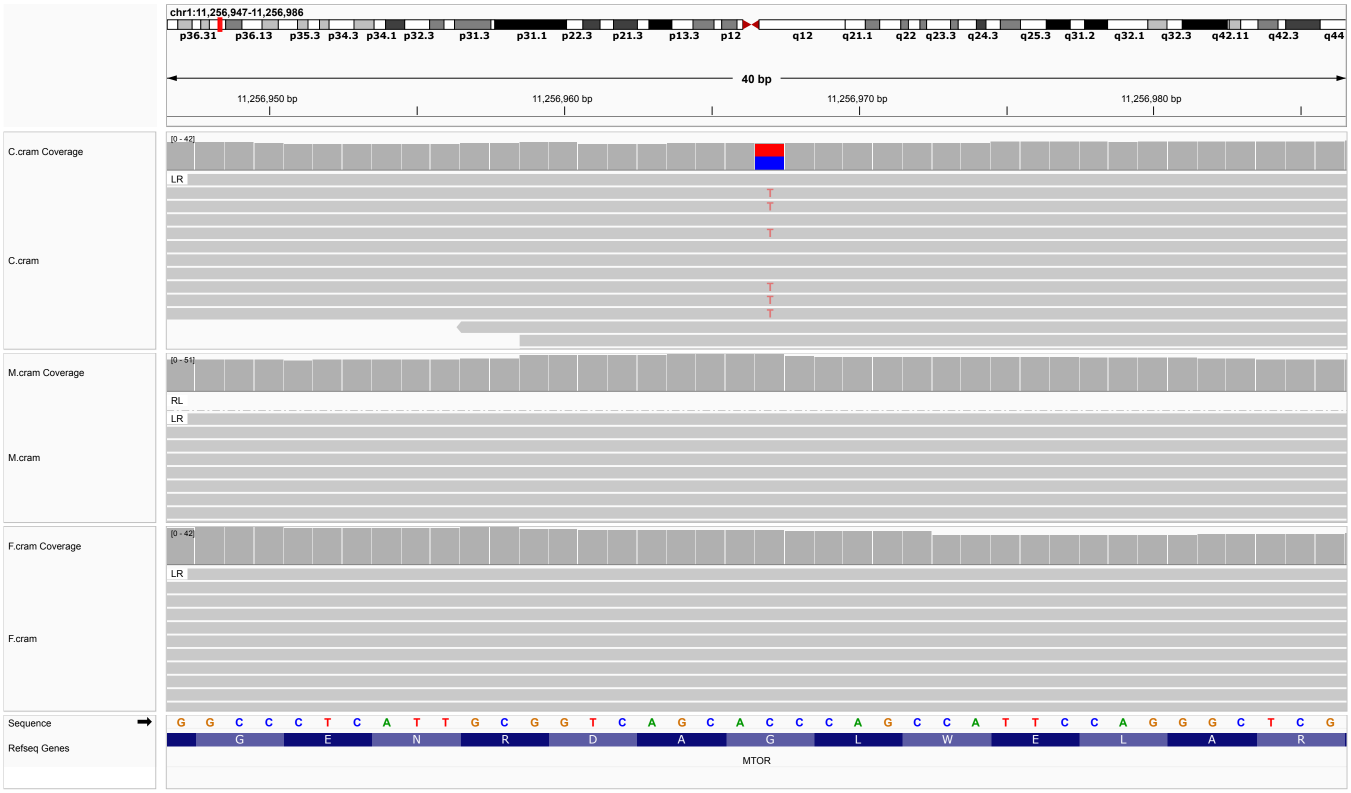


**Figure S27: P27.** De novo hetterozygous missense variant in *MTOR*, 1-11256967-C-T, ENST00000361445.9, c.470G>A, p.(Gly157Asp). ACMG classification LP:PS2, PM2, PP4.

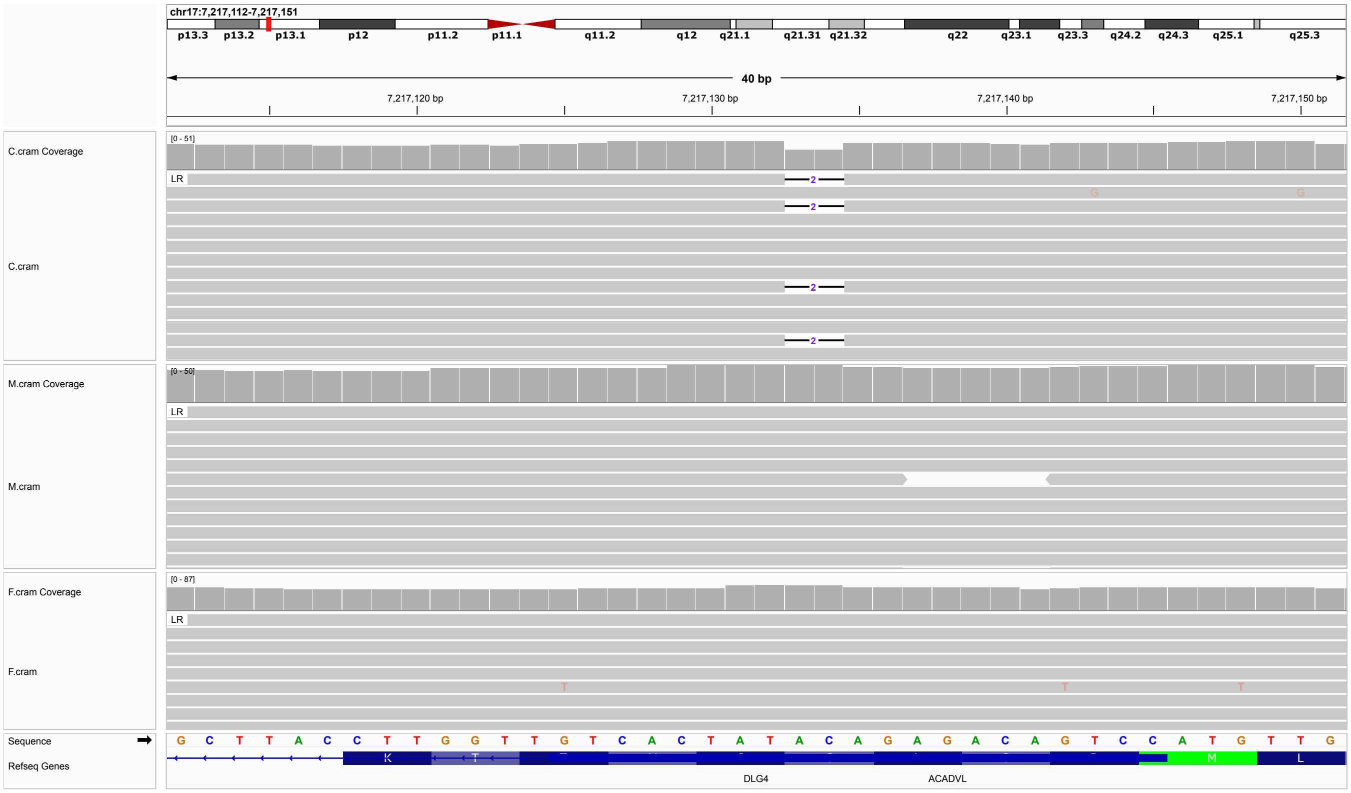


**Figure S28: P28.** De novo two-nucleotide deletion in *DLG4*, 17-7217132-TAC-T, ENST00000399506.9, c.14_15del, p.(Cys5Tyrfs*12). ACMG classification P: PVS1, PS2, PM2. Autosomal dominant intellectual developmental disorder-62 (OMIM:618793) is caused by heterozygous mutation in the DLG4 gene.

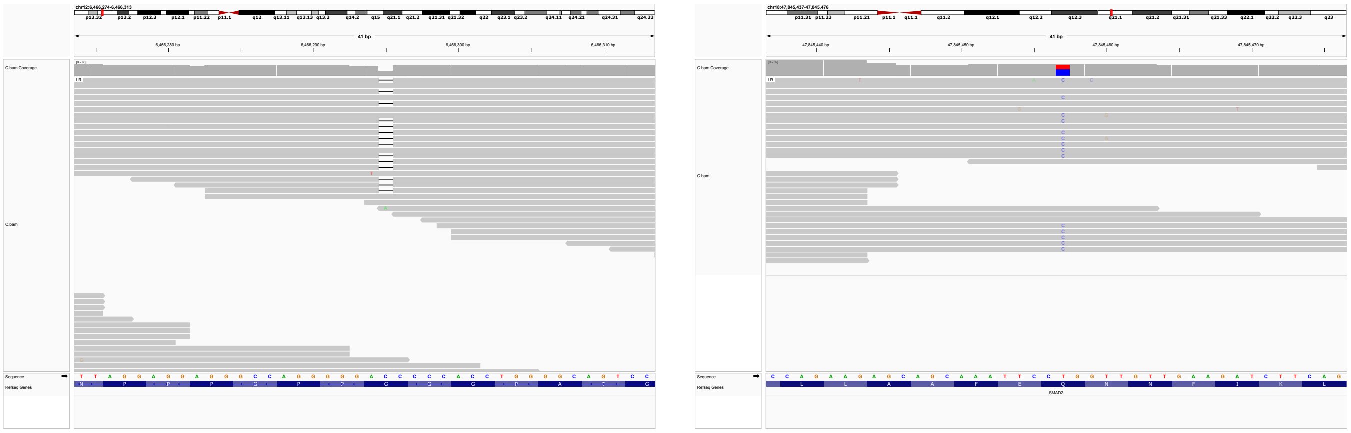

**Figure S29: P29.** Single-nucleotide deletion variant in *VAMP1*, 12-6466294-AC-A, ENST00000396308.4, c.59del, p.(Gly20Valfs*9). ACMG classification P: PP4,PP5,PVS1. Listed as pathogenic in ClinVar (diagnoss: Spastic paraplegia, one submission; Variation ID: 1459195). An additional heterozygous variant was observed in SMAD2, ENST00000262160.11, c.1163A>G,p.(Gln388Arg), that is interpreted in ClinVar 1 Path, 2 VUS (Variation ID: 1327533), interpreted as pathogenic in PMID:26247899. Autosomal dominant spastic ataxia-1 (SPAX1) is caused by heterozygous mutation in the VAMP1 gene (OMIM:108600).

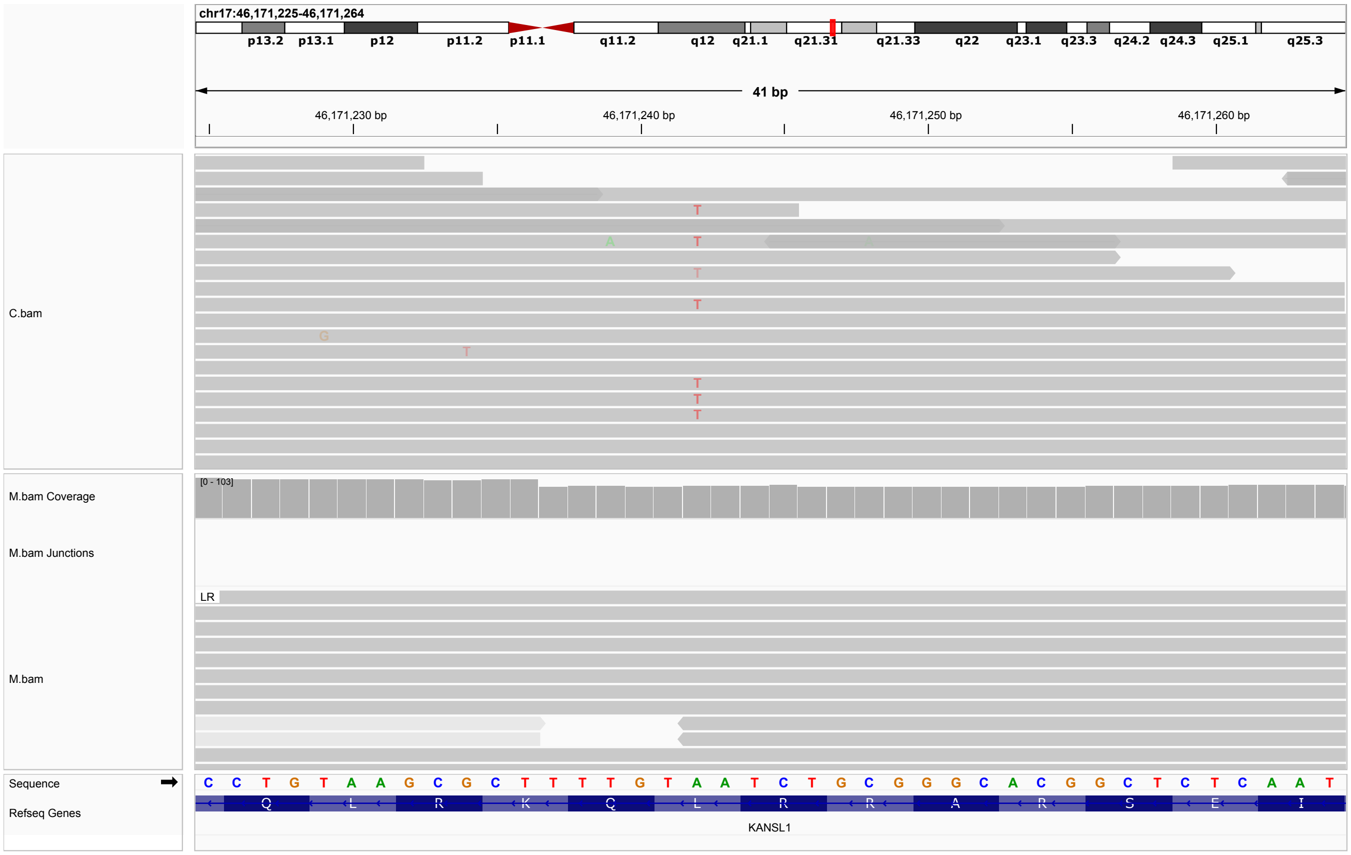


**Figure S30: P30.** Single-nucleotide substitution in *KANSL1*, 17-46171242-A-T, ENST00000432791.7, c.902T¿A, p.(Leu301*). ACMG classification LP: PVS1, PP4. Two entries in ClinVar (LP and VUS; Variation ID: 1014855). Koolen-de Vries syndrome (KDVS) can be caused by heterozygous mutation in the *KANSL1* gene. Paternal sample not available.

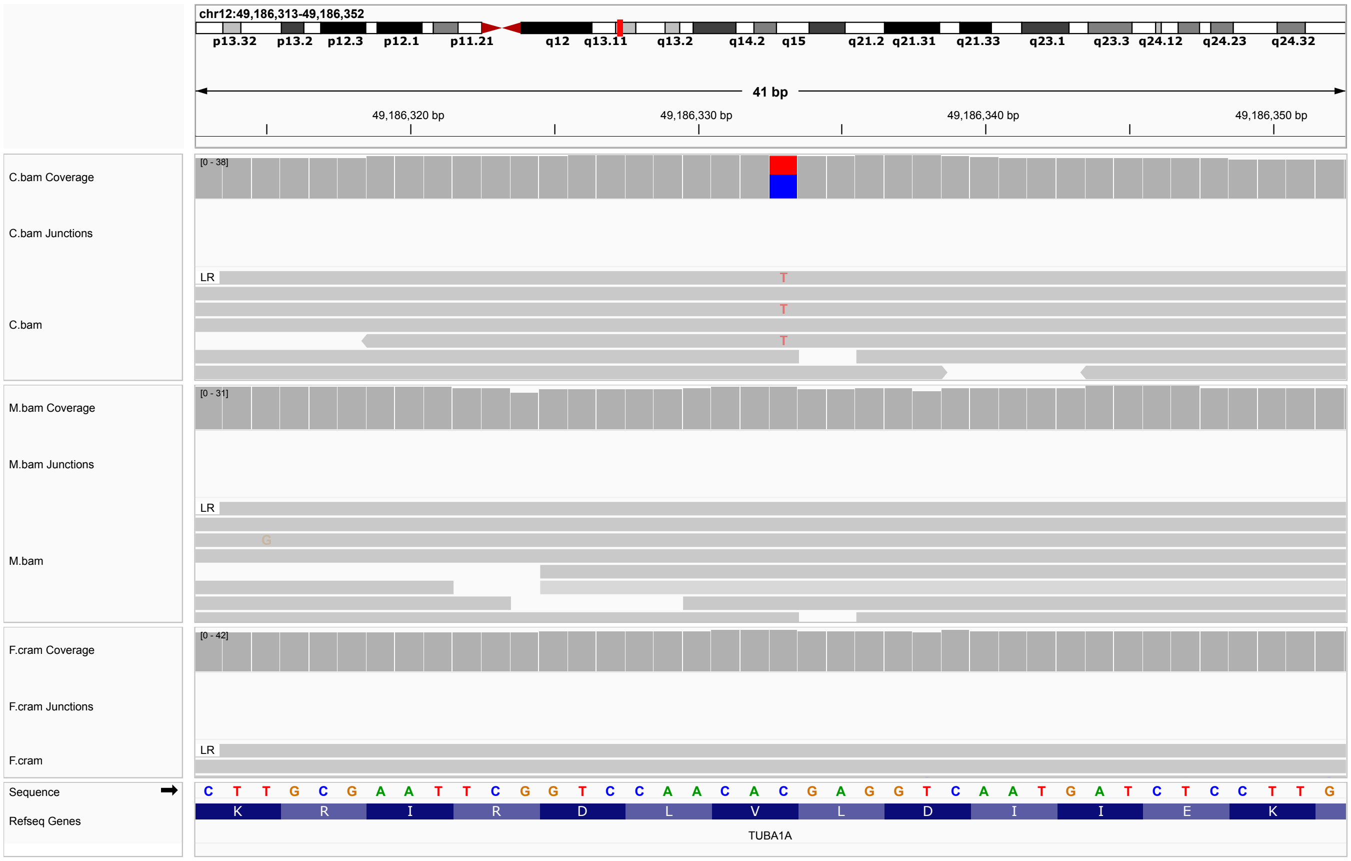


**Figure S31: P31.** Single-nucleotide substitution in *TUBA1A*, 12-49186333-C-T, ENST00000301071.12, c.352G¿A, p.(Val118Met). ACMG classification P: PM1, PM2_Supporting, PM5, PP2, PP5_Strong]. The variant was identified as a de novo event. ClinVar P/LP (Variation ID: 217023, four submissions). Lissencephaly-3 (LIS3) is caused by heterozygous mutation in the *TUBA1A* gene. An identical variant was reported previously [5].

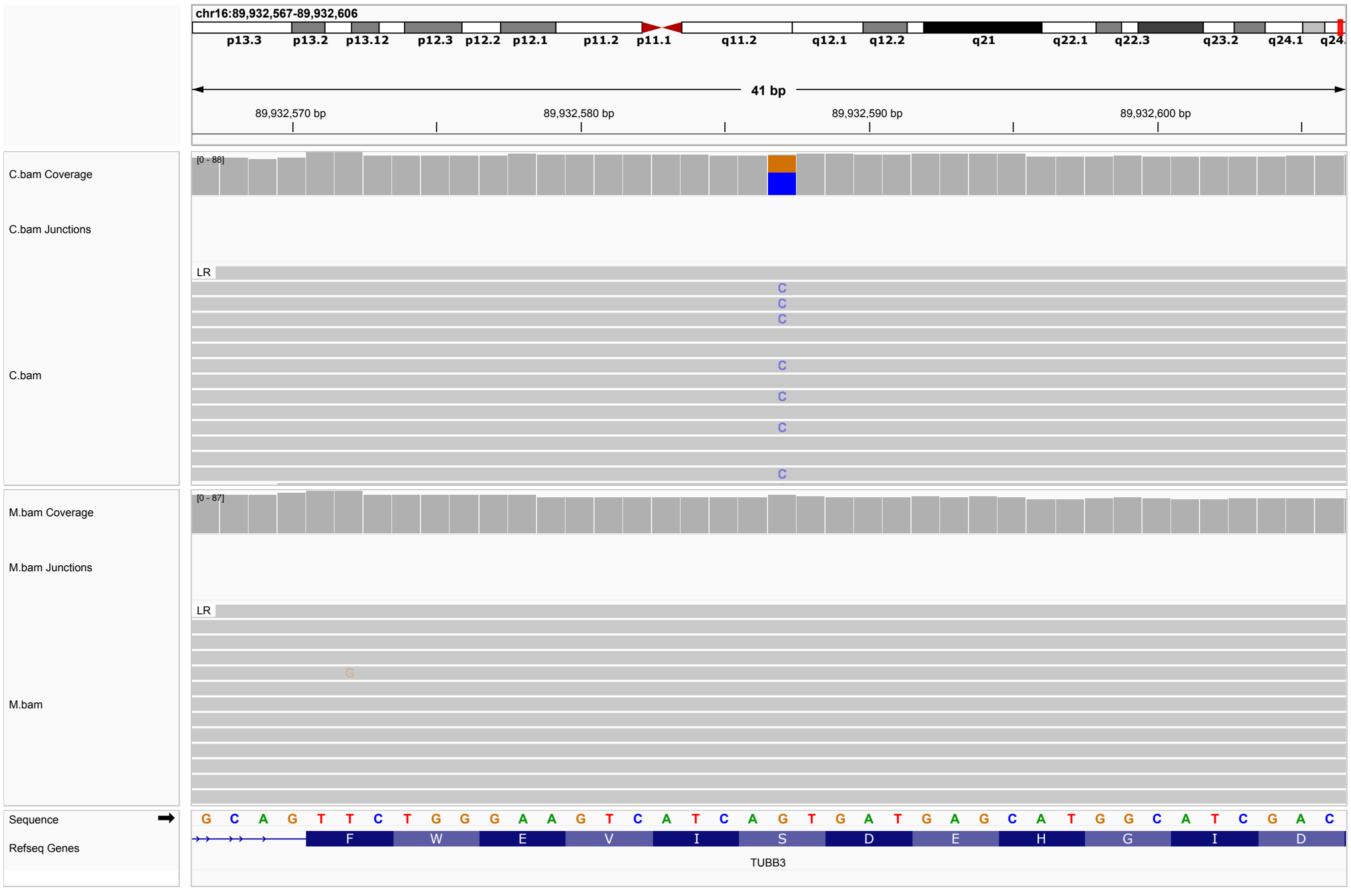


**Figure S32: P32.** Single-nucleotide substitution in *TUBB3*, 16-89932587-G-C, ENST00000315491.12, c.74G¿C, p.(Ser25Thr). ACMG classification LP: PM2,PM5,PP2,PP4. Not listed in ClinVar, but another variant at the same residue is listed as LP: NM_006086.4(TUBB3):c.74G>A (p.Ser25Asn) (Variation ID: 3340227). Complex cortical dysplasia with other brain malformations-1 (OMIM:614039) is caused by heterozygous mutation in the TUBB3 gene.

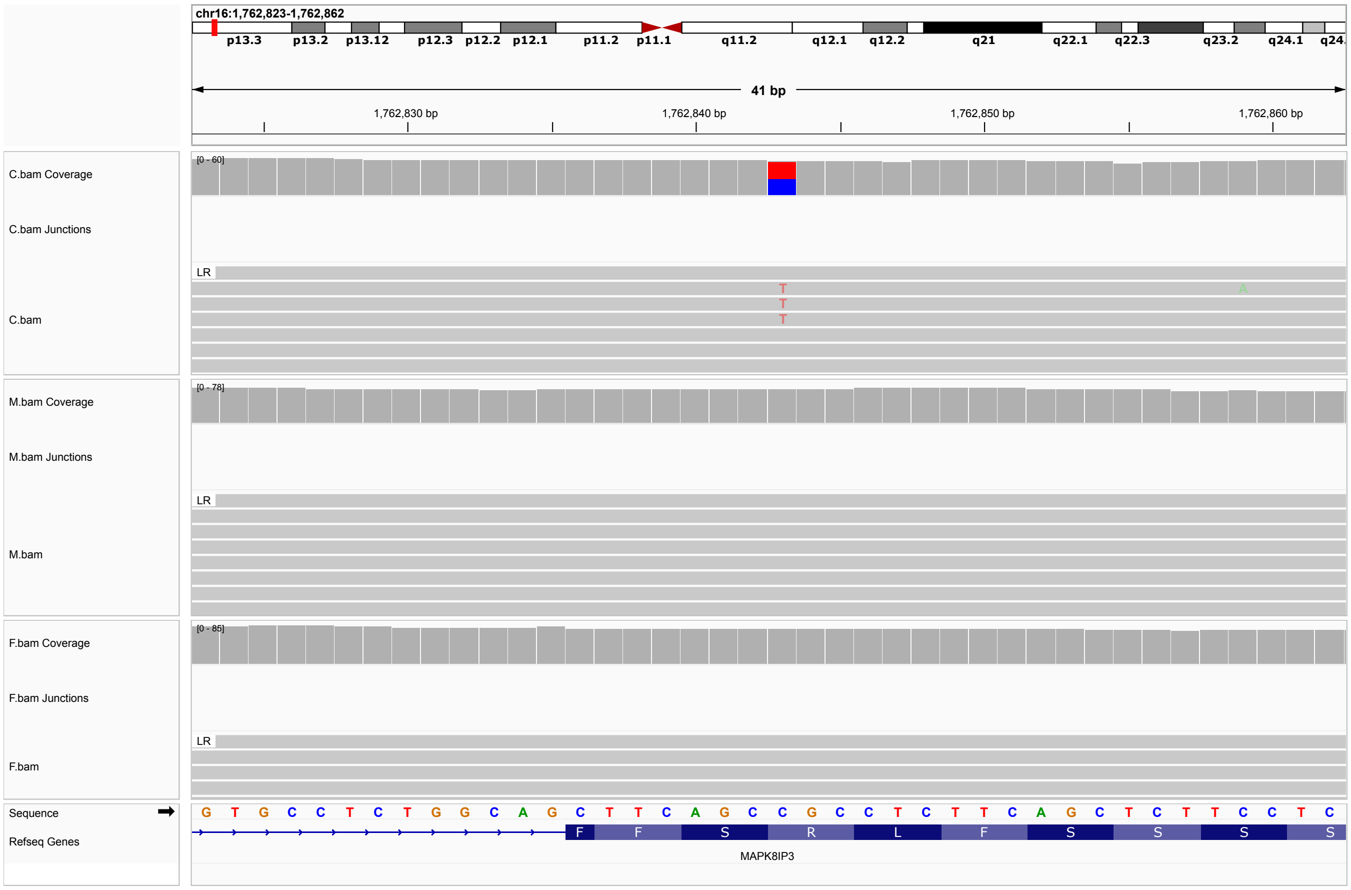


**Figure S33: P33.** Single-nucleotide substitution in *MAPK8IP3*, 16-1762843-C-T, ENST00000610761.2, c.1735C¿T, p.(Arg579Cys). ACMG classification P: PS2, PM2_Supporting, PP3, PP4, PP5_Strong. The identical variant is classified as P/LP in ClinVar (9 submissions, Variation ID: 632564). Variants in MAPK8IP3, including recurrent variants to be associated with Neurodevelopmental disorder with or without variable brain abnormalities (OMIM:618443). Variant found to be de novo.

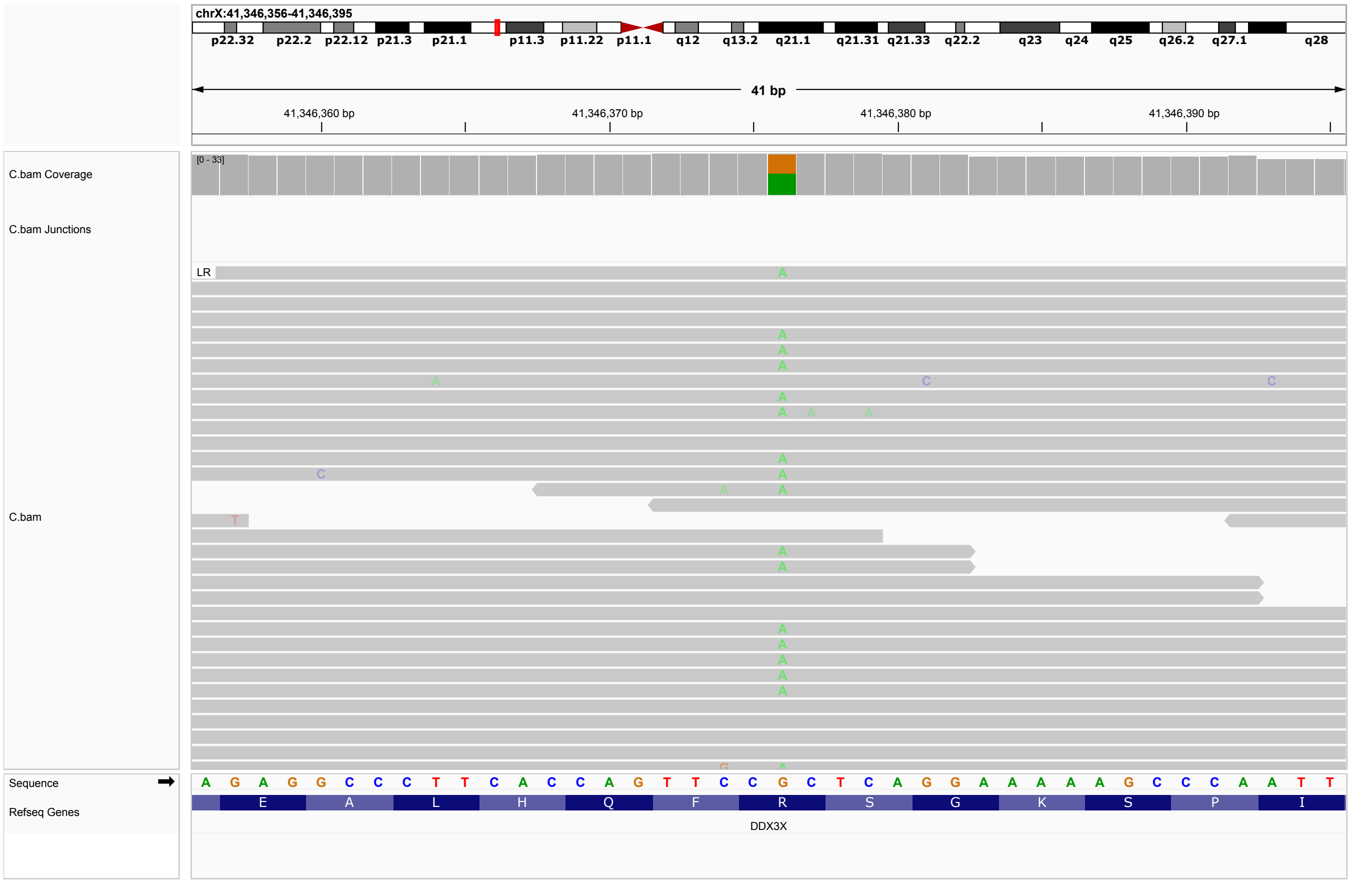


**Figure S34: P34.** Single-nucleotide substitution in *DDX3X*, SNV X-41346376-G-A, ENST00000644876.2, c.1463G¿A, p.(Arg488His). ACMG classification LP: PM1, PM2_Supporting, PM5_Supporting, PP3, PP5_Strong. The identical variant is listed as P/LP in ClinVar (4 submissions, Variation ID: 207817). The variant is a heterozygous X chromosomal variant in a female child, parental samples were not available. De novo heterozygous variants in DDX3 have been identified in females with ID and various other features including hypotonia, movement disorders, behavior problems, corpus callosum hypoplasia, and epilepsy [6], Intellectual developmental disorder, X-linked syndromic, Snijders Blok type (OMIM:300958).

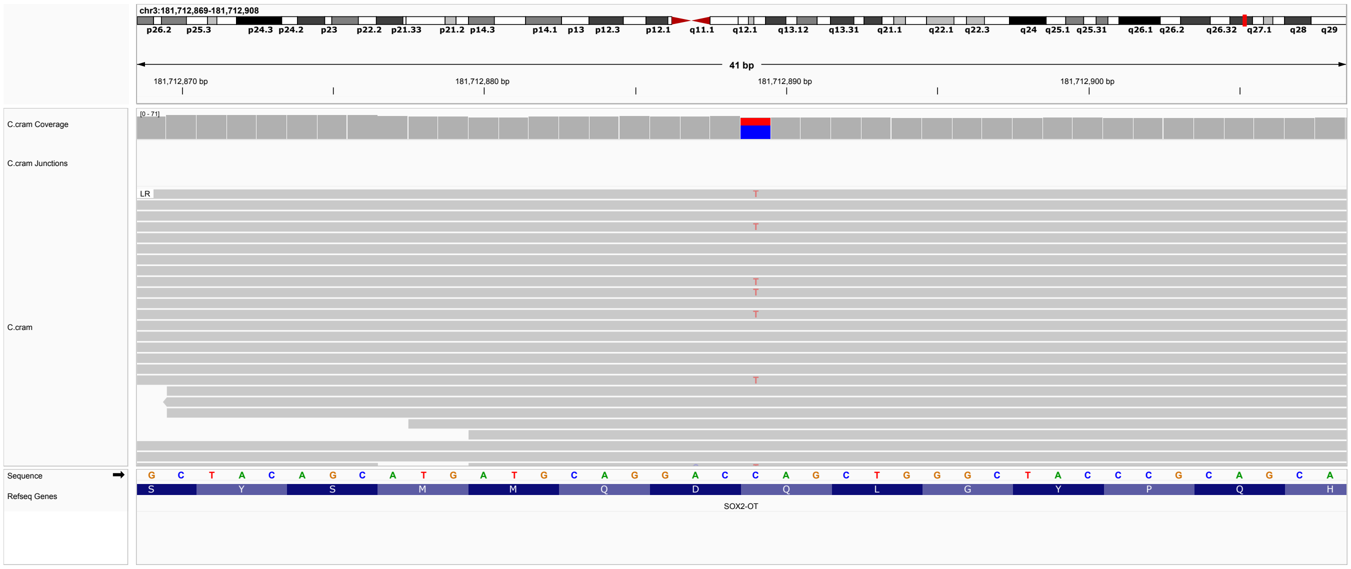


**Figure S35: P35.** Single-nucleotide substitution in *SOX2*, 3-181712889-C-T, ENST00000325404.3, c.529C¿T, p.(Gln177*). ACMG classification LP: PVS1_Strong, PM2_Supporting, PP5. This variant is listed as pathogenic in ClinVar (2 entries, Variation ID: 12814). Variants in SOX2 are assocaited with Optic nerve hypoplasia and abnormalities of the central nervous system as well as Microphthalmia, syndromic 3 (OMIM:206900). The identical variant has been previous published [7, 8]. Some individuals with truncating SOX2 variants have been observed to have spastic diplegia [7, 8]. Parental samples not available.

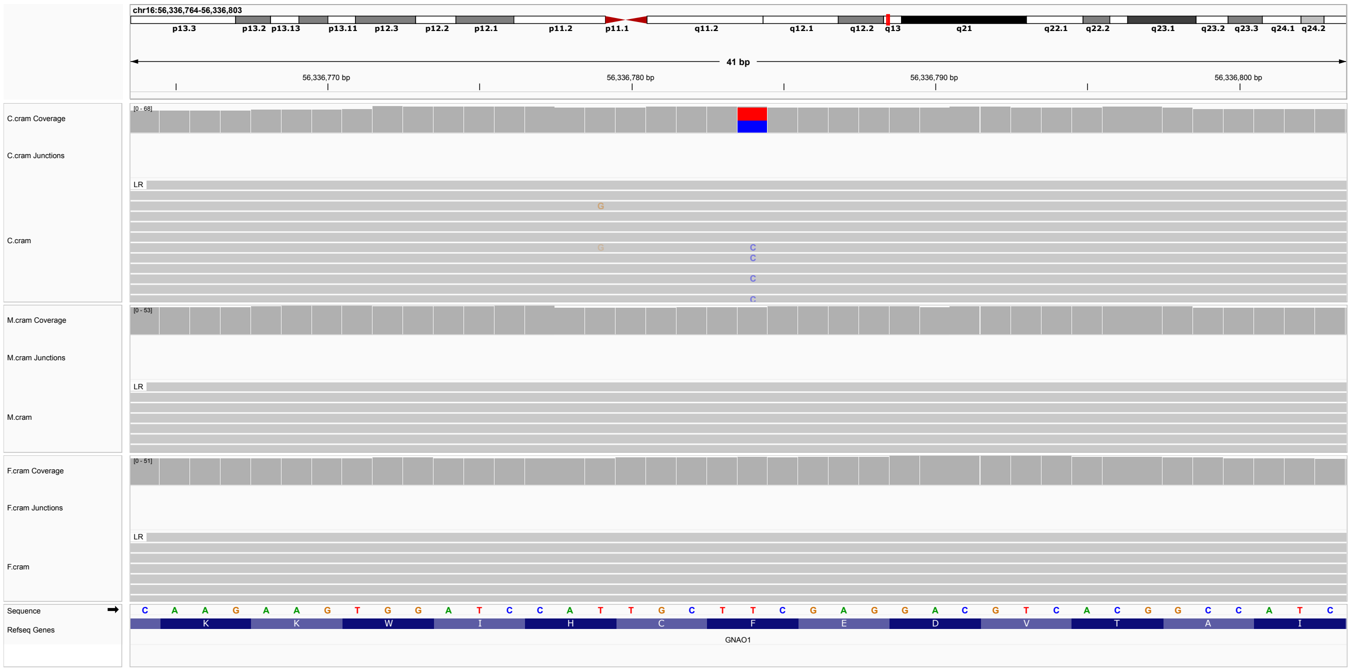


**Figure S36: P36.** De novo Single-nucleotide substitution in *GNAO1*, 16-56336784-T-C, ENST00000262493.12, c.647T¿C, p.(Phe216Ser). ACMG classification P: PS2, PP3, PP4, PM1, PM2, PP2. Not listed in ClinVar. Pathogenic variants in GNAO1 are associated with Developmental and epileptic encephalopathy 17 (OMIM:615473) and Neurodevelopmental disorder with involuntary movements (OMIM:617493).

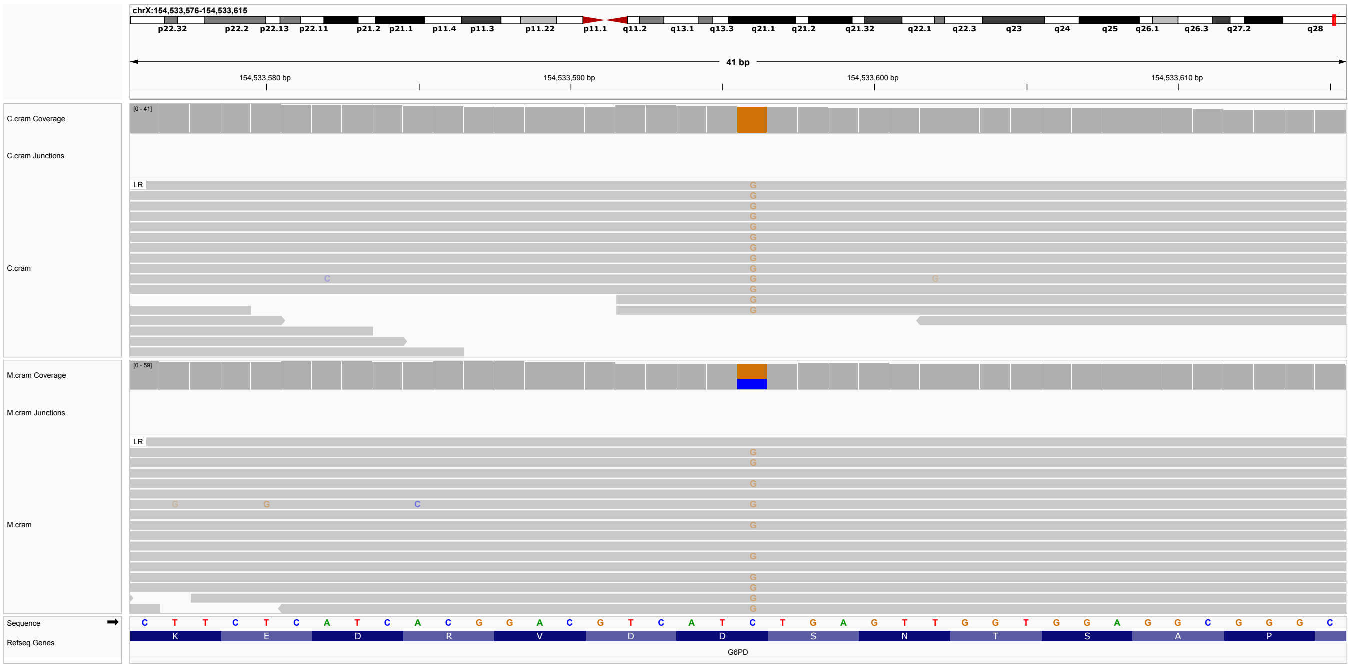


**Figure S37: P37.** Single-nucleotide substitution in *G6PD*, X-154533596-C-G, ENST00000393562.10, c.844G¿C, p.(Asp282His). ACMG classification P: PM1, PM5_Supporting, PP2, PP3_Moderate, PP5_Strong. Variant listed in ClinVar as pathogenic (19 entries, Variation ID: 10372). Paternal sample not available. Mother is heterozygous, male child is hemizygous for the variant. The variant was not present in the affected sibling of P37.

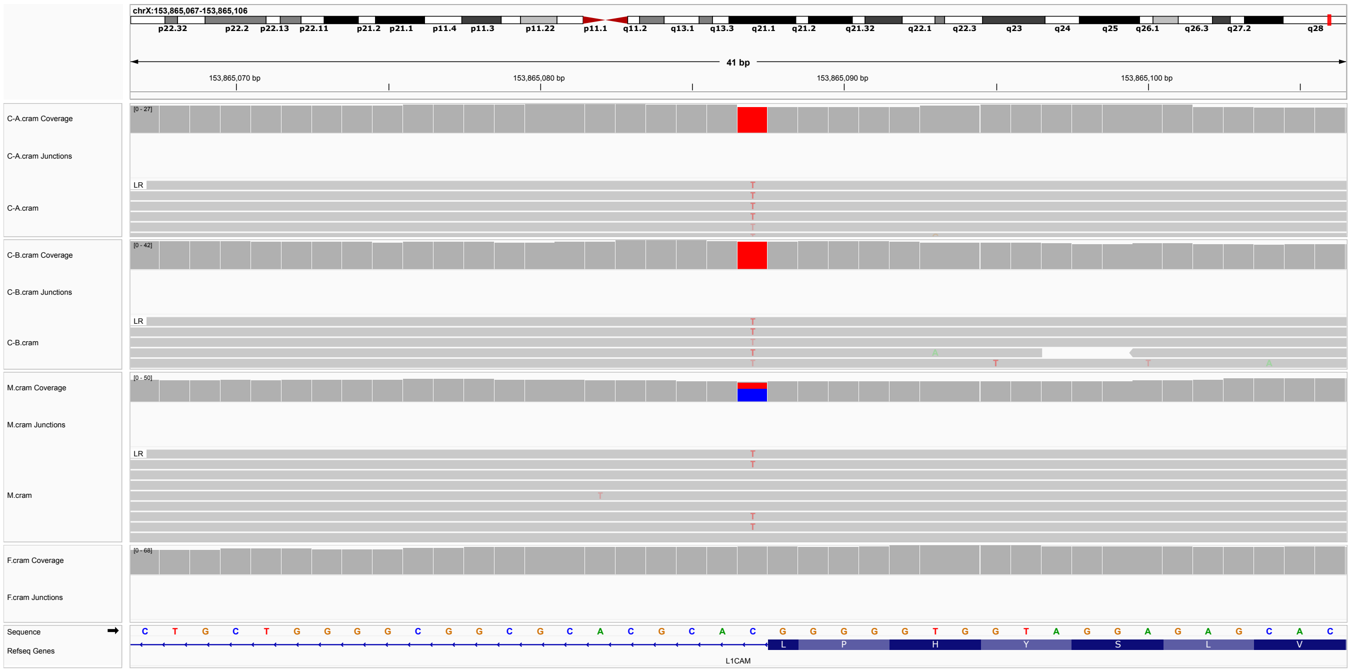


**Figure S38: P38.** Single-nucleotide substitution in *L1CAM*, X-153865087-C-T, ENST00000370060.7, c.2872+1G¿A, p.? ACMG classification P: PVS1,PP5,PM2. Variant listed in ClinVar as pathogenic (Variation ID: 2737422). Variants in L1CAM are associated with the X-chromosomal recessive conditions Corpus callosum, partial agenesis of (OMIM:304100), Hydrocephalus, congenital, X-linked (OMIM:307000), and MASA syndrome (OMIM:303350). Both brothers show a hemizygous variant inherited from the heterozygous mother.

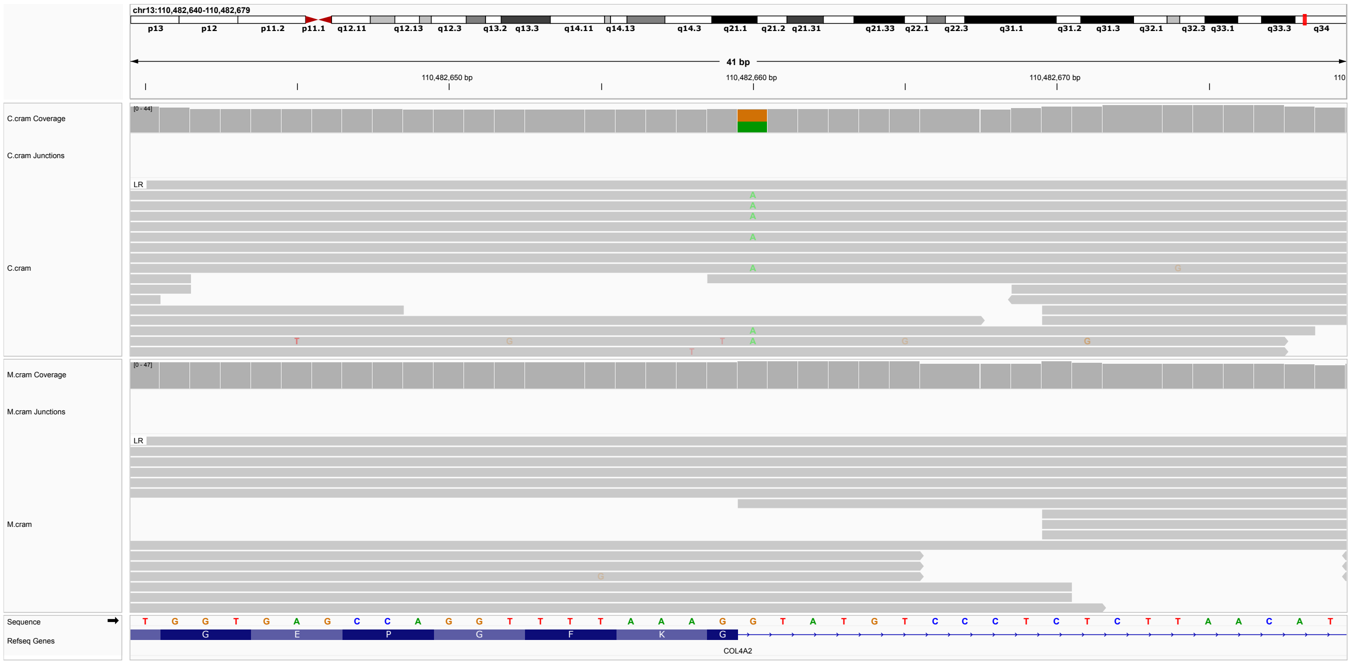


**Figure S39: P39.** Single-nucleotide substitution in *COL4A2*, 13-110482660-G-A, ENST00000360467.7, c.2902+1G¿A, p.? ACMG classification P: PVS1, PM2_Supporting, PP4, PP5. The variant is listed in ClinVar as LP (One submission, Variation ID: 864858). Brain small vessel disease-2 (BSVD2) is caused by heterozygous mutation in the COL4A2 gene (OMIM:614483). Paternal sample not available.

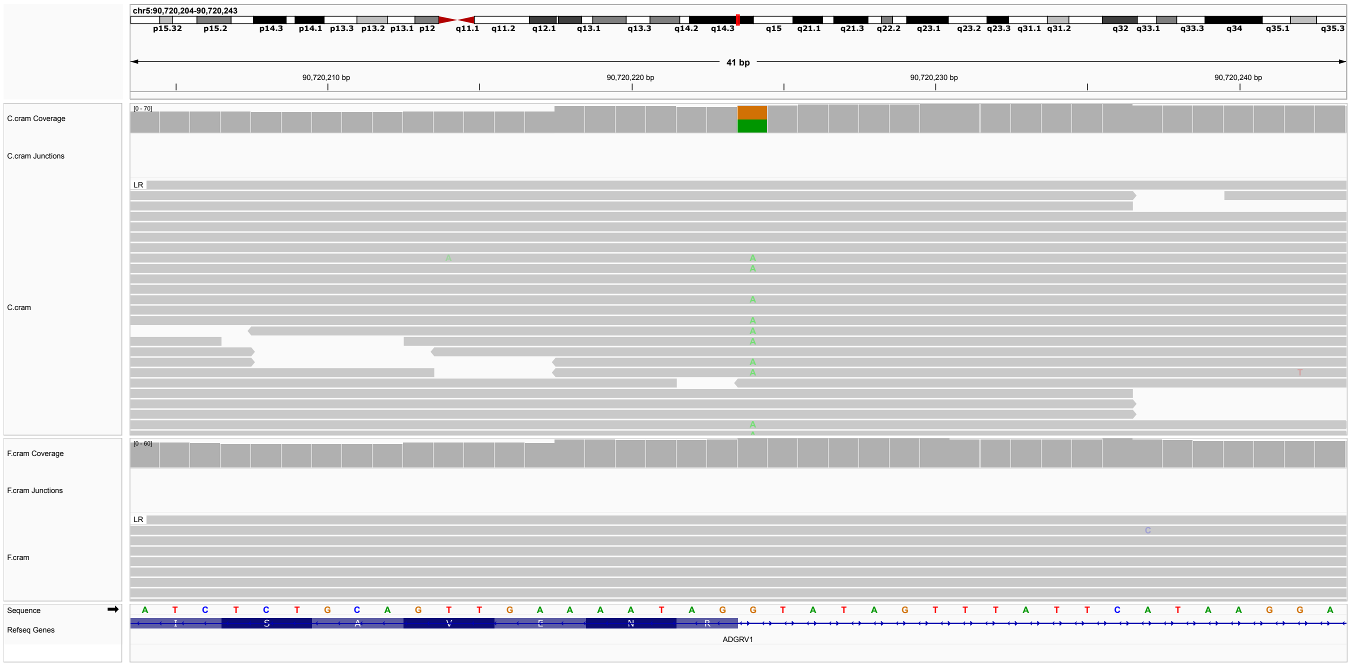


**Figure S40: P40.** Single-nucleotide substitution in *ADGRV1*, SNV 5-90720224-G-A, ENST00000405460.9, c.9623+1G¿A, p.?. ACMG classification P: PVS1, PP4, PP5_Strong. Listed in ClinVar as LP (three submissions, Variation ID: 1207273). The variant has been published in heterozygosity in an individual with Usher syndrome [9]. Paternal sample not available.

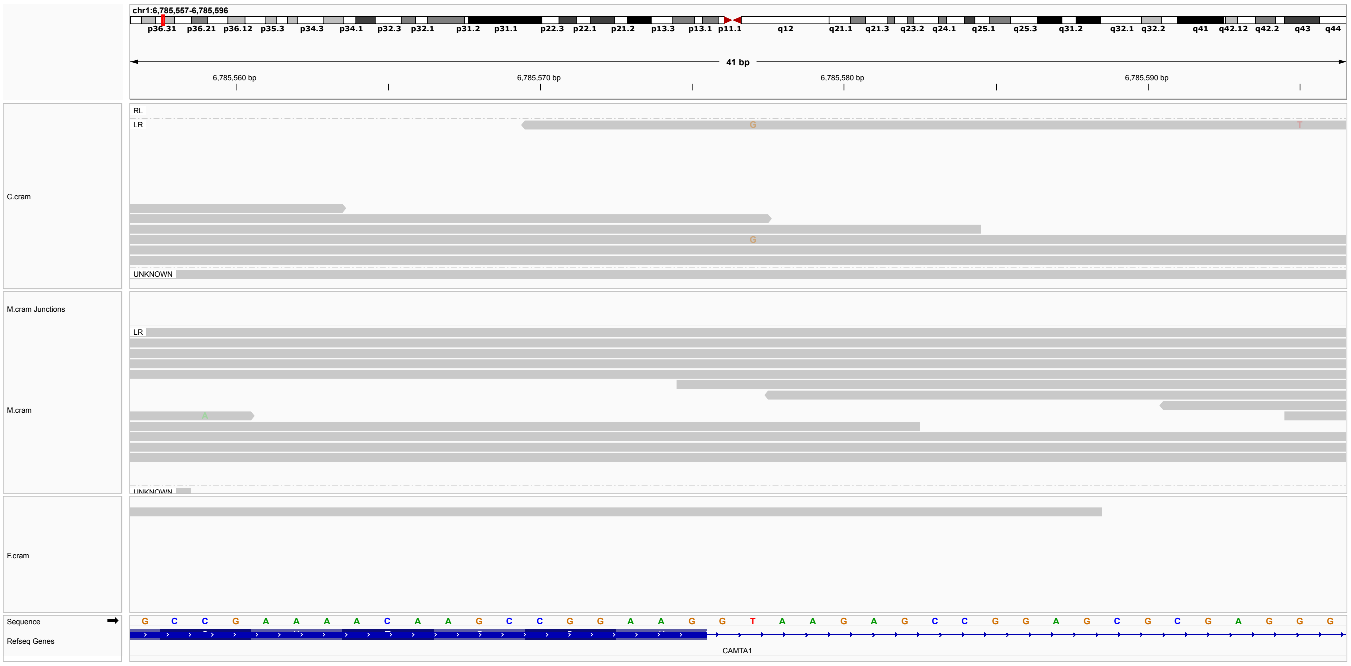

**Figure S41: P41.** De novo single-nucleotide substitution in *CAMTA1*, 1-6785577-T-G, ENST00000303635.12, c.45+2T¿G, p.?. ACMG classification P: PVS1, PS2, PM2_Supporting, PP4. Not listed in ClinVar. Pathogenic variants in *CAMTA1* are associated with Cerebellar dysfunction with variable cognitive and behavioral abnormalities (OMIM:614756).

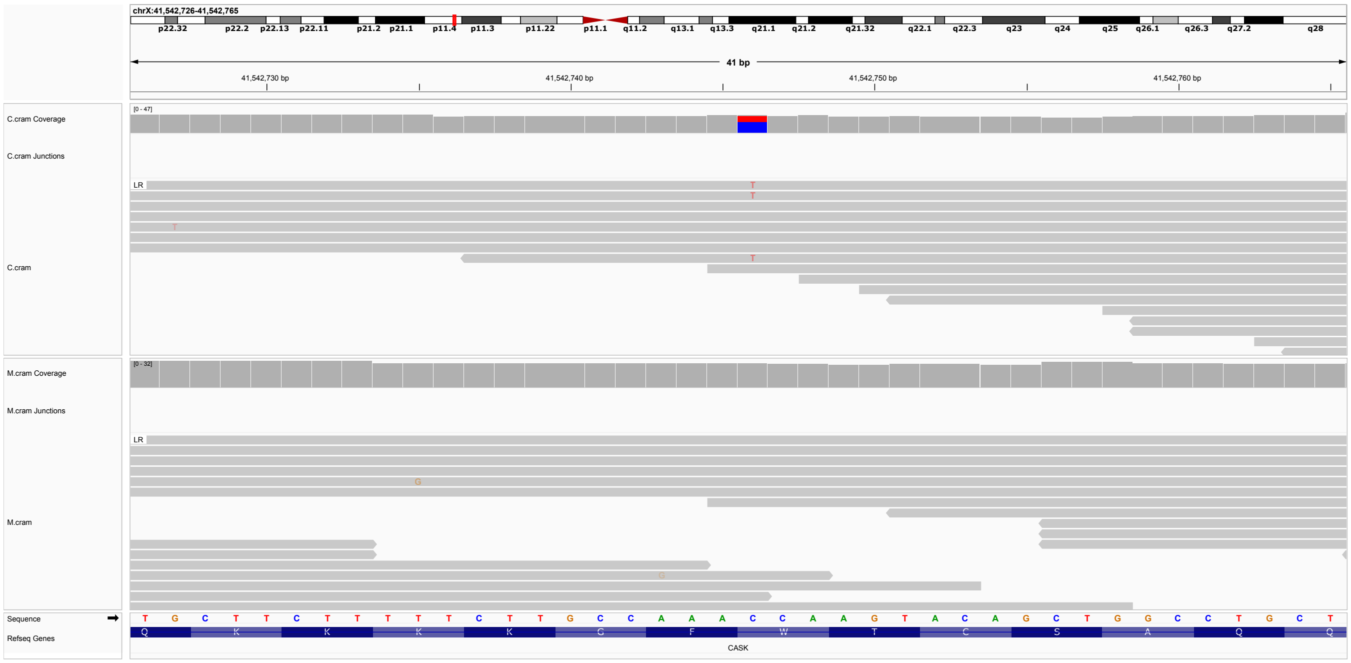


**Figure S42: P42.** A single-nucleotide substitution in *CASK*, X-41542746-C-T, ENST00000378163.7, c.2100G¿A, p.(Trp700*). ACMG classification P: PP5,PP4,PM2,PVS1. Variant listed in ClinVar as pathogenic (one submission, Variation ID: 265654). Variant found here in a female child, paternal sample not available. Intellectual developmental disorder with microcephaly and pontine and cerebellar hypoplasia (MICPCH) is an X-linked disorder affecting males and females. In females, it is characterized by severely impaired intellectual development and variable degrees of pontocerebellar hypoplasia. Affected females have poor psychomotor development, often without independent ambulation or speech, and axial hypotonia with or without hypertonia. De novo stop-gain variants have been previously published in girls with this condition [10].

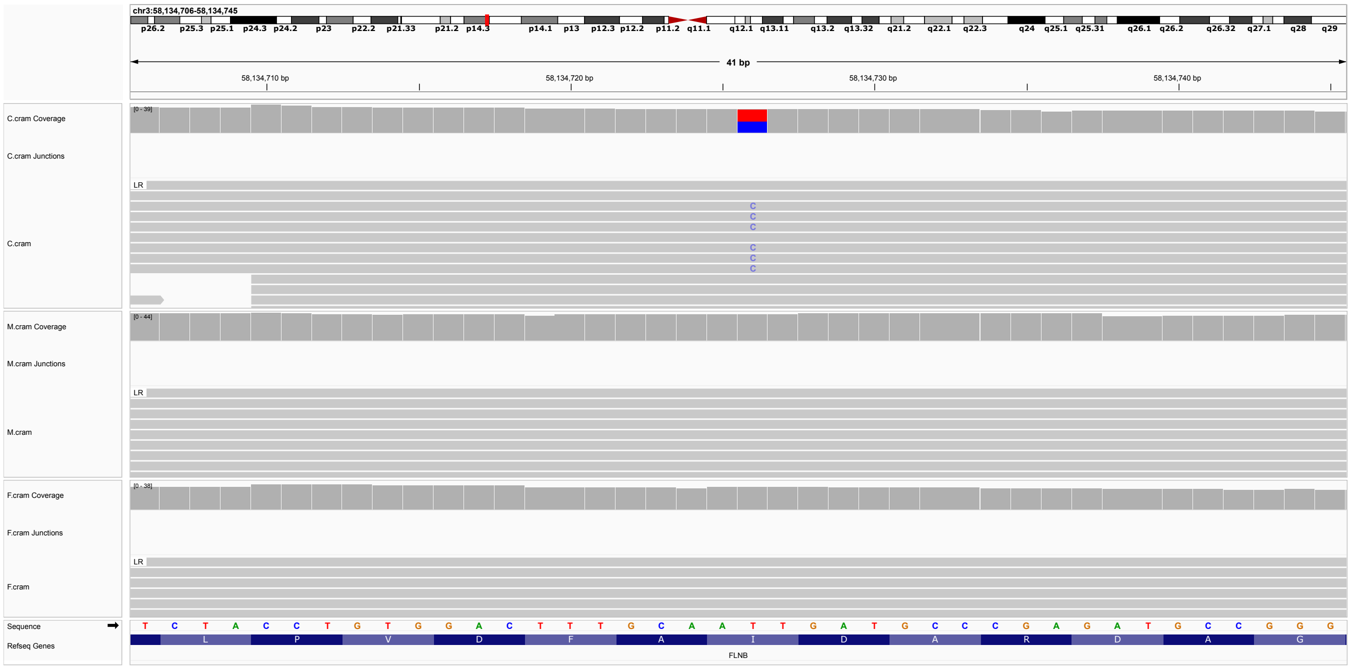


**Figure S43: P43.** A de novo single-nucleotide substitution in *FLNB*, 3-58134726-T-C, ENST00000295956.9, c.4625T¿C, p.(Ile1542Thr). ACMG classification LP: PS2, PM2_Supporting, PP3_Moderate]. Variant is represented in ClinVar as no assertion provided; there are four submissions, one pathogenic, one likely pathogenic, and two VUS (Variation ID: 801980). An identical variant was previously published as a de novo event in an individual with Larsen syndrome [11]. Variants in *FLNB* are associated with diseases including Larsen syndrome (OMIM:150250). Cerebral palsy is not a known association of Larsen syndrome, but speculatively cervical spinal cord compression or other pathology could be related [12].

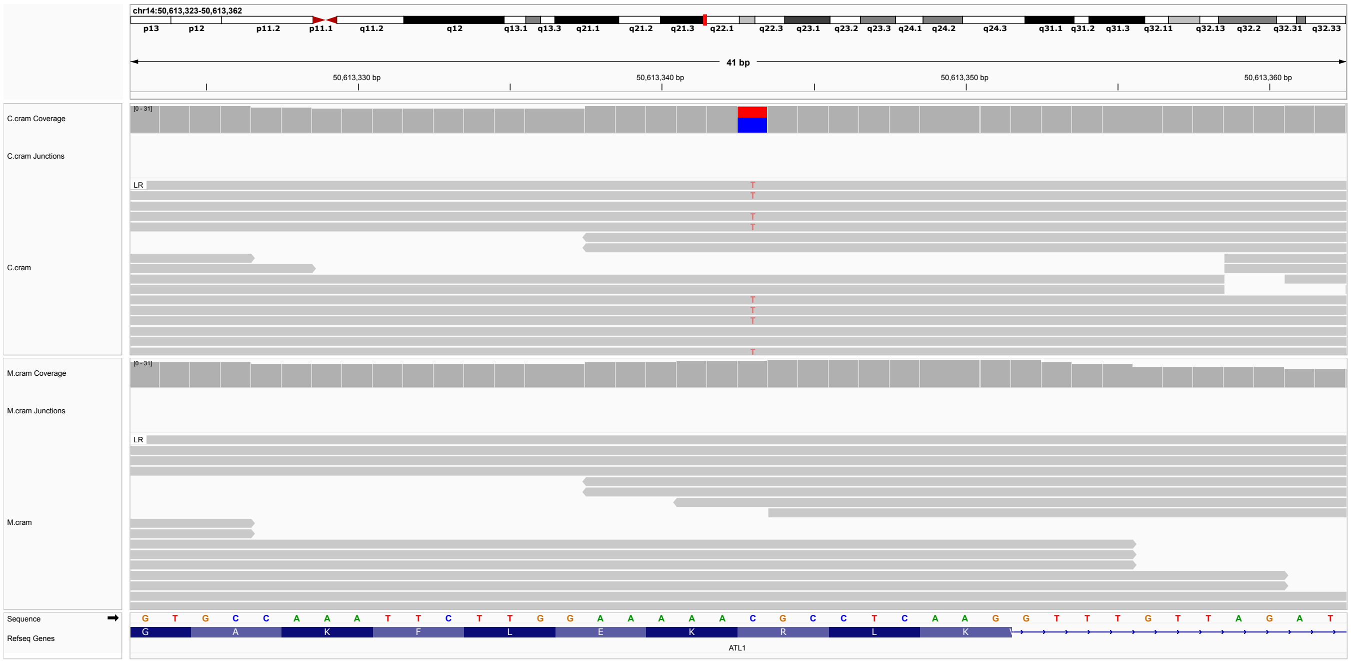


**Figure S44: P44.** A single-nucleotide substitution in *ATL1*, 14-50613343-C-T, ENST00000358385.12, c.715C¿T,p.(Arg239Cys). ACMG classification P: PS2, PM5_Supporting, PP2, PP4, PP5_Strong. Variant listed in ClinVar as P/LP (17 submissions, Variation ID: 4346) Variants in ATL1 have been associated with Spastic paraplegia 3A, autosomal dominant (OMIM:182600). The variant has been published in an individual with spastic paraplegia and shown to disrupted BMPRII trafficking to the cell surface [13].

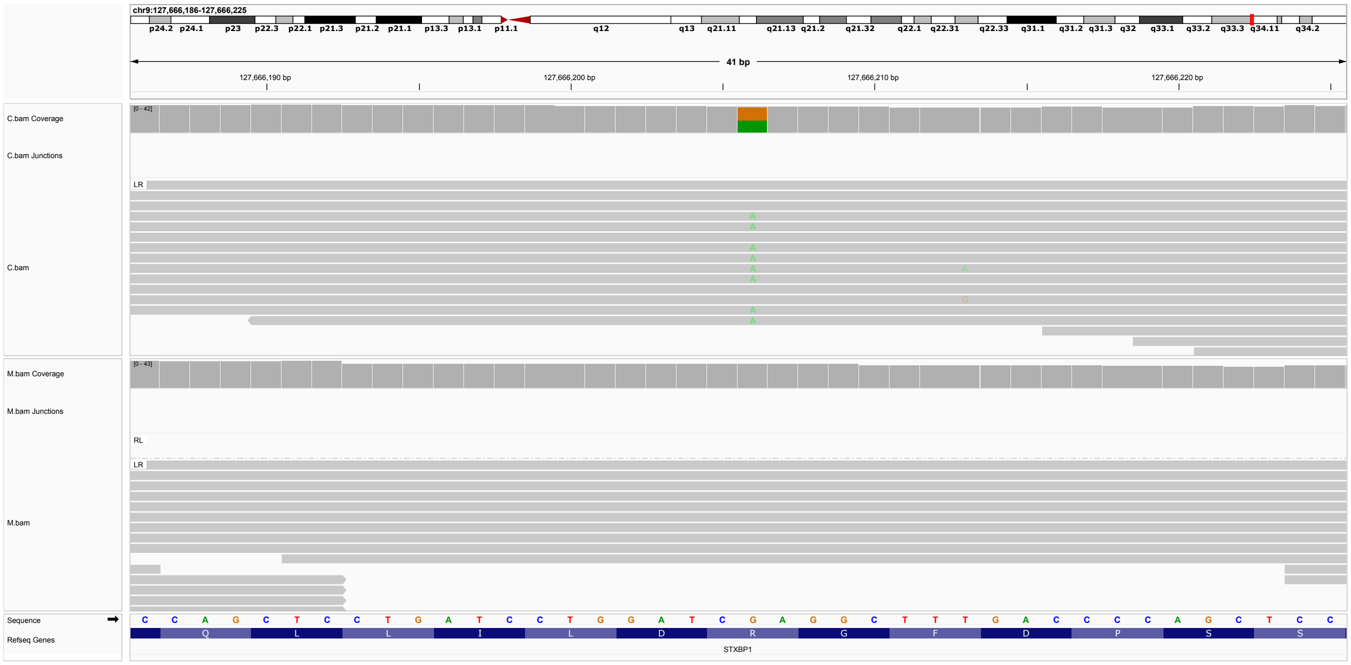

**Figure S45: P45.** A single-nucleotide substitution in *STXBP1*, 9-127666206-G-A, ENST00000373299.5, c.704G¿A, p.(Arg235Gln). ACMG classification P: PM2_Supporting, PP3_Strong, PP4, PP5_Strong. The variant is listed in ClinVar as pathogenic (six submissions, Variation ID: 199083). Developmental and epileptic encephalopathy-4 (DEE4) is caused by heterozygous mutation in the STXBP1 gene (OMIM:612164). The variant was published previously [14].

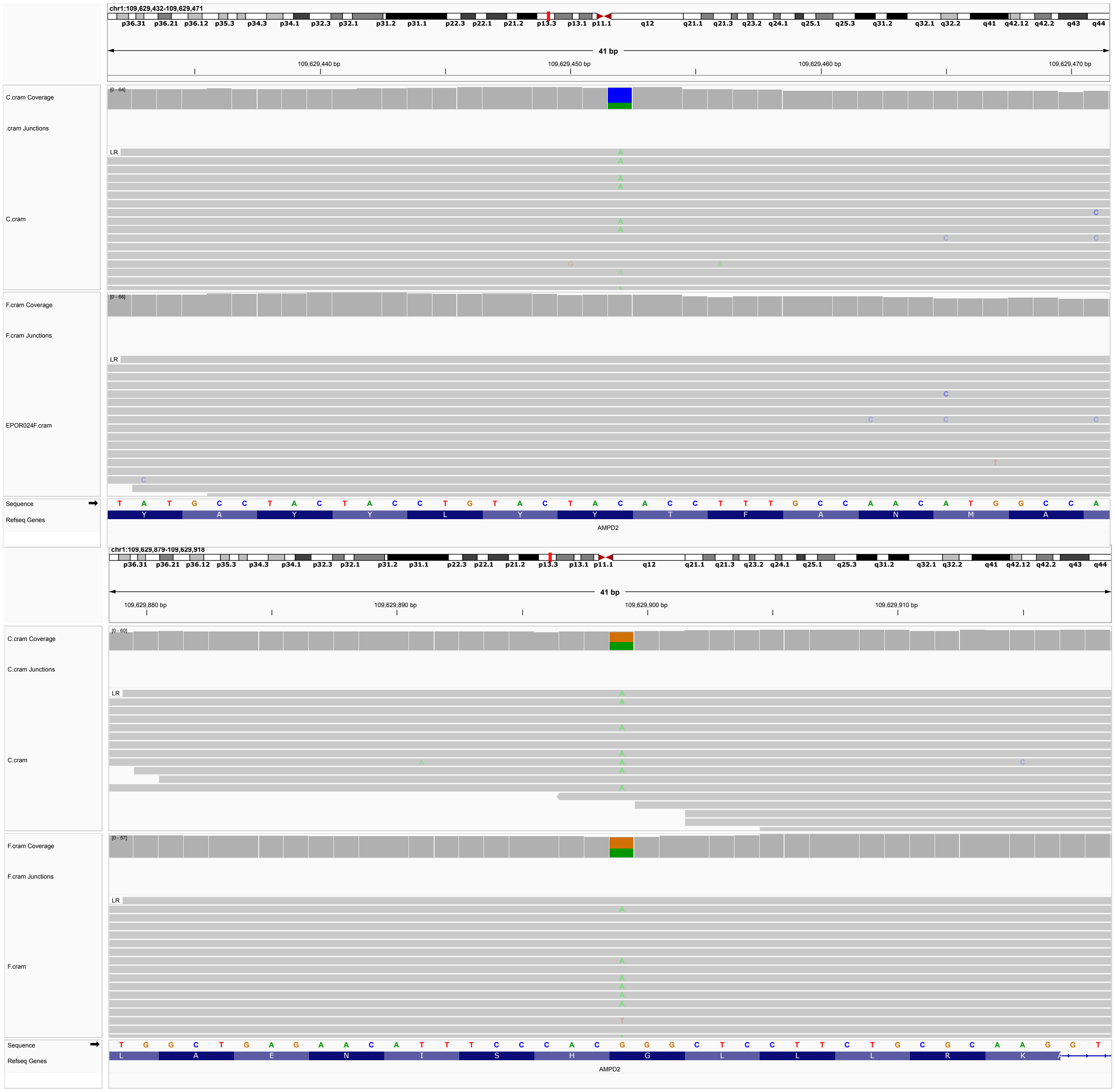

**Figure S46: P46.** Compound heterozygous single-nucleotide substitutions in *AMPD2*, Maternal chromosome: 1-109629452-C-A, ENST00000528667.7, c.1824C¿A, p.(Tyr608*). ACMG classification P: PVS1, PM2_Supporting, PP4. Not listed in ClinVar. Paternal chromosome: 1-109629899-G-A, ENST00000528667.7, c.1966G¿A, p.(Gly656Arg). ACMG classification LP: PM2_Supporting, PP3_Strong, PP4. Not listed in ClinVar. Biallelic pathogenic variants in *AMPD2* in spastic paraplegia-63 (OMIM:615686) or Pontocerebellar hypoplasia, type 9 (OMIM:615809).

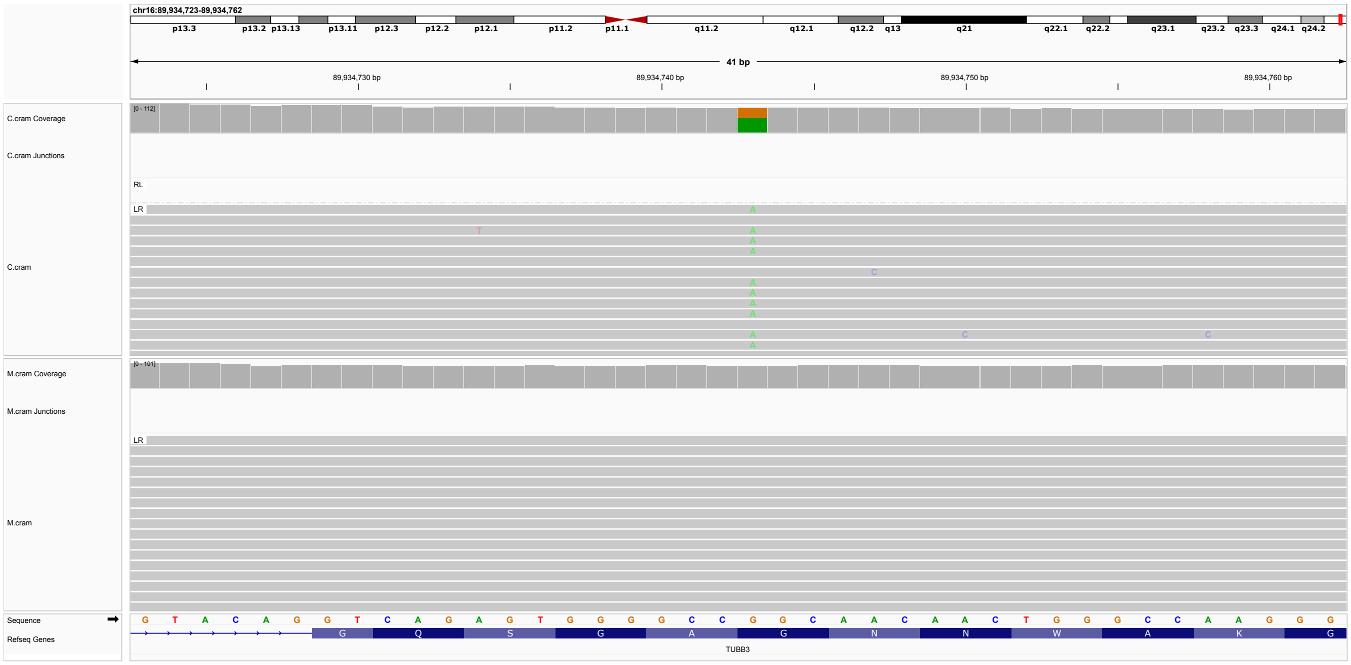


**Figure S47: P47.** A single-nucleotide substitutions in *TUBB3*, 16-89934743-G-A, ENST00000315491.12, c.292G¿A, p.(Gly98Ser) ACMG classification LP: PM2, PP2, PP3, PP4, PP5. Variant listed in ClinVar as P/LP (5 submissions, Variation ID: 160191). Paternal sample not available.

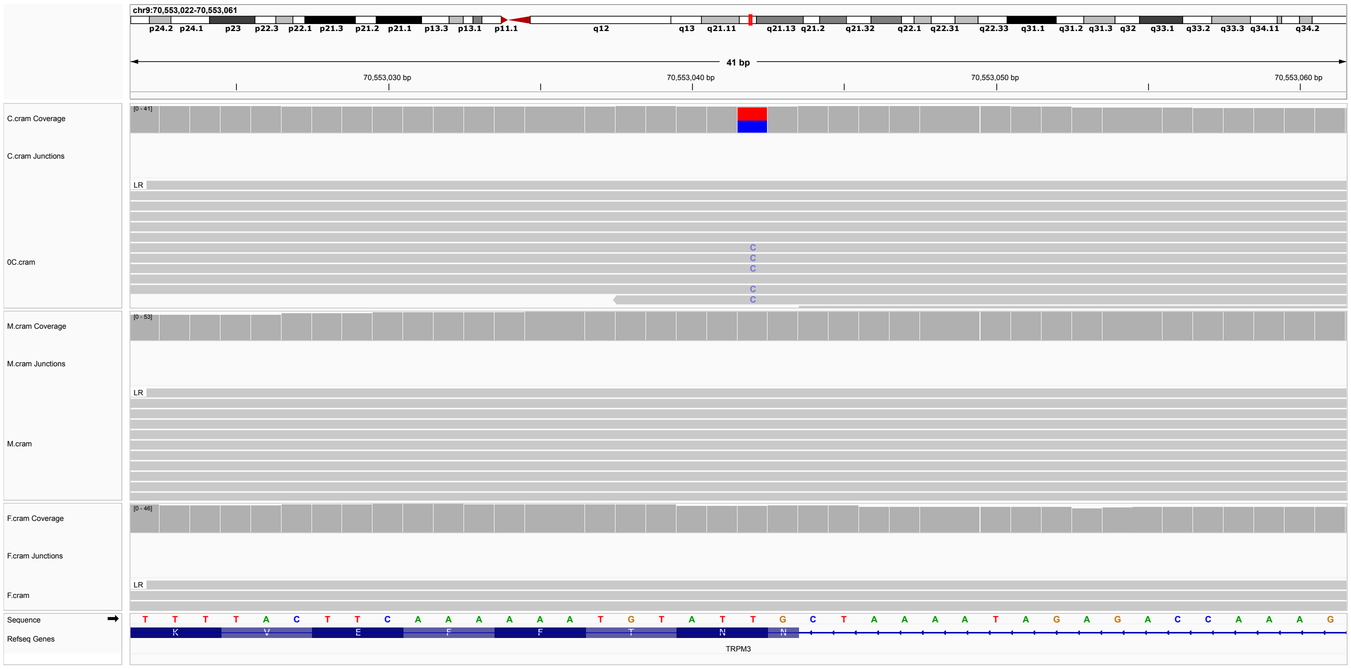


**Figure S48: P48.** A single-nucleotide substitutions in *TRPM3*, 9-70553042-T-C, ENST00000677713.2, c.3376A¿G, p.(Asn1126Asp). ACMG classification P: PS2, PM2_Supporting, PP4, PP5_Strong Variant is listed in ClinVar as P/LP (Two submissions, Variation ID: 1193351). Neurodevelopmental disorder with hypotonia, dysmorphic facies, and skeletal anomalies, with or without seizures (NEDFSS), is caused by heterozygous mutation in the TRPM3 gene (OMIM:620224).

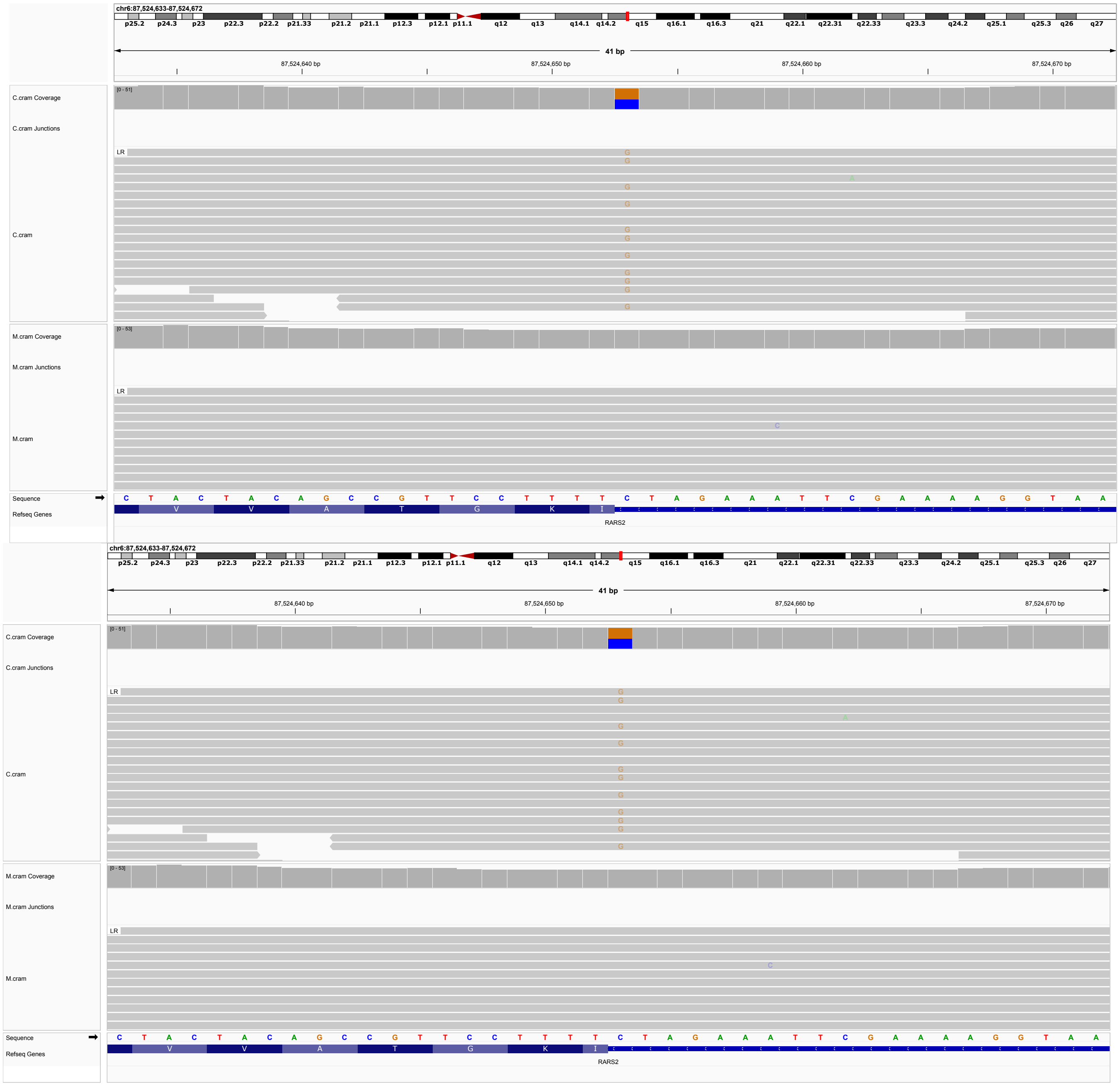


**Figure S49: P49.** Compound heterozygous single-nucleotide substitutions in *RARS2*, Paternal chromosome: 6-87524653-C-G, ENST00000369536.10, c.879-1G¿C, p.? ACMG classification P: PVS1, PS1_Supporting, PM2_Supporting, PP4. Variant listed in ClinVar as P/LP (three submissions, Variation ID: 1067780) Maternal chromosome: 6-87548623-A-C, ENST00000369536.10, c.419T¿G, p.(Phe140Cys). ACMG classification LP: PP3_Moderate, PP4, PP5_Strong, BP1. Variant listed in ClinVar as P/LP (9 submissions, Variation ID: 215055). Pontocerebellar hypoplasia type 6 (PCH6) is caused by homozygous or compound heterozygous mutation in the gene encoding mitochondrial arginyl-tRNA synthetase (OMIM:611523). Paternal sample not available.

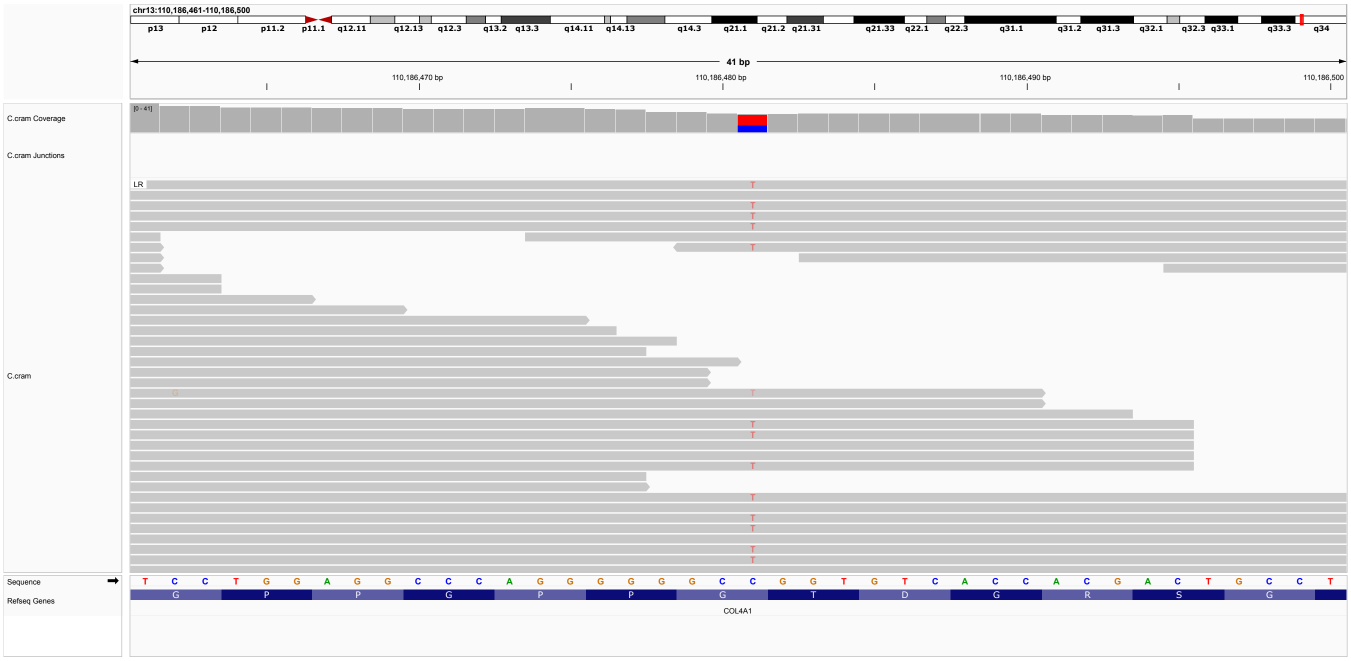


**Figure S50: P50.** Single-nucleotide substitution in *COL4A1*, 13-110186481-C-T, ENST00000375820.10, c.1801G¿A, p.(Gly601Ser). ACMG classification LP: PP3_Moderate, PP4, PP5_Strong. Variant listed in ClinVar as pathogenic (three submissions, Variation ID: 420899). Variants in COL4A1 are associated with several diseases with neurological manifestations, including Microangiopathy and leukoencephalopathy, pontine, autosomal dominant (OMIM:618564). The identical variant was previously published [15]

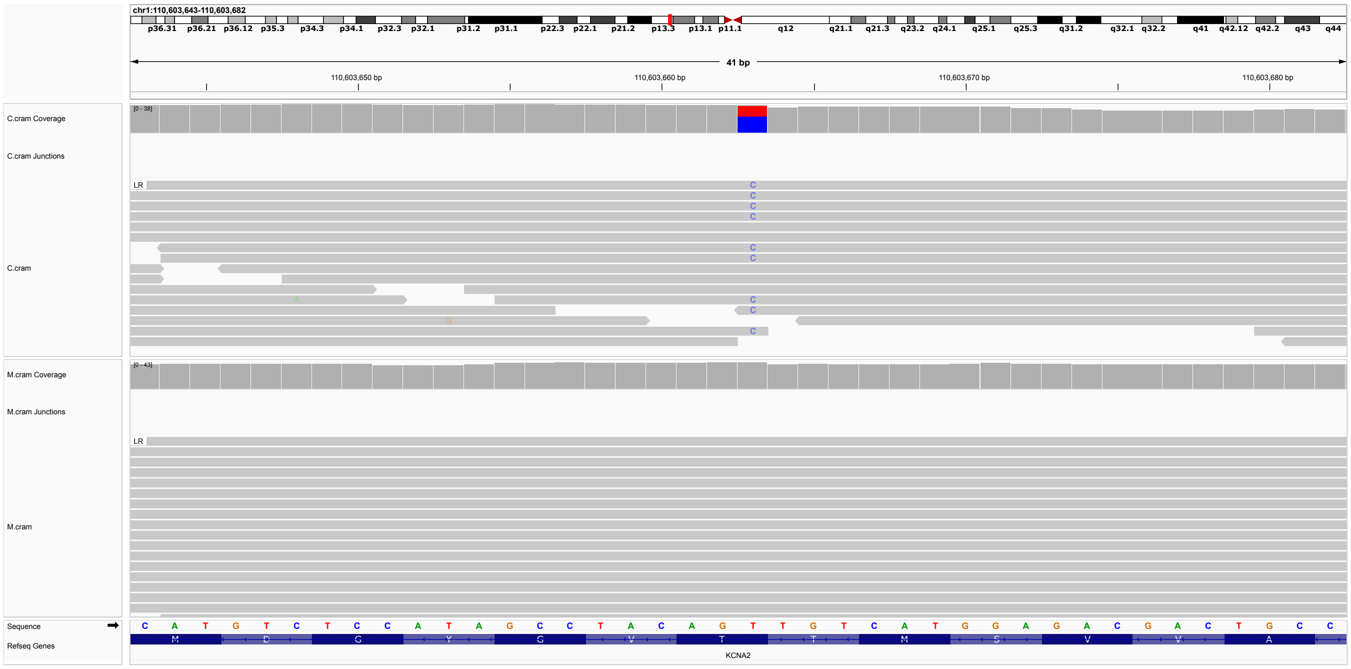


**Figure S51: P51.** Single-nucleotide substitution in *KCNA2*, 1-110603663-T-C, ENST00000316361.10, c.1120A¿G, p.(Thr374Ala). ACMG classification P: PM_Supporting, PP2, PP3_Strong, PP4, PP5_Strong. Variant listed in ClinVar as pathogenic (six submissions, Variation ID: 559647). Developmental and epileptic encephalopathy-32 (DEE32) is caused by heterozygous mutation in the KCNA2 gene (OMIM:616366). Paternal sample not available.

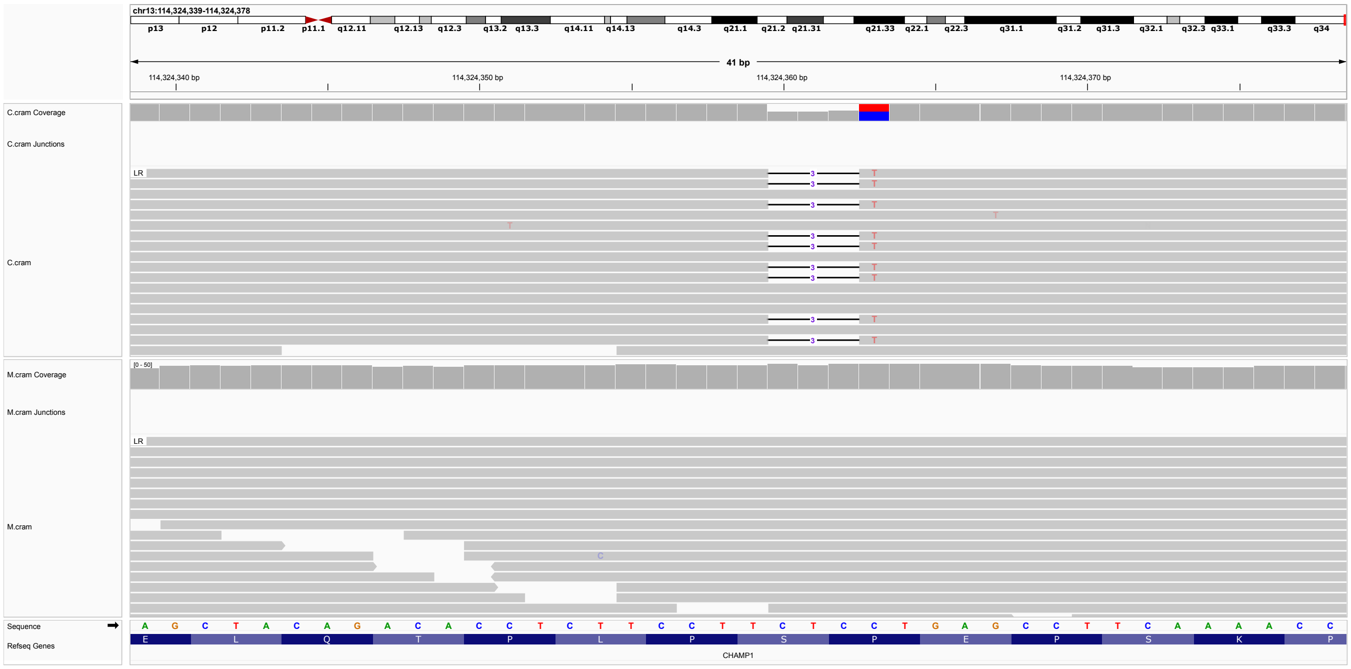


**Figure S52: P52.** Single-nucleotide deletion in *CHAMP1*, 13-114324359-TC-T, ENST00000361283.4, c.518del, p.(Ser173Phefs*46). ACMG classification LP: PVS1_Strong, PM2_Supporting, PP4. Variant not in ClinVar, but numerous P/LP frameshift variants are listed. neurodevelopmental disorder with hypotonia, impaired language, and dysmorphic features (NEDHILD) is caused by de novo heterozygous mutation in the CHAMP1 gene (OMIM:616579). Paternal sample not available.

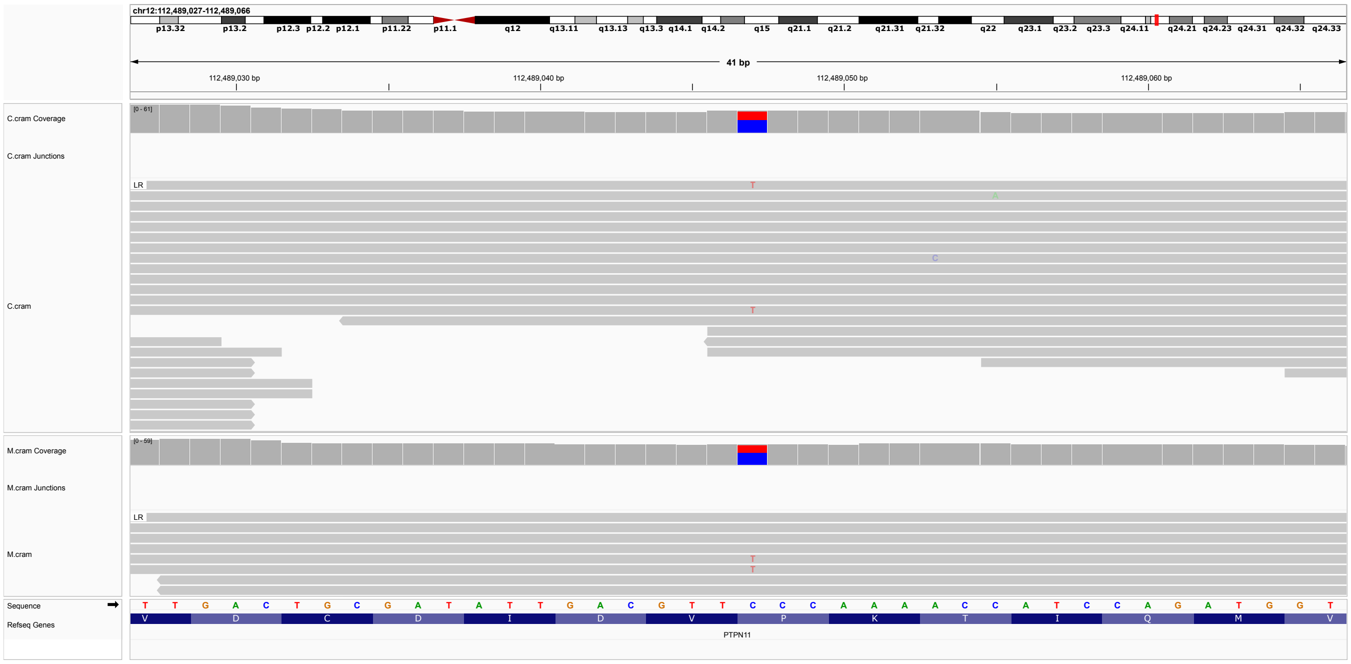


**Figure S53: P53.** Single-nucleotide deletion in *PTPN11*, 12-112489047-C-T, ENST00000351677.6, c.1471C¿T, p.(Pro491Ser). ACMG classification LP: PP5,PP4,PP3,PS1 Variant listed in ClinVar as pathogenic (12 submissions, Variation ID: 40550). Noonan syndrome-1 (NS1) is caused by heterozygous mutation in the PTPN11 gene. Paternal sample not available. Variant additionally identified in mother. Limited clinical information about the mother is available.

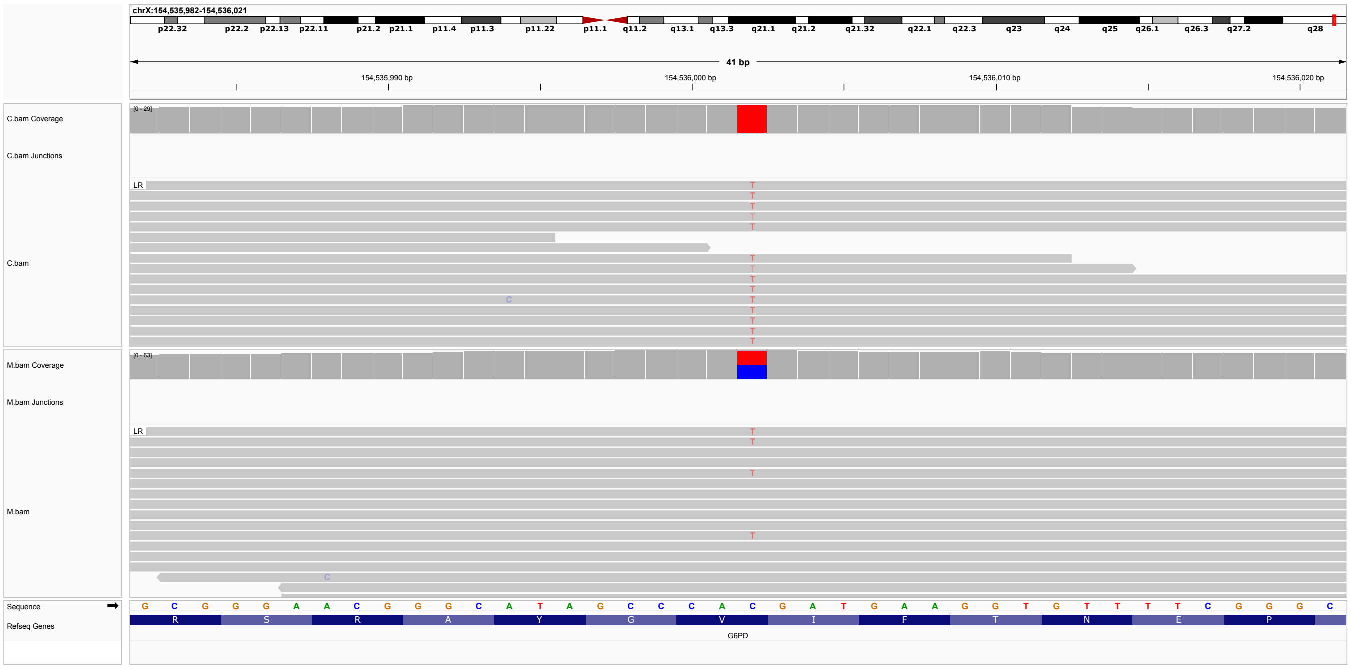


**Figure S54: P54.** Hemizygous single-nucleotide substitution in *G6PD*, X-154536002-C-T, ENST00000393562.10, c.202G¿A, p.(Val68Met). ACMG classification LP: PS4, PS3_SUP, PP3 Variant listed in ClinVar as P/LP (28 submissions, Variation ID: 37123). Paternal sample not available.

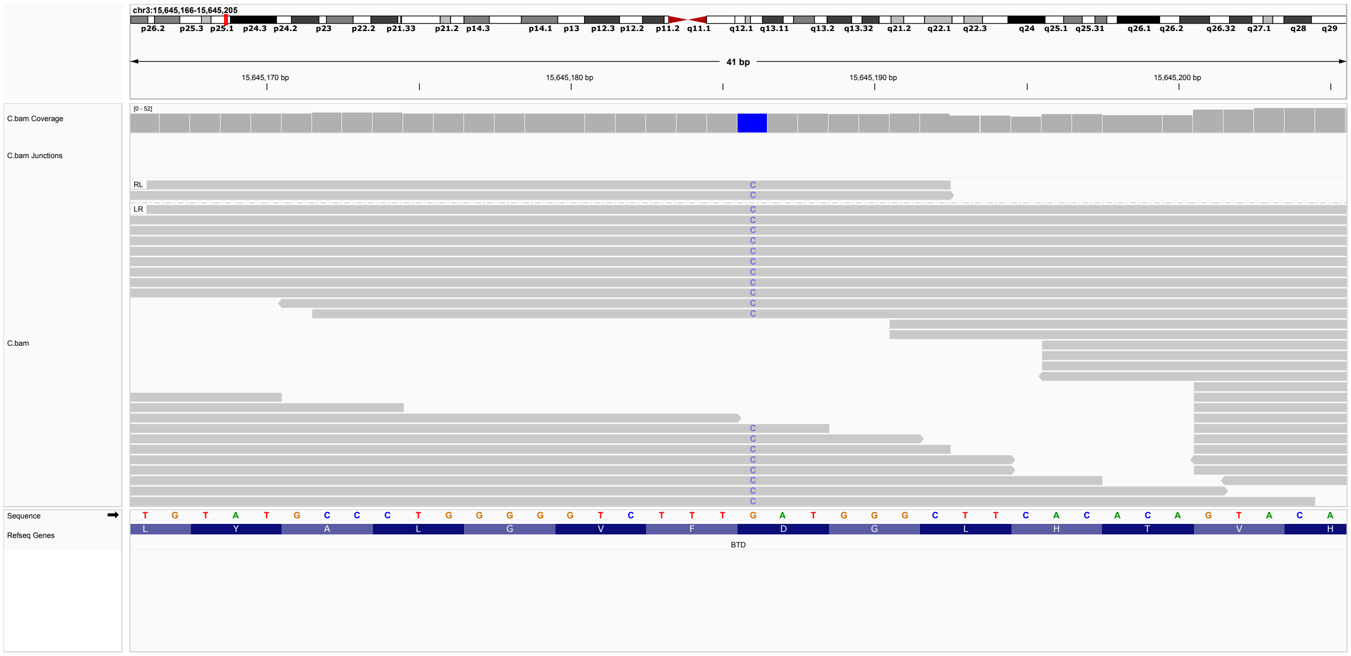


**Figure S55: P55.** Homozygous single-nucleotide substitution in *BTD*, 3-15645186-G-C, ENST00000643237.3, c.1270G¿C, p.(Asp424His). ACMG classification LP: PM1, PM5_Supporting, PM3_Very Strong, PP3, PP5_Strong. Variant listed in ClinVar as P/LP (40 submissions, Variation ID: 1900).

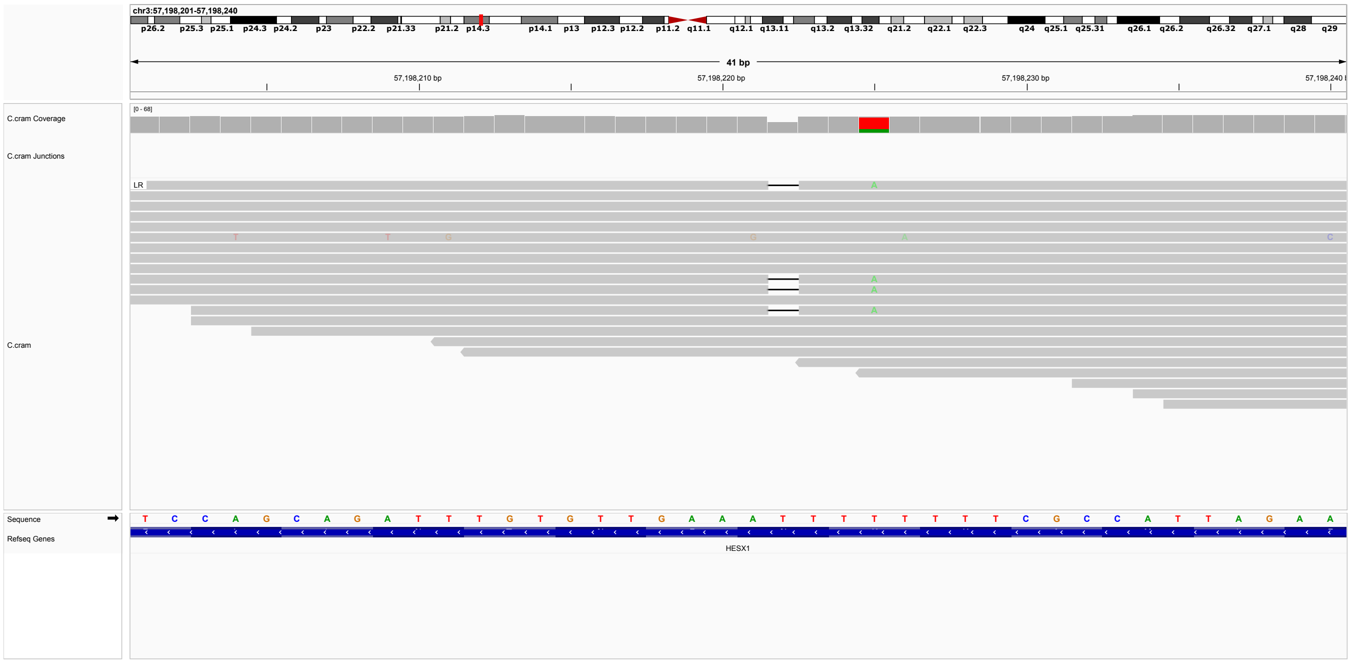


**Figure S56: P56.** Heterozygous single-nucleotide deletion in *HESX1*, 3-57198221-AT-A, ENST00000295934.8, c.533del, p.(Asn178Ilefs*10). ACMG classification P: PVS1,PP4,PP3,BS1 Septooptic dysplasia (OMIM:182230) can be caused by heterozygous, homozygous, or compound heterozygous mutation in the homeobox gene HESX1. No clear genotype-phenotype correlation is obvious among the cases with HESX1 mutations, but there is a trend that the recessive mutations cause more severe phenotypes and appear to be fully penetrant while heterozygous mutations may be associated with milder phenotypes and reduced penetrance [16].

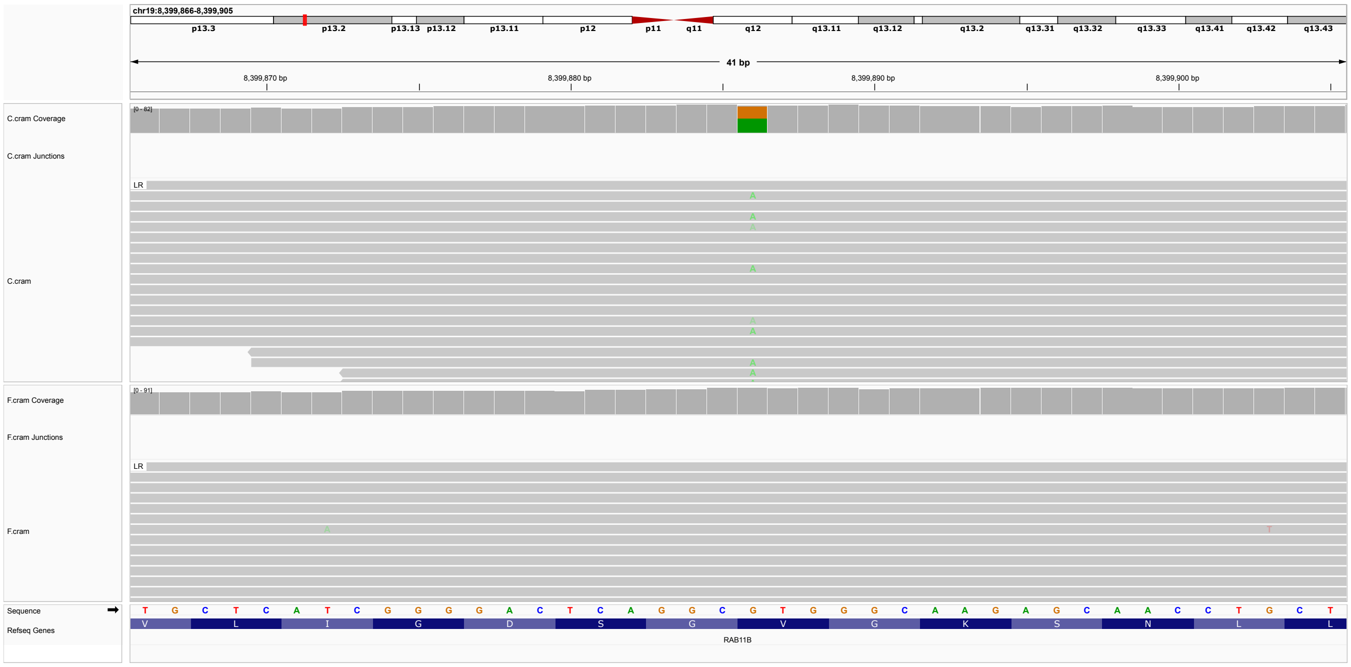


**Figure S57: P57.** Heterozygous single-nucleotide substitution in *RAB11B*, 19-8399886-G-A, ENST00000328024.11, c.64G>A, p.(Val22Met) ACMG classification LP: PM2_Supporting, PP3_Moderate, PP4, PP5_Strong. The variant is listed in ClinVar as P/LP (three submissions, Variation ID: 453254). Neurodevelopmental disorder with ataxic gait, absent speech, and decreased cortical white matter (OMIM:617807) is caused by heterozygous mutation in the RAB11B gene.

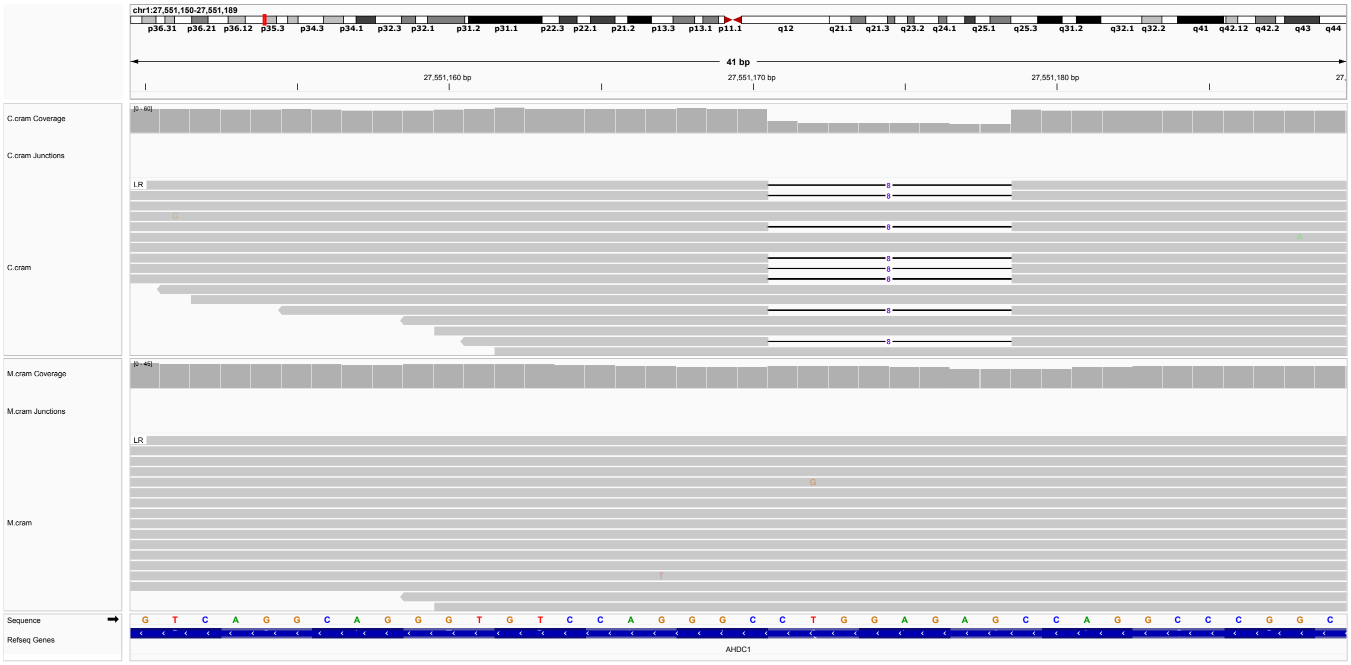

**Figure S58: P58.** Heterozygous 8-nucleotide deletion in *AHDC1*, 1-27551170-CCTGGAGAG-C, ENST00000673934.1, c.938_945del, p.(Ala313Glyfs*7). ACMG classification P: PVS1, PM2_Supporting, PP4. No entry in ClinVar. Xia-Gibbs syndrome (OMIM:615829) is caused by heterozygous mutation in the *AHDC1* gene. Truncating variants in *AHDC1* can cause Xia-Gibbs syndrome [17].

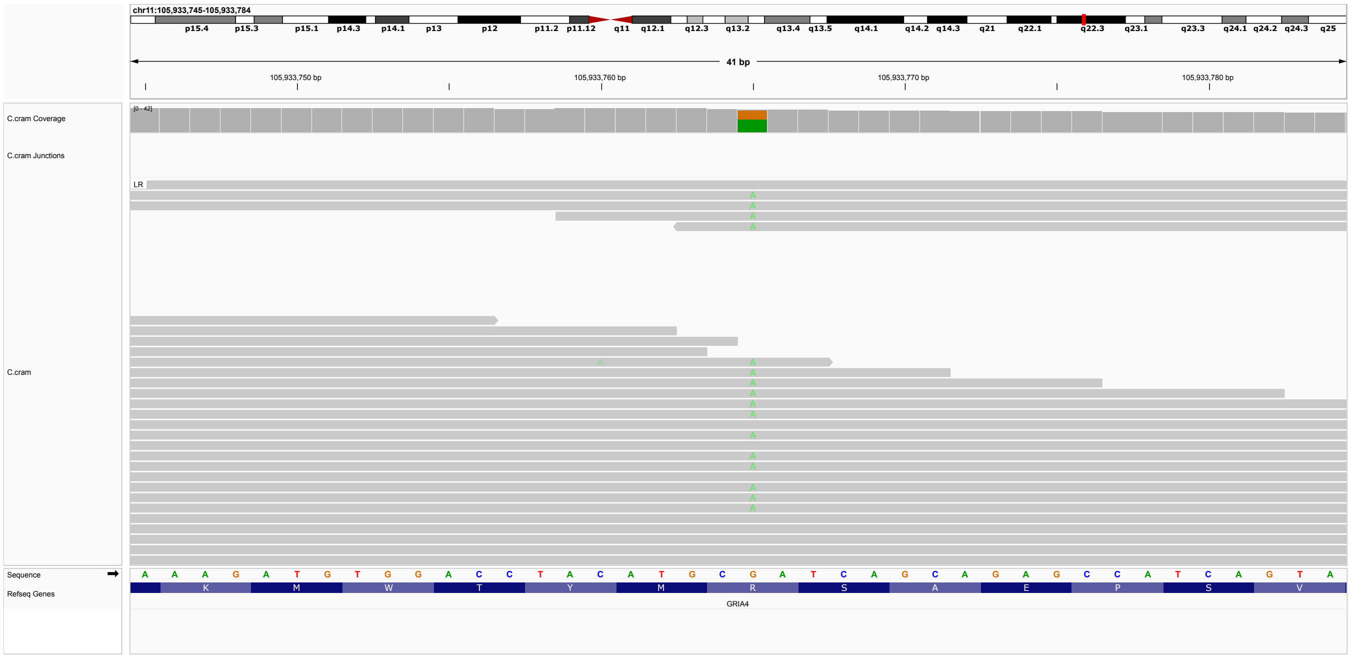


**Figure S59: P59.** Heterozygous 8-nucleotide deletion in *GRIA4*, 11-105933765-G-A, ENST00000282499.10, c.2090G>A, p.(Arg697Gln). ACMG classification VUS: PM5_Supporting, PP5, BP4. Variant listed in ClinVar as pathogenic (One submissions, Variation ID: 2500997). Neurodevelopmental disorder with or without seizures and gait abnormalities (OMIM:617864) is caused by heterozygous mutation in the GRIA4 gene.

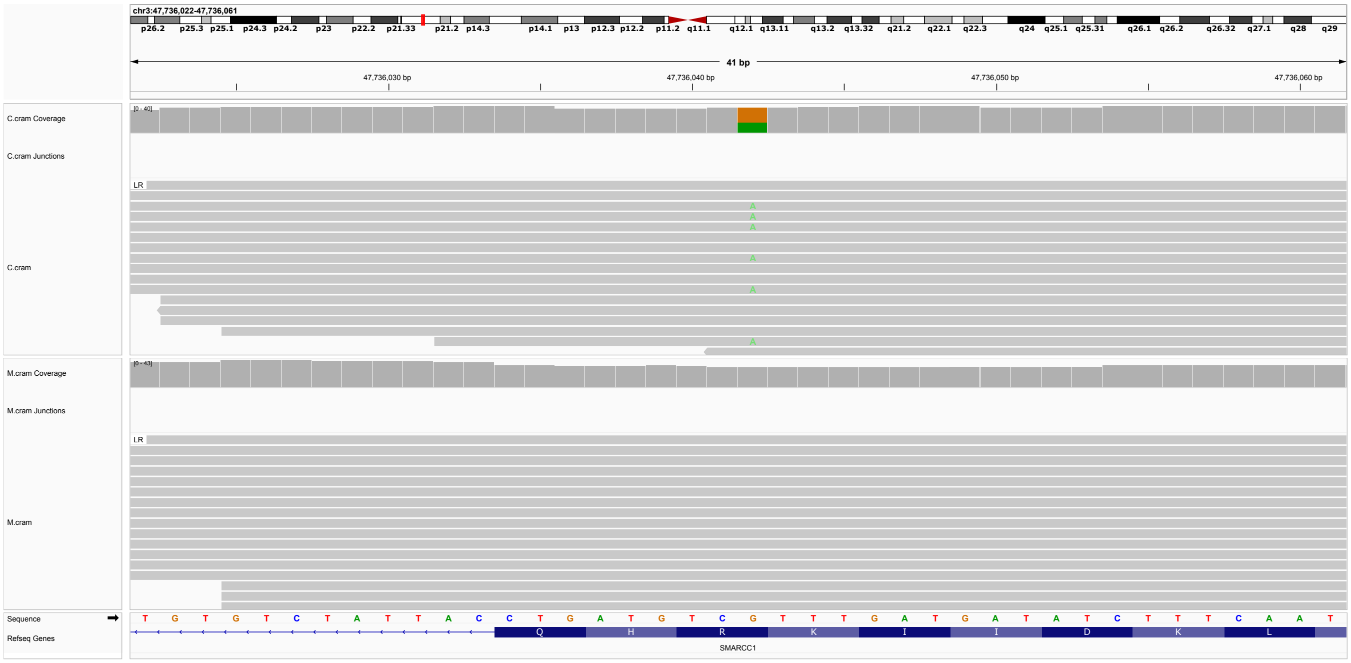

**Figure S60: P60.** Heterozygous single-nucleotide substitution in *SMARCC1*, 3-47736042-G-A, ENST00000254480.10, c.568C>T, p.(Arg190*). ACMG classification LP: PVS1, PM2. Not listed in ClinVar. Susceptibility to congenital hydrocephalus-5 (OMIM:620241) is conferred by heterozygous mutation in the SMARCC1 gene. The affected child was not to have Aqueductal stenosis. Individuals with aqueductal stenosis have been observed to have cerebral palsy PMID: 29588243. Paternal sample not available.

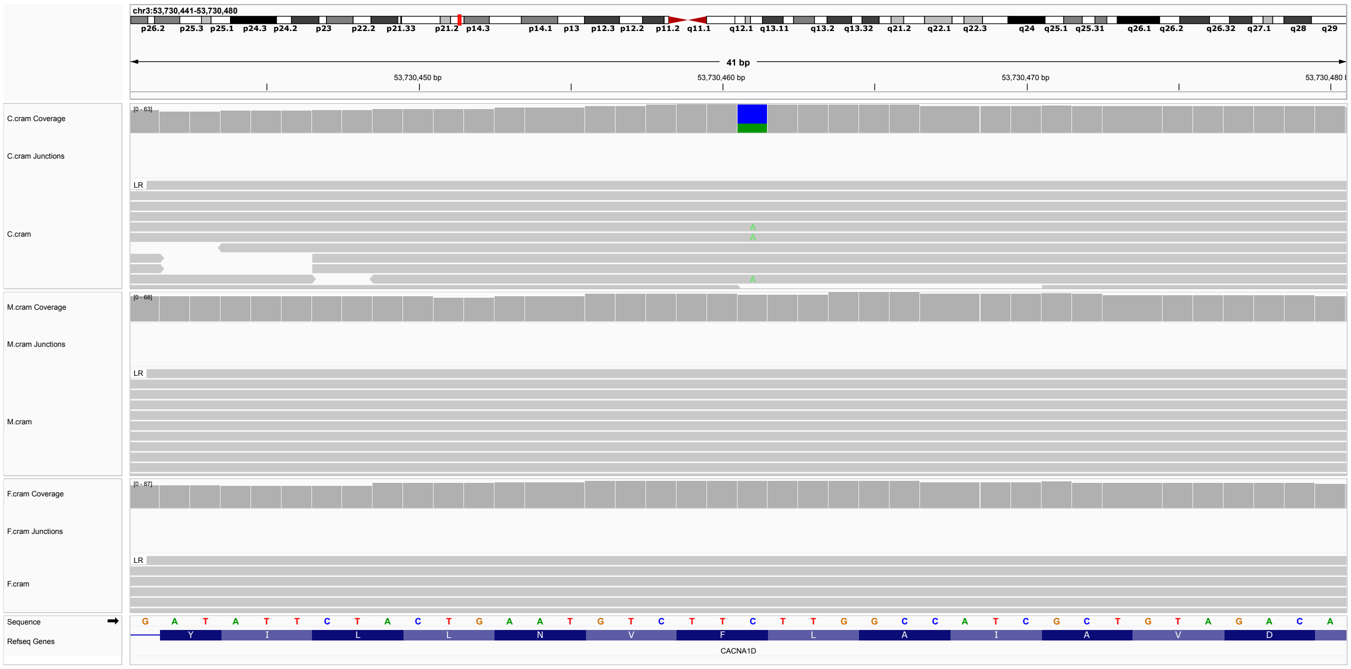


**Figure S61: P61.** De novo heterozygous single-nucleotide substitution in *CACNA1D*, 3-53730461-C-A, ENST00000288139.11, c.2301C¿A, p.(Phe767Leu). ACMG classification P: PS1_Supporting, PS2, PM1, PM2_Supporting, PP3_Moderate, PP4. Variant not listed in ClinVar. Primary aldosteronism with seizures and neurologic abnormalities (OMIM:615474) is caused by heterozygous mutation in the CACNA1D gene At least two children diagnosed with cerebral palsy and de novo *CACNA1D* variants have been published [18].

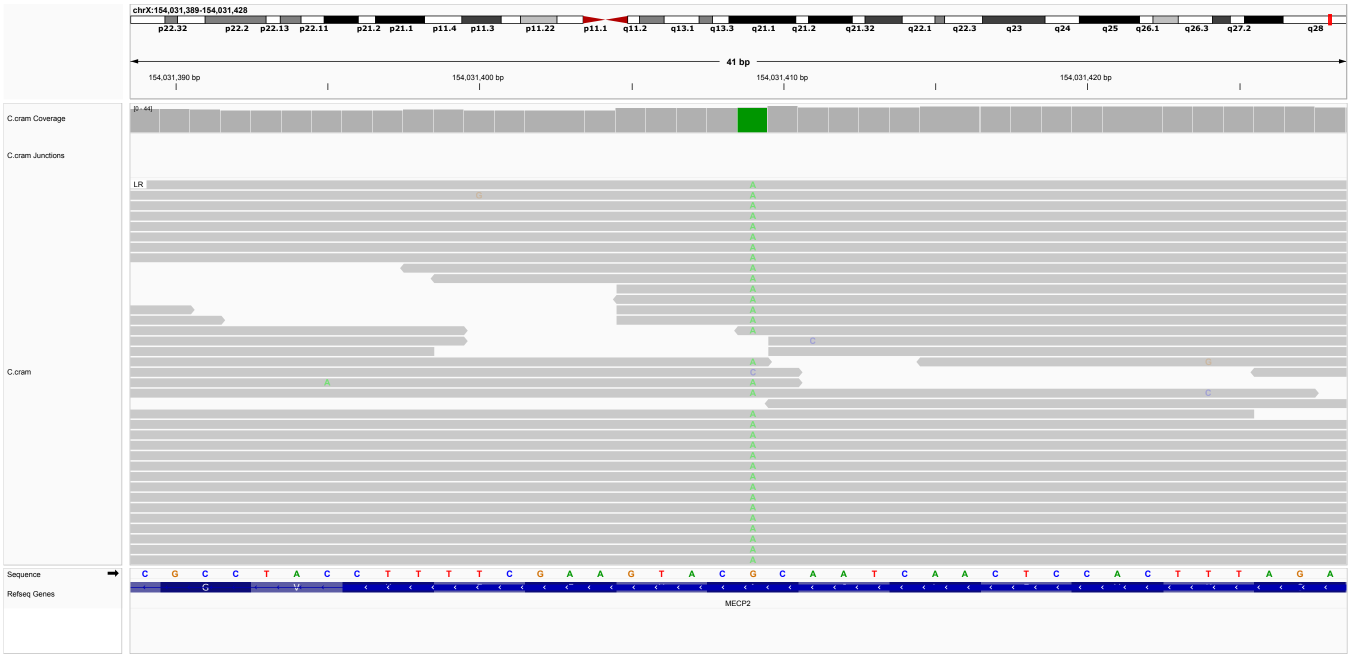


**Figure S62: P62.** hemizygous single-nucleotide substitution in *MECP2*, X-154031409-G-A, ENST00000303391.11, c.419C¿T, p.(Ala140Val). ACMG classification LP: PM1, PP3_Moderate, PP4, PP5_Strong, BP1. The variant is listed in ClinVar as P/LP (24 submissions, Variation ID: 11823) Certain MECP2 variants are associated with Intellectual developmental disorder, X-linked syndromic 13 (OMIM:300055). Parental samples not available. This variant has been published in males with this diagnosis.PMID: 11007980 PMID: 11309367 PMID: 12325019 PMID: 11885030 (TODO summariz).

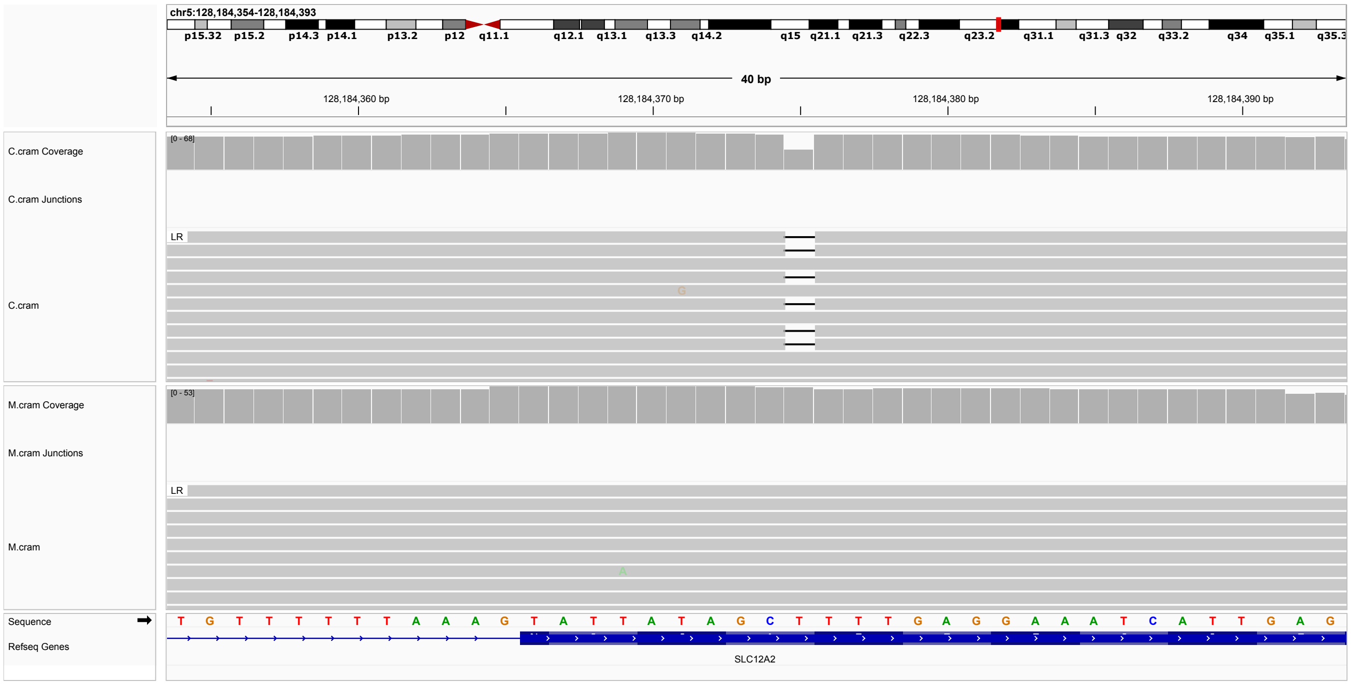


**Figure S63: P63.** hemizygous single-nucleotide deletion in *SLC12A2*, 5-128184374-CT-C, ENST00000262461.7, c.3312del, p.(Phe1104Leufs*23). ACMG classification P: PVS1, PP4, PP5_Strong. The variant is listed in ClinVar as P (3 submissions, Variation ID: 2062696). The c.3312delT (p.F1104Lfs*23) alteration, located in exon 25 (coding exon 25) of the SLC12A2 gene, consists of a deletion of one nucleotide at position 3312, causing a translational frameshift with a predicted alternate stop codon after 23 amino acids. Pathogenic variants in SLC12A2 are associated with Deafness, autosomal dominant 78 (OMIM:619081, DFNA78), Delpire-McNeill syndrome (OMIM:619083) Kilquist syndrome (OMIM:619080). SLC12A2 functions as a dimer and has several isoforms; only one isoform contains exon 21, and this isoform is almost exclusively expressed in the inner ear/cochlea. This isoform is necessary for homeostasis of the endolymph. To date, the published pathogenic variants causing DFNA78 are missense mutations located within exon 21 or in the 3' splice site of exon 21 [19]. Therefore, the current variant does not indicate DFNA78. Kilqvist syndrome is autosomal recessive. Premature truncation codon variants have been identified in individuals with Delpire-McNeill syndrome (e.g., [20]), a condition that can display delayed development and spasticity. Paternal sample not available.

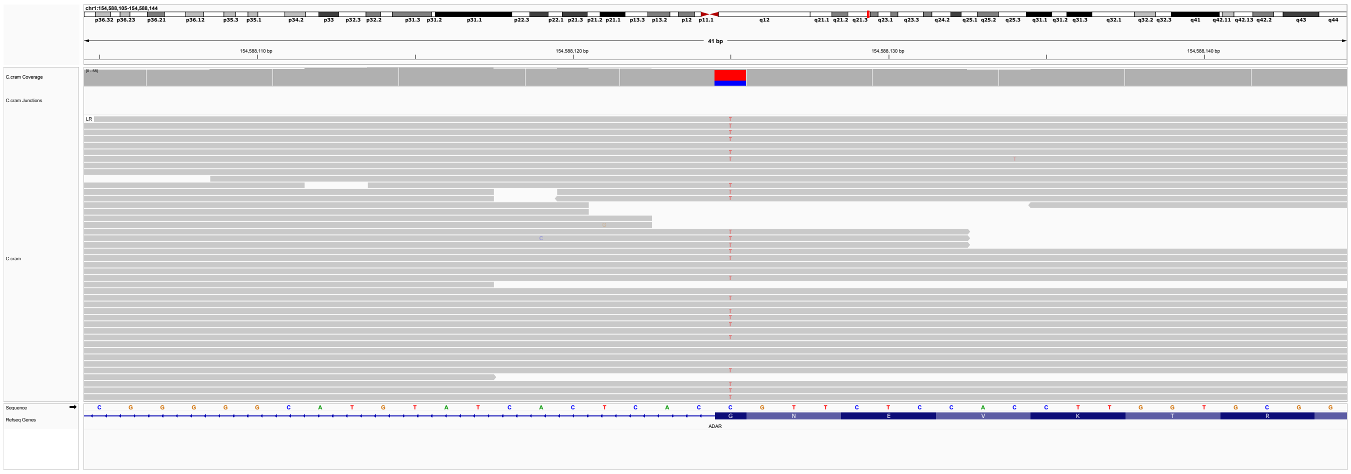

**Figure S64: P64.** hemizygous single-nucleotide deletion in *ADAR*, 1-154588125-C-T, ENST00000368474.9, c.3019G>A, p.(Gly1007Arg). ACMG classification LP: PS1_Moderate, PM2_Supporting, PP3_Moderate, PP4. The identical variant is listed in ClinVar as P/LP (16 submissions, Variation ID: 39458). Although Aicardi-Goutieres syndrome-6 is generally transmitted as an autosomal recessive trait (OMIM:615010; AGS6), specific heterozygous autosomal dominant pathogenic variants in ADAR have been described to cause AGS6. This variant has been reported in multiple individuals with autosomal dominant Aicardi-Goutieres syndrome: individuals AGS150 and AGS474 from [21]; patients 7 and 8 from citeLivingston2014. In another study, the variant was found to occure de novo in four patients, and was inherited in three families in association with marked phenotypic variability [22]. ADAR variants have also been identified in individuals with CP, including five cases in [23]. One of these cases was reported with the identical heterozygous variant (SNVs ID 407), and in another case (SNVs ID 42) the indentical variant was reported in heterozygosity with a second missense variant. In a further publication, monoallelic p.(G1007R) variant was reported tpo be “an important mimic of hereditary spastic paraplegia and cerebral palsy”[24]. Parents not available.

**Figure S65: P65.** Deletion of Exons 8-15 in *CTNNB1* diagnosed by external Invitae report received after inclusion in this study.

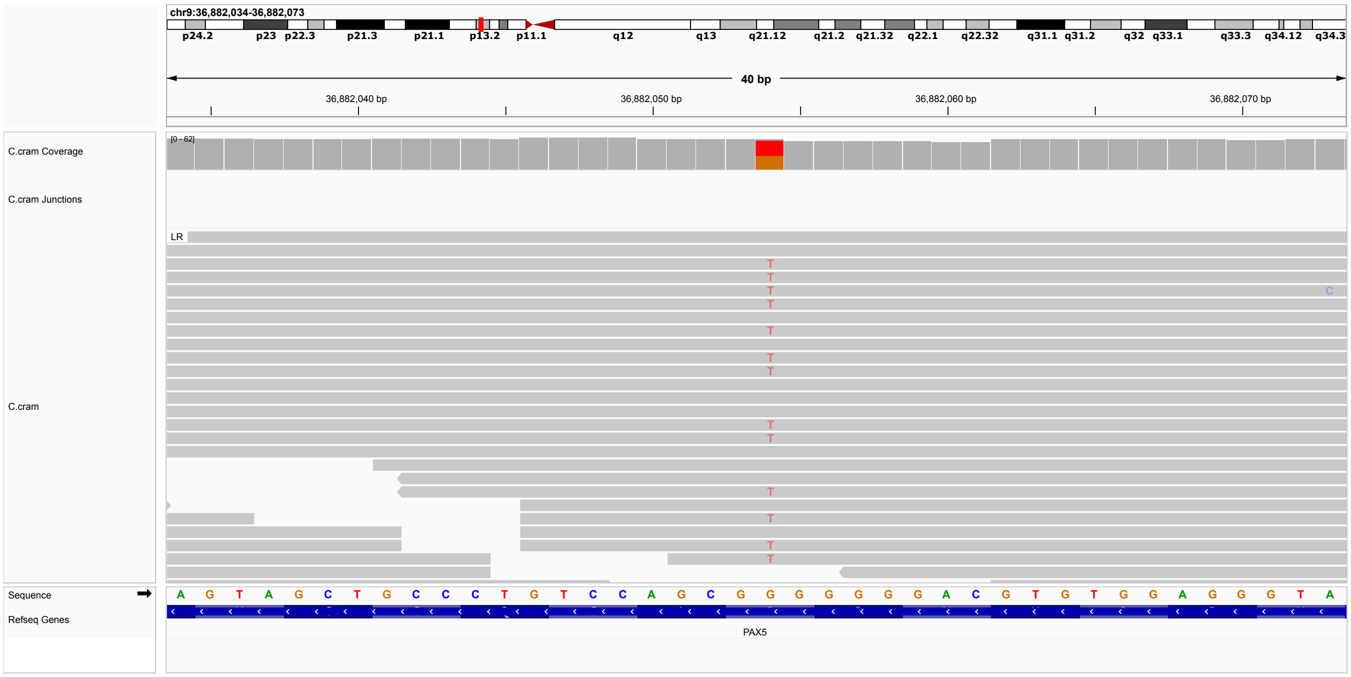


**Figure S66: P66.** hemizygous single-nucleotide deletion in *PAX5*, 9-36882054-G-T, ENST00000358127.9, c.962C>A, p.(Pro321His). ACMG classification: LP (PS1, PP3, PP4). The identical variant is listed in ClinVar as P/LP (1 submission, Variation ID: 1210159). Here the affected individual was said to have Number of individuals with the variant: 2 Clinical Features: Global developmental delay (present) , Learning difficulties (present) , Attention deficit hyperactivity disorder (present) , Motor neuropathy (present) , Seizures (present) , Thinning of the corpus callosum (present) , Delayed speech (present). PAX5 is not listed as a disease-associated gene in OMIM, but a novel neurodevelopmental syndrome associated with PAX5 haploinsufficiency was published in 2022 [25], and the identical variant was also published in that article. We interpret our finding as being most consistent with PAX5 being a novel disease-associated gene. Parental samples were not available.

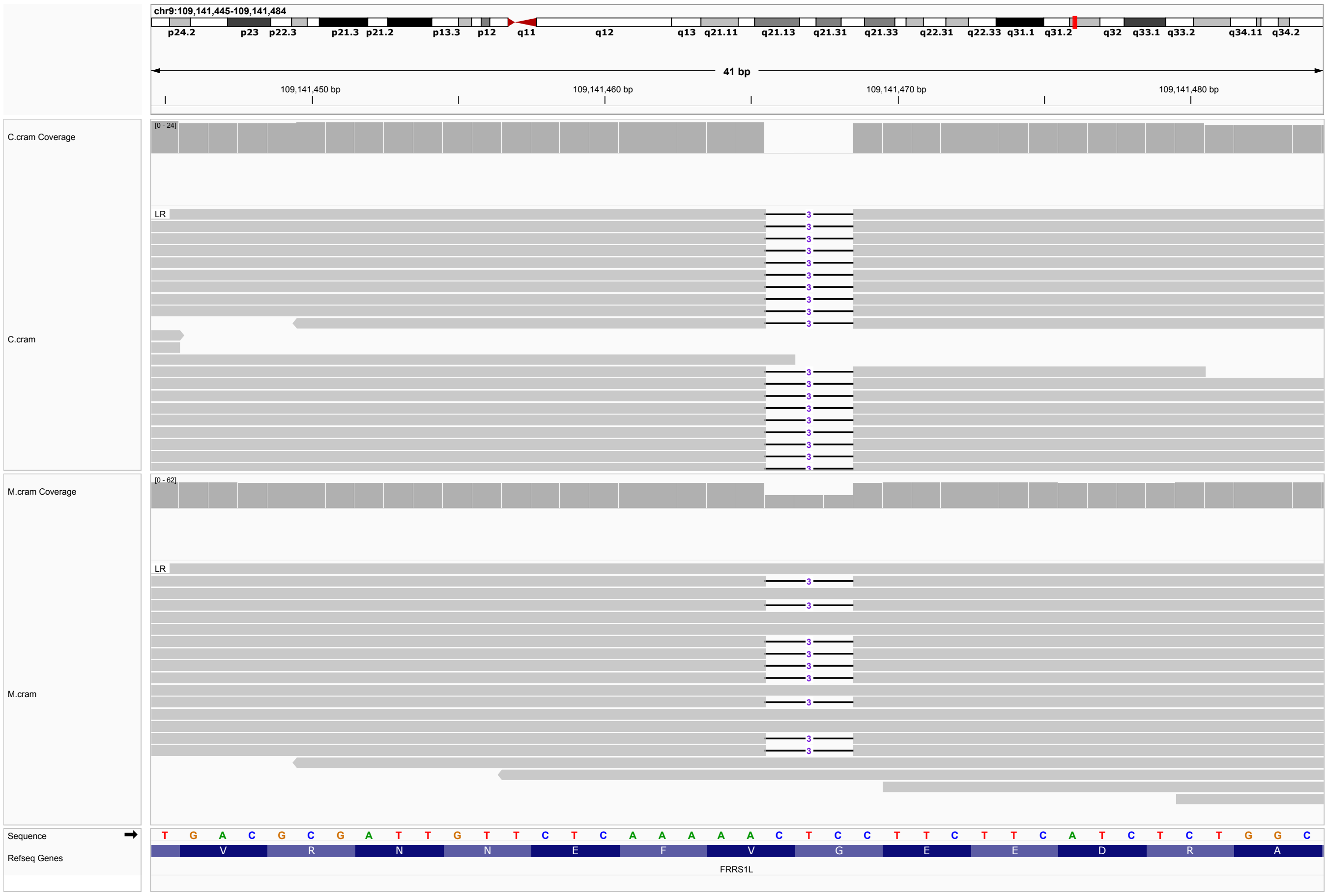


**Figure S67: P67.** homozygous three-nucleotide deletion in *FRRS1L*, 9-109141465-ACTC-A , ENST00000561981.5, c.584_586del, p.(Gly195del). ACMG classification: LP (PM4, PP5_Strong). The variant is listed in ClinVar as P/LP (Variation ID: 218153, 7 of 10 submissions supporting). Variants in are observed in Developmental and epileptic encephalopathy 37 (DEE37; OMIM:616981). The identical variant was published 21 individuals with DEE37 [26, 27, 28], none of whom were noted to have CP. Paternal sample was not available. The variant was identified in homozygous state in the affected child and in heterozygous state in the mother, and can be inferred to be heterozygous in the father.

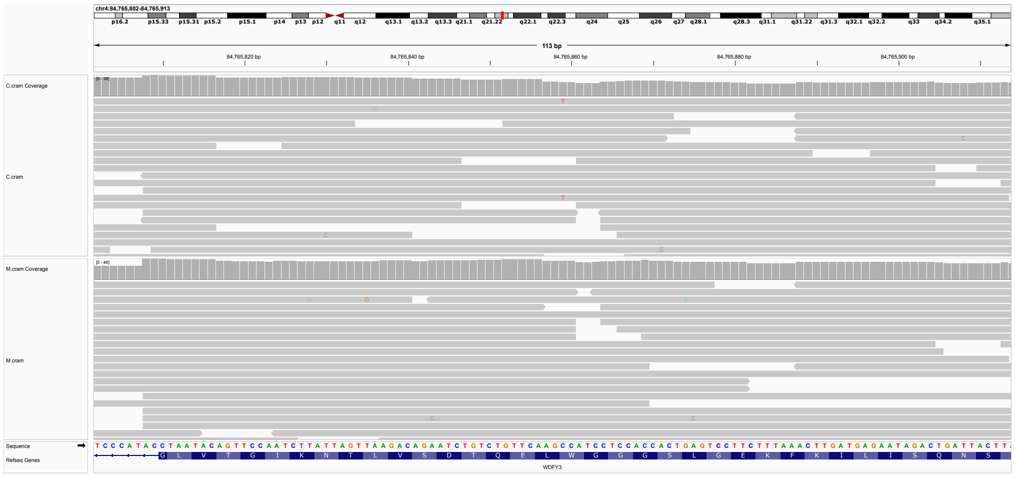


**Figure S68: P68.** Heterozygous stop-gained variant in *WDFY3*, 4-84765859-C-T, ENST00000295888.9, c.5139G>A, p.(Trp1713*). ACMG classification LP: PVS1, PM2_Supporting]. Paternal sample not available. Only 4 of 25 reads display the variant, raising the possibility of a mosaic. Heterozygous, mostly de novo protein-truncating and missense variants in WDFY3 have been reported as a monogenic cause of mild and non-specific neurodevelopmental delay. Overlapping symptoms included neurodevelopmental delay, intellectual disability, macrocephaly, and psychiatric disorders (autism spectrum disorders/attention deficit hyperactivity disorder) [29, 30, 31].

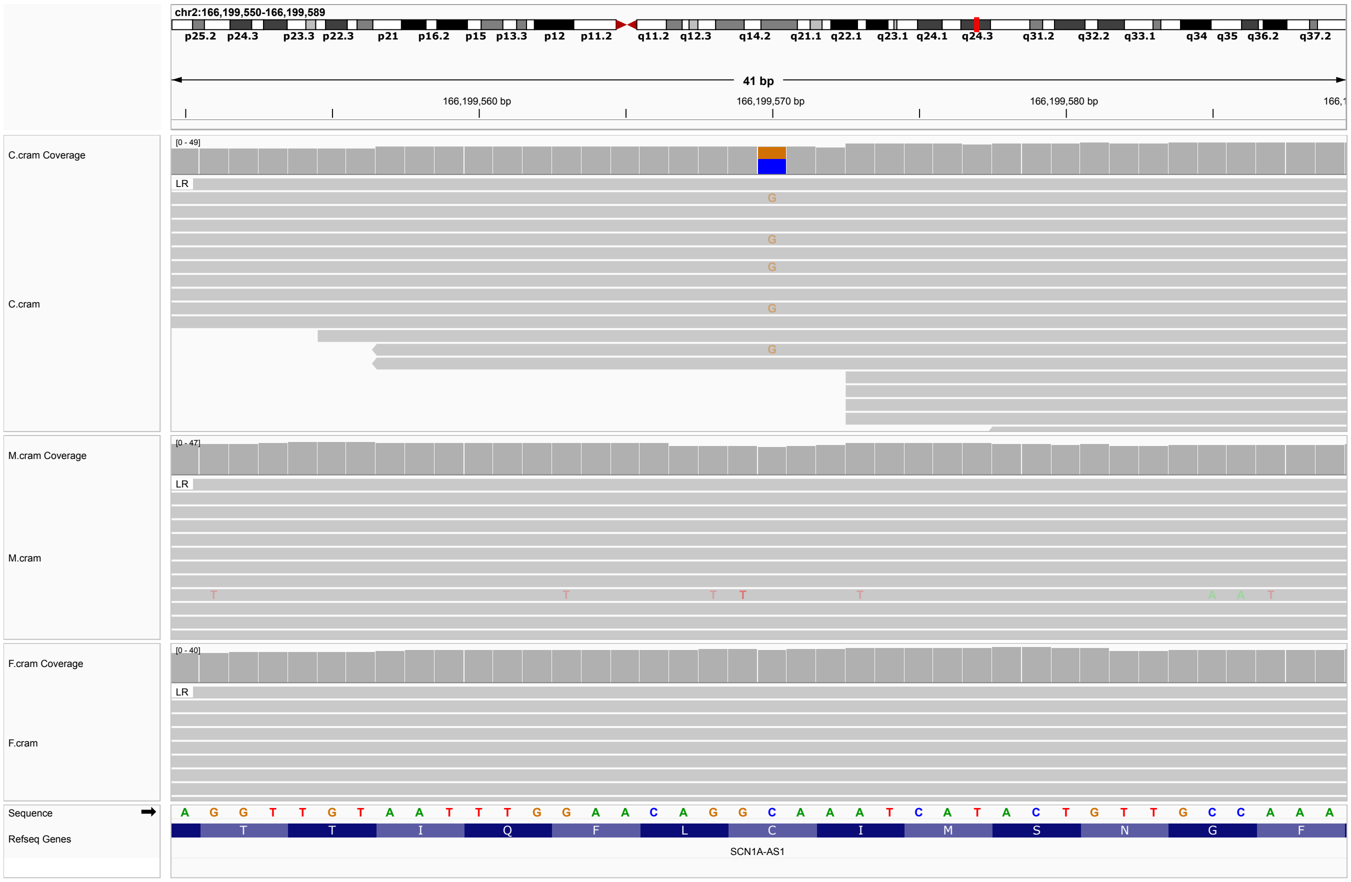


**Figure S69: P69.** De novo heterozygous missense variant in *SCN9A*, 2-166199570-C-G, ENST00000642356.2, c.5069G¿C, p.(Cys1690Ser) ACMG classification LP PS2, PM2, PP3, PP4, BP1. Heterozygous variants in *SCN9A* are associated with Paroxysmal extreme pain disorder (OMIM:167400) and Small fiber neuropathy (OMIM:133020). No corresponding clinical manifestations were reported. However, an *SCN9A* missense variant was reported as causal in an individual with CP and Epilepsy, generalized with febrile seizures plus, type7 Dravet syndrome[32]. An additional *SCN9A* missense variant was identified in an infant with clinical manifestations of moderate to severe hypoxic-ischemic encephalopathy without significant risk factors for perinatal asphyxia[33].

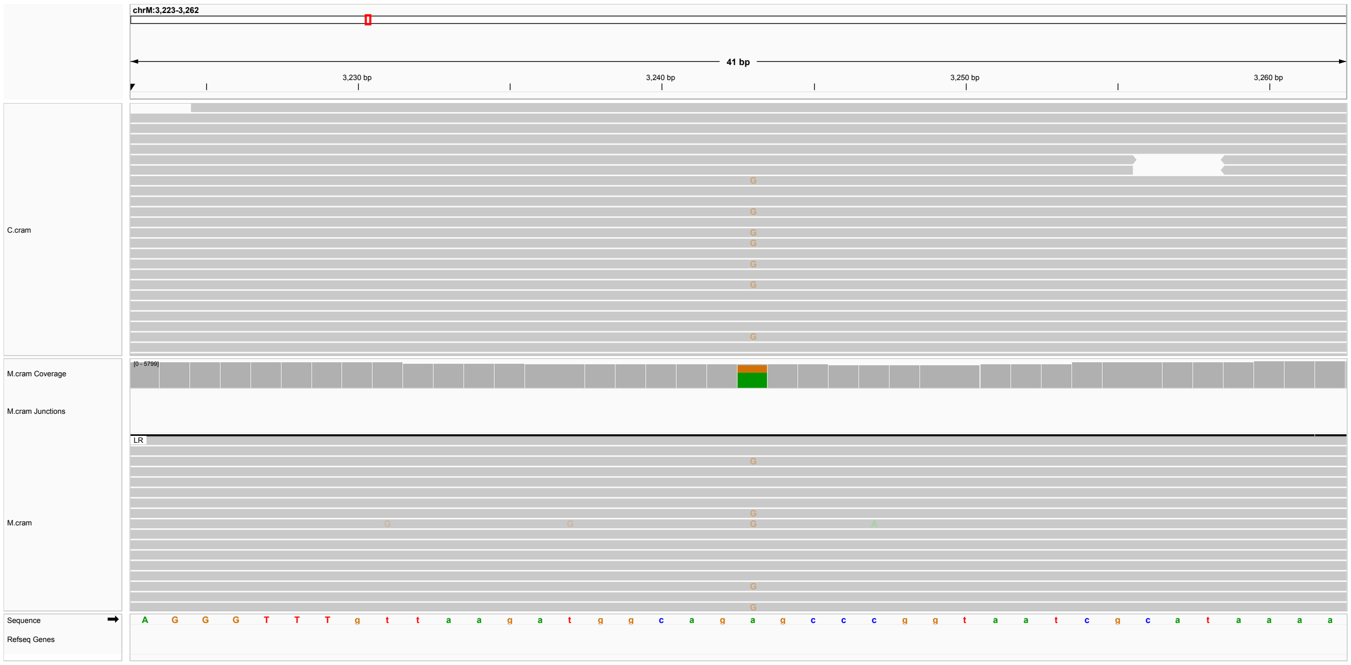


**Figure S70: PC8.** Mitochondrial variant, MT-3243-A-G, NC_012920.1(MT-TL1), m.3243A>G. ACMG classification LP: PM2_Supporting, PP4, PP5_Strong. Variant is listed in ClinVar as P/LP (28 submissions, Variation ID: 9589). The heteroplasmy was 17% in the child (632 of 3134 reads), and 32% in the mother (1490 of 3212 reads). According to CLinVar, The m.3243A>G variant (rs199474657) disrupts the mitochondrial tRNA for leucine (UUR), and is one of the most common pathogenic variants in the mitochondrial genome. The clinical presentation associated with this variant is highly variable and depends on the total percentage of abnormal mitochondria and tissue-specific distribution. The m.3243A¿G variant was initially identified in patients with mitochondrial myopathy, encephalopathy, lactic acidosis, and stroke-like episodes (MELAS) syndrome, and recent epidemiological studies found that the most frequent presentation is maternally inherited diabetes and deafness. Other clinical manifestations include hypertrophic cardiomyopathy, ataxia, basal-ganglia calcifications, and ophthalmoplegia. According to [34], certain interpretation of pathogenicity is possible with heteroplasmy of 50 percent or above. This specific variant is a common cause of MELAS syndrome [35], and individuals with MELAS have been diagnosed with cerebral palsy [36]. We therefore interpret this variant as a candidate for the etiology of cerebral palsy in this individual.

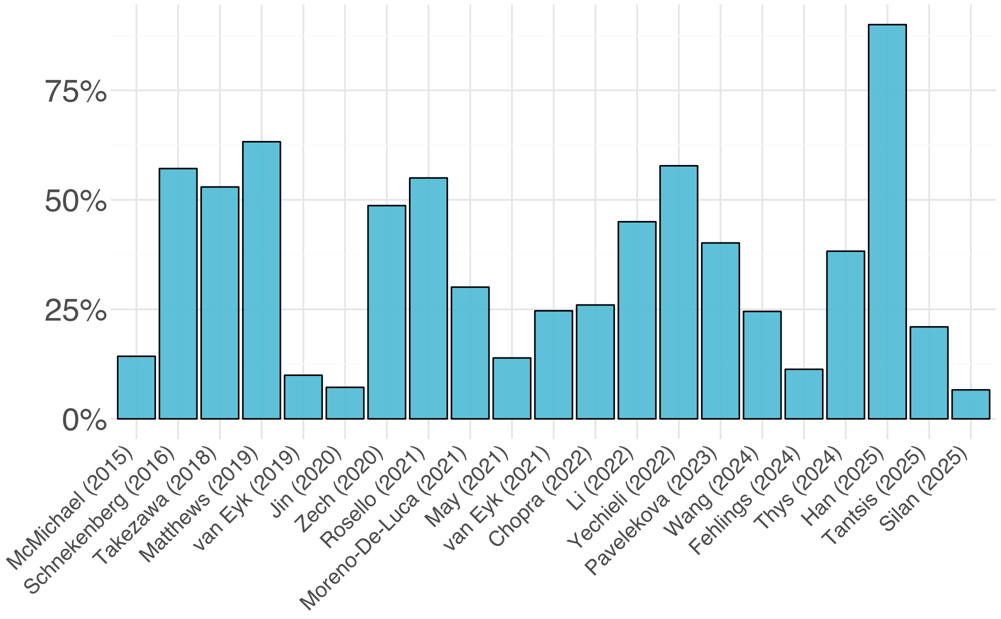


**Figure S71: Diagnostic yield**. The bar plot shows the diagnostic yield reported for the 21 cohorts analyzed in this work [37, 38, 39, 40, 41, 42, 43, 44, 23, 45, 46, 47, 48, 49, 50, 51, 52, 53, 54, 55, 56].

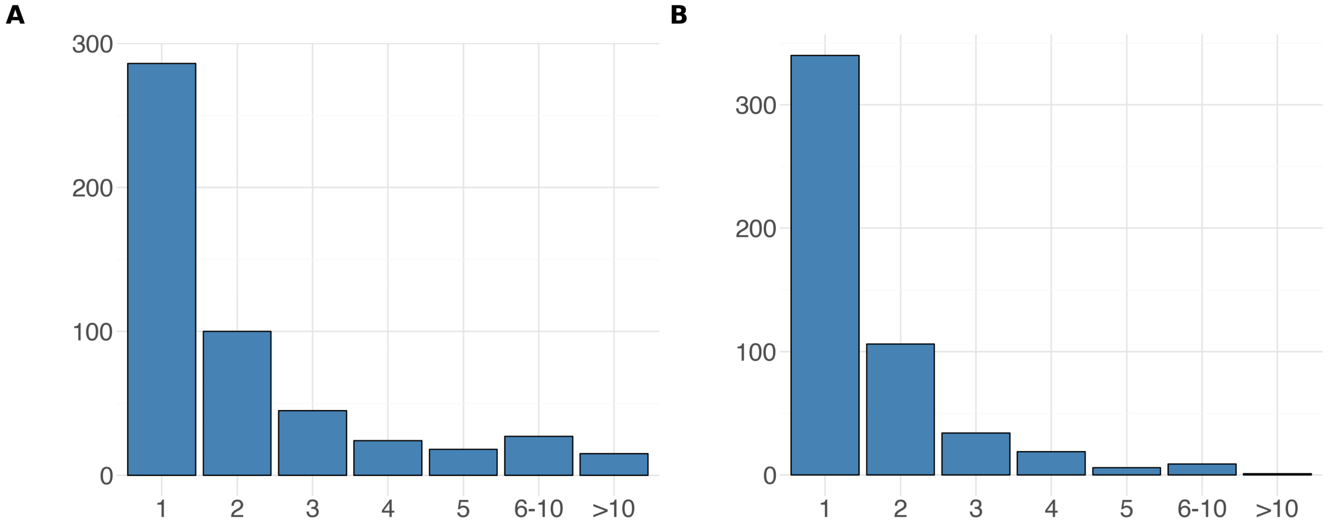


**Figure S72: Distribution of genes**. A) Total number of times a gene was reported to have a causal P/LP variant across the entire 21 CP NGS cohorts. B) Total number of cohorts in which a gene was reported.
